# Fifty-one novel and replicated GWAS loci for polyunsaturated and monounsaturated fatty acids in 124,024 Europeans

**DOI:** 10.1101/2022.05.27.22275343

**Authors:** Michael Francis, Yitang Sun, Huifang Xu, J. Thomas Brenna, Kaixiong Ye

## Abstract

Circulating polyunsaturated and monounsaturated fatty acid (PUFA and MUFA) levels, whose imbalances co-occur with human metabolic diseases, have strong heritable components. We performed the largest genome-wide association study (GWAS) to-date on fourteen PUFA and MUFA phenotypes, measured by nuclear magnetic resonance in plasma. We identified 612 significant locus-phenotype associations (115 unique loci; *P* < 1.678×10^-8^) in a European cohort from UK Biobank (UKB-EUR; n=101,729). Replication of five phenotypes (omega-3, omega-6, DHA, LA, MUFAs) was conducted in two external European studies: FinMetSeq (n=8,751) and a meta-analysis by Kettunnen et al. (n=3,644-13,544). Meta-analysis of these three studies yielded 254 significant locus-phenotype associations (109 unique loci; *P* < 2.439×10^-8^); we identified 87 novel loci, 51 of which were replicated. A transcriptome-wide association study of the UKB-EUR cohort revealed an additional twelve novel loci. This study improves our understanding of the genetic architecture of unsaturated fatty acids and can inform future genotype-based dietary interventions.

## Introduction

Polyunsaturated fatty acids (PUFAs) are dietary fats containing two to six double bonds along linear carbon chains from 14 to 22 carbons in length. Imbalances of tissue PUFAs are involved in the pathophysiology of a broad array of diseases, including cardiovascular disease, cancer, depression, and dementia ^1-3^. Omega-3 long-chain PUFAs (n-3 LCPUFAs) have been consistently shown to improve aspects of metabolic syndrome related to the risk factors of cardiovascular disease and obesity, such as insulin resistance, hypertension, and dyslipidemia ^1,4,5^. Omega-6 (n-6) LCPUFAs have been associated with both positive and negative health outcomes ^6^. Excess n-6 linoleic acid (LA) suppresses tissue and circulating n-3 LCPUFAs, due to common enzymes operating on both PUFA families; balance in dietary n-6 and n-3 is necessary to avoid suppression of the functional n-3 LCPUFAs, eicosapentaenoic acid (EPA) and docosahexaenoic acid (DHA) ^7^. Modest overall dietary PUFAs and an n-6/n-3 ratio up to 4/1 have been recommended, while the typical modern industrialized diet has a ratio approximating 15/1 ^8,9^.

Dietary PUFA intake strongly influences circulating fatty acid levels ^10^. Heritability analyses in twin studies and large cohorts have indicated that substantial genetic components also contribute to determining circulating PUFA levels ^11-13^. Previous genome-wide association studies (GWAS) have identified 37 unique genomic loci related to PUFAs and monounsaturated fatty acids (MUFAs; Supplementary Table 1). However, they collectively only explain a small fraction of the phenotypic variance ^12,14^, suggesting more loci may be found in large-sample GWAS. High-throughput nuclear magnetic resonance spectroscopy (NMR) enables rapid large-scale quantification of metabolic biomarkers ^15^. The UK Biobank (UKB) has recently released NMR data of plasma fatty acid levels for over 110,000 participants, presenting an opportunity to identify novel genetic loci associated with PUFAs and MUFAs.

Here, we perform a linear mixed model (LMM) GWAS to identify the genetic variants associated with the fourteen available NMR-derived plasma unsaturated fatty acid phenotypes in UKB participants: n-3 LCPUFAs, n-6 LCPUFAs, DHA, LA, total PUFAs, total MUFAs, the percentages of each of these fatty acid groups per the total amount of fatty acids, as well as the ratios of n-6/n-3 and PUFA/MUFA. Our discovery cohort consisted of UKB participants with NMR PUFA and MUFA trait measurements, who were determined to be genetically European (EUR) by the Pan-UK Biobank project ^16^ (n=101,729). Additional multi-ancestry replication analyses were performed in African (AFR), Central and South Asian (CSA), and East Asian (EAS) UKB participants (n=4,400). Two external EUR studies were used in our replication and meta-analysis: the Finnish Metabolic Sequencing (FinMetSeq) study ^11^ (n=8,751); and a meta-analysis of 14 datasets derived from ten EUR GWAS studies by Kettunnen et al. ^14^ (n=3,644 to 13,544). In our five meta-analyzed traits (n-3, DHA, n-6, LA, MUFAs) we identified 254 significant locus-trait associations, 102 of which were novel and replicated. These consisted of 51 unique, novel and replicated loci across traits (overview in Figure 1). We also performed a transcriptome-wide association analysis in the fourteen discovery traits, which revealed an additional 12 novel and significant loci that were not found in GWAS. We have provided follow-up analyses and functional interpretations to put these significant associations into plausible biological context, and provide a contemporary description of the genetics involved in circulating PUFA and MUFA levels.

**Figure 1.**
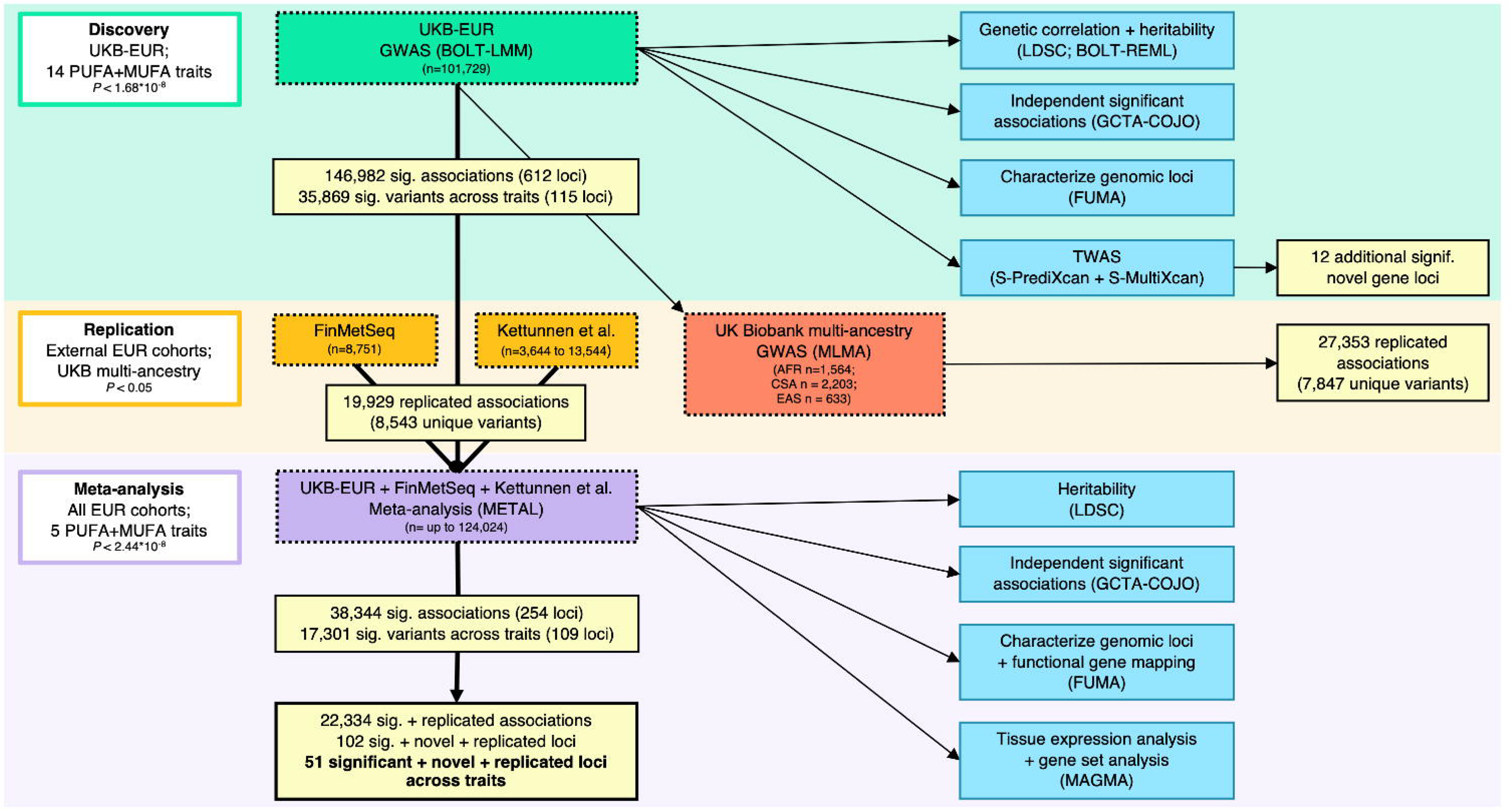
Overview of analyses. Top left moving down: fourteen polyunsaturated fatty acid (PUFA) and monounsaturated (MUFA) traits were analyzed in the UK Biobank European discovery cohort (UKB-EUR). Significant associations were sent to replication in external European cohorts FinMetSeq and Kettunnen et al. for five available PUFA and MUFA traits. These three EUR studies were meta-analyzed, and 22,334 significant and replicated associations were identified across the five traits. Across these meta-analysis results we identified 51 unique, novel, and significant replicated loci. Middle: additional replication was also performed in UKB multi-ancestry cohorts. Right: additional software analyses are shown in blue. FUMA: Functional Mapping and Annotation of Genome-Wide Association Studies; COJO: Genome-wide Complex Trait Analysis Conditional and Joint Association Analysis; LDSC: Linkage Disequilibrium Score Regression; TWAS: transcriptome-wide association analysis.

## Results

### Discovery analysis

We performed a GWAS of NMR-measured polyunsaturated fatty acid (PUFA) and monounsaturated fatty acid (MUFA) traits in individuals of European ancestry (EUR). An overview of our three-stage discovery, replication, and meta-analysis approach can be seen in Figure 1. First, we performed a GWAS discovery analysis using a UKB-EUR discovery cohort composed of 101,729 participants with NMR data, who were designated as genetically EUR by the Pan UKBB project ^16^. Mean age of participants was 56.8 years old and 45.98% were male (Supplementary Figure 1; Supplementary Table 2). Two discovery stage sensitivity models were compared, M1 and M2. The *P*-values were highly correlated between the two models and no residual confounding was observed with either model (Supplementary Figure 2; Supplementary Table 3). The ranges of LD score regression (LDSC) intercepts and genomic control (λ) were 1.00-1.041 and 1.1475-1.2545 for M1 and 0.99-1.045 and 1.1475-1.2 for M2, respectively (Supplementary Table 3). The number of significant variants identified in M2 was always comparable to or greater than the same trait of M1 despite no inflation identified in M2; we inferred this was because our inclusion of relevant covariates reduced residual variability and enhanced the statistical power for variant discovery in M2 ^17^. All results are therefore based on M2.

A total of 15,578,593 variants were tested for associations with all fourteen available PUFA and MUFA traits in our discovery analysis. We found a total of 146,982 significant associations (35,869 unique variants across traits; Supplementary Figure 3; Supplementary Table 3) at the significance threshold corrected for the effective number of traits (*P* < 1.678×10^-8^). Conditional and joint analysis (COJO) identified 968 independent significant associations in this cohort (471 unique variants; Supplementary Tables 4 and 5). We used FUMA to group significant associations into LD blocks and merged loci < 250Kb apart; this yielded 612 genomic risk loci (115 unique loci across traits; Supplementary Tables 4 and 6). There were 404 novel locus-trait associations (95 unique loci across phenotypes) identified in the discovery stage.

### Replication and meta-analysis

Our primary replication analysis utilized two external EUR GWAS studies: FinMetSeq and Kettunen et al. (Supplementary Table 11). These studies contained five out of the fourteen traits analyzed in the discovery stage: n-3, n-6, DHA, LA, and MUFAs. After munging all three EUR studies to ensure high quality alleles and to harmonize alleles to the reference genome, ∼8.7 million variants overlapped between Kettunen et al. and UKB-EUR, and 209,509 variants overlapped between FinMetSeq and UKB-EUR. Across the five available phenotypes, there were 19,929 UKB-EUR associations (8,543 unique variants) replicated at *P* < 0.05 in one of the two external EUR studies. Of these, 615 associations (266 unique variants) were replicated in both studies (Supplementary Table 11).

We performed additional replication analyses across the three UKB multi-ancestry groups with adequate sample sizes, to evaluate the reproducibility of associations found in the UKB-EUR cohort. No significant phenotypic differences were found between UKB ancestry groups in any of fourteen PUFA and MUFA traits (Supplementary Figure 1; Supplementary Table 2). Mixed linear model-based association analysis (MLMA) was performed in UKB African (UKB-AFR), Central and South Asian (UKB-CSA) and East Asian (UKB-EAS) cohorts (Supplementary Figure 4). Counts of UKB-EUR associations replicated (*P* < 0.05) were: UKB-AFR 5,327 (2,358 unique variants), UKB-CSA 16,560 (5,179 unique variants), and UKB-EAS 5,466 (2,113 unique variants) (Supplementary Table 12). Out of the 612 significant loci for UKB-EUR associations, 170 were replicated in ≥ 1 UKB multi ancestry group (Supplementary Tables 6 and 12). Interestingly, despite having a smaller sample size, UKB-EAS contained more replicated UKB-EUR loci than UKB-AFR (46 vs. 31).

Meta-analysis was performed on variants which appeared in at least two out of three EUR studies, and ∼10,200,000 variants were tested. Across the five PUFA and MUFA traits there were 38,344 significant associations (*P* < 2.439×10^-8^; 17,301 unique variants across traits; Figure 2; Supplementary Table 13). LDSC intercepts for the five meta-analyzed traits ranged from 1.013 (SE = 0.0079) for DHA to 1.04 (0.011) for MUFAs, indicating that there was not inflation or residual confounding (Supplementary Table 13). COJO revealed 402 independent significant associations (258 unique variants; Supplementary Tables 13 and 14). Of these 402 significant COJO associations, 265 were replicated in at least one external EUR study (*P* < 0.05). When grouping all significant meta-analysis variants into loci, we found 254 significant loci (109 unique across traits; Supplementary Tables 13 and 15). Of the 254 grouped meta-analysis loci, 171 were replicated in at least one external EUR study (Supplementary Table 18).

**Figure 2.**
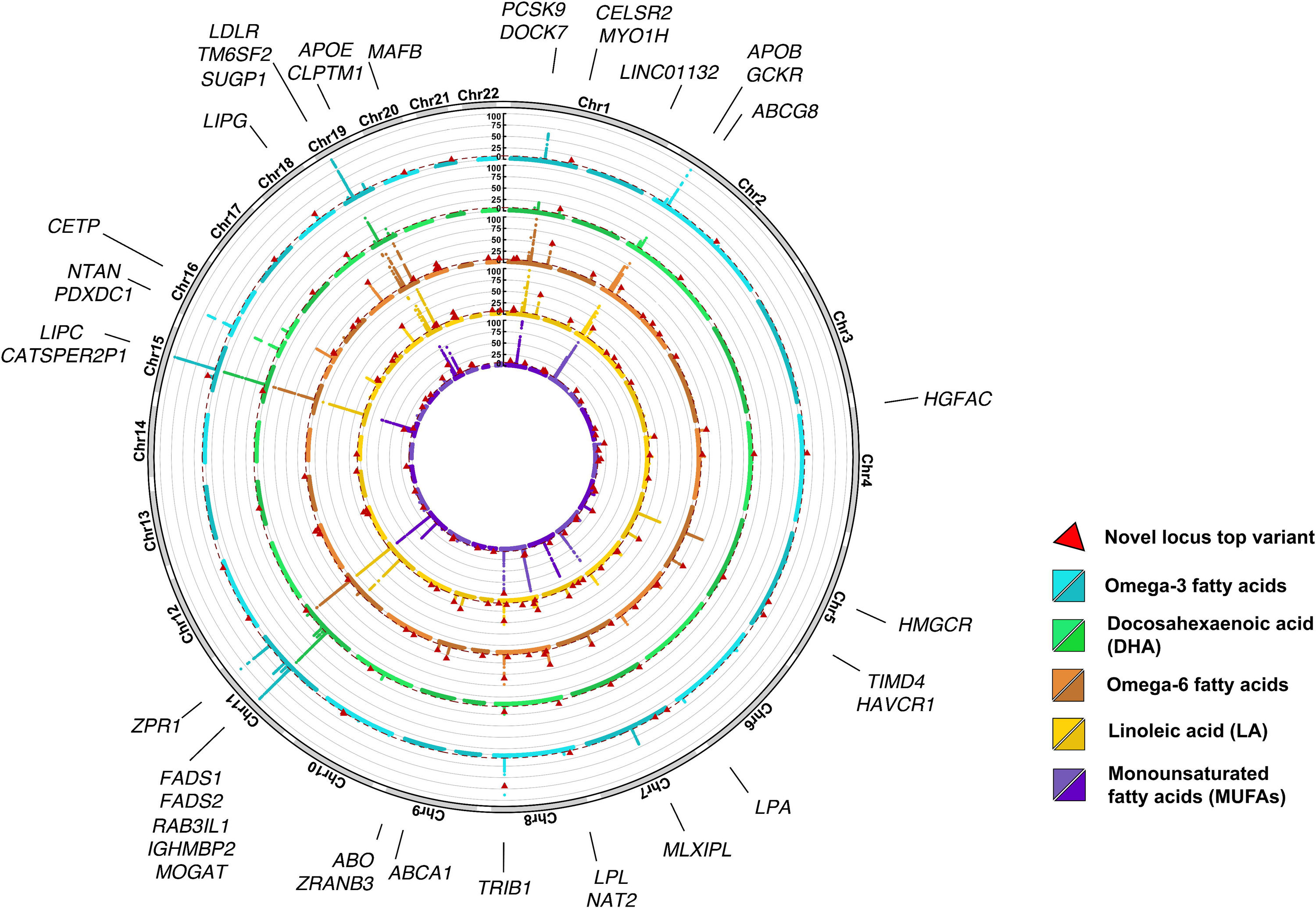
Circular Manhattan plot of five meta-analyzed PUFA and MUFA traits. Plots show the -log_10_*P* of meta-analyzed GWAS for polyunsaturated fatty acid (PUFA) and monounsaturated (MUFA) traits. Red triangles designate the lead variant of a novel locus associated with a trait in our analysis. Red dotted lines at *P* < 2.439×10^-8^ indicate the genome-wide significance threshold corrected for effective number of traits. Alternating color shades within each ring designate breaks between chromosomes. Genes corresponding to loci with *P* < 1e-20 are labeled. All *P*-values were constrained to an upper limit of 1e-100 for visualization. Rings from outer to inner: omega-3 fatty acids, docosahexaenoic acid, omega-6 fatty acids, linoleic acid, and monounsaturated fatty acids.

Our literature review found a total of 210 previously reported significant PUFA and MUFA GWAS associations (106 unique variants, 37 loci based on 1Mb grouping; Supplementary Table 1). We compared these known loci with our significant meta-analysis loci. Of our 254 meta-analysis loci-trait associations, 173 were novel (87 unique across traits), and 102 of our novel loci were replicated in at least one external EUR study. This yielded a total of 51 unique loci across traits (Supplementary Table 13).

### Notable associated genes

Among the 109 unique genomic loci identified in our meta-analysis of five PUFA and MUFA traits, thirteen loci were associated with all five traits (Figure 2; Supplementary Table 15), nine of which were identified in previous GWAS. These loci spanned genes that are well-known in lipid metabolism, including the apolipoprotein gene clusters at chr11q23 (*APOA5, APOA4, APOC3*, and *APOA1*) and chr19q13 (*APOE, APOC1, APOC4*, and *APOC2*), plus *APOB*, LDL receptor adaptor protein 1 (*LDLRAP1)*, LDL receptor (*LDLR*), lipase C (*LIPC*), and lysophosphatidic acid receptor 2 (*LPAR2*). Another notable known gene is glucokinase regulator (*GCKR*), which has been associated with docosapentaenoic acid (DPA) and palmitoleic acid ^18-20^. Of the four novel loci associated with all five traits, the locus of chr18q21 covers a candidate gene of lipase G (*LIPG*). The candidate genes at loci chr1p13 and chr8q24 include *PSRC1, SORT1, TRIB1*, and *SQLE*. Both *PSRC1* and *TRIB1* have been previously associated with familial hypercholesterolemia ^21^. *SQLE* encodes squalene epoxidase, a rate-limiting enzyme catalyzing the first oxygenation step in sterol biosynthesis.

When excluding MUFAs and only considering n-3, DHA, n-6, and LA, there are three loci associated with these four traits, all of which are novel and externally replicated. These are located at chr2q21 (the *LCT* locus), chr4q13, and chr7p22 (Supplementary Table 15). The chr4q13 locus (Figure 3A) encompasses multiple members of the UDP-glycosyltransferase family (*UGT2B17; UGT2B10; UGT2B11; UGT2A1; UGT2A2;* and *UGT2A1*), which play important roles in bile acid (BA) detoxification by catalyzing the glucuronidation of BA substrates, and impact dietary lipid absorption ^22^. One candidate gene at chr7p22 is *CYP2W1* (Figure 3B), a member of the cytochrome P450 superfamily, which encodes monooxygenases and oxidizes steroids, fatty acids, and xenobiotics ^23^.

**Figure 3.**
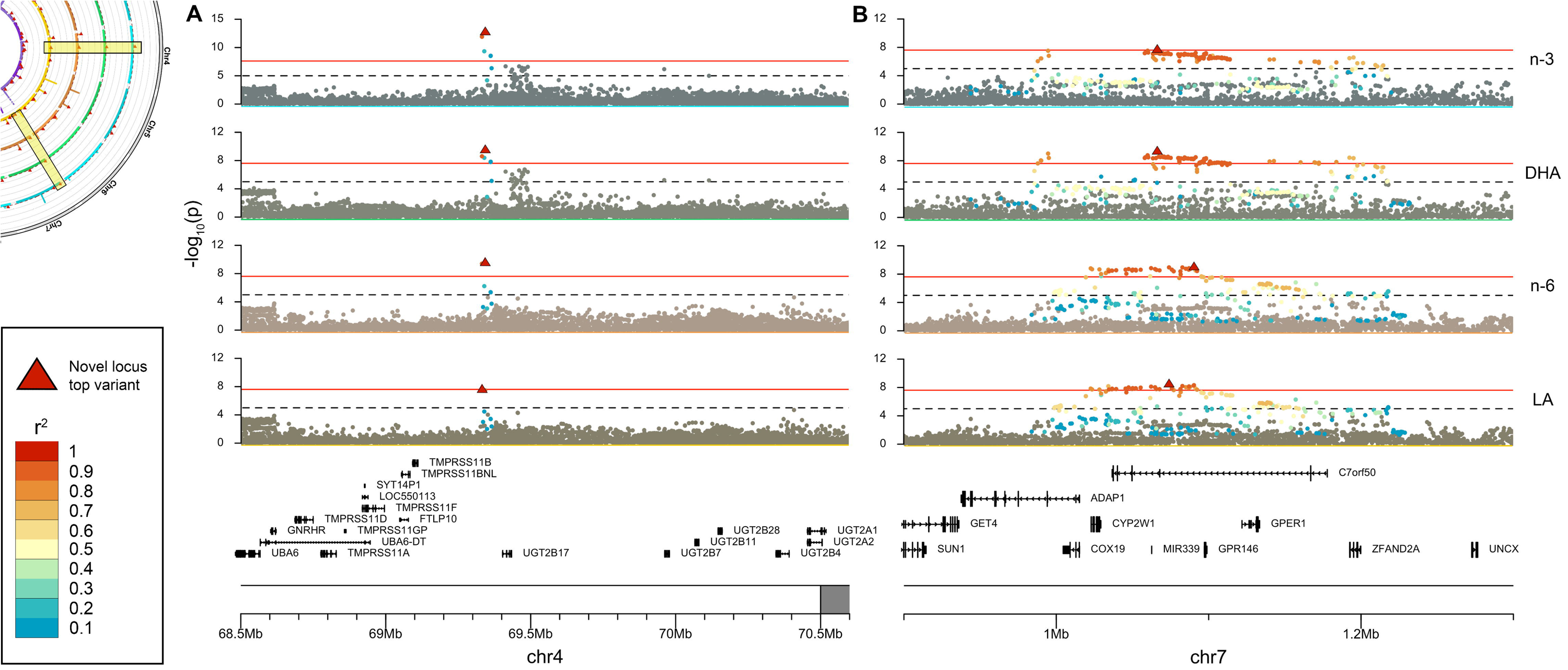
Regional Manhattan plots for selected novel loci. Local association plots of significant loci at (**A**) chr4q13 and (**B**) chr7p22. These loci have novel, replicated associations with all four meta-analyzed polyunsaturated fatty acid (PUFA) traits (from top to bottom: omega-3 fatty acids, docosahexaenoic acid, omega-6 fatty acids, linoleic acid). Genes in each region are shown below the Manhattan plots. Red triangles designate the lead variant of each locus-trait association. Variants in linkage disequilibrium with the lead variant are color-coded according to their r^2^ values.

Nine genomic loci are only associated with n-3, DHA, or both, eight of which are novel. The one known locus has at least two relevant candidate genes, choline kinase alpha (*CHKA*) and carnitine palmitoyltransferase 1A (*CPT1A*). *CHKA* encodes the initial enzyme which catalyzes the phosphorylation of ethanolamine in the CDP-choline pathway for phosphatidylcholine biosynthesis. *CPT1A* locates in the outer membrane of mitochondria and catalyzes the transport of long chain fatty acids from cytosol into mitochondria, enabling beta-oxidation. One novel locus at chr10q23 has a cluster of genes in the cytochrome P450 superfamily (*CYP2C18, CYP2C19, CYP2C9*, and *CYP2C8*). Another notable novel locus at chr11q24 has a candidate gene of *ST3GAL4*, which is involved in the terminal sialylation of glycolipids. There are 35 loci associated with only n-6, LA, or both; 33 of these are novel. Multiple novel candidate genes are implicated in lipid metabolism, such as LDL receptor related protein 2 (*LRP2*), NPC1 like intracellular cholesterol transporter 1 (*NPC1L1*), scavenger receptor class B member 1 (*SCARB1*), phospholipase C gamma 1 (*PLCG1*), and lipin 3 (*LPIN3*). Three additional novel loci carry cytochrome P450 genes, including chr2q33 (*CYP20A1*), chr8q12 (*CYP7A1)*, and chr19q13 (*CYP2A6*). Another two notable candidate genes are arachidonate 5-lipoxygenase (*ALOX5*) and peroxisome proliferator activated receptor delta (*PPARD*). ALOX5 catalyzes conversion of DHA to signaling molecules ^24^; supplemental DHA modulates PPARD in adult men ^25^.

The two key sets of genes which catalyze LCPUFA biosynthesis are fatty acid desaturase (*FADS*) and elongase protein family genes (*ELOVL*) ^7^. We have re-confirmed the primary importance of *FADS* genes in n-3 LCPUFA genetics, as these genes had the most significant *P*-values in our meta-analysis, at lead SNPs rs174528 (DHA, *P* < 1E-300; MAF = 0.39) and rs509360 (n-3, *P* < 1E-300; MAF = 0.33). Both of these variants were mapped to *FADS1, FADS2*, and *FADS3*. The *ELOVL* gene family has been rarely associated in GWAS; of the seven *ELOVL* genes, only *ELOVL2* (chr6:10,980,992-11,044,547) has been previously associated with PUFAs, specifically with the n-3 traits DHA, EPA, and DPA (Supplementary Table 1, locus 13). We found an association in UKB-EUR with the *ELOVL2* locus surpassing the suggestive significance threshold (*P* < 5e-05) for DHA (rs9380082, *P* = 2.6e-05), but did not find this locus associated at genome-wide significance. We did identify a novel, unreplicated association for MUFAs to total fatty acids percentage close by to *ELOVL6* (Supplementary Table 6). The variant rs114816312 (chr4:110,578,226; *P* = 1.9e-08) is ∼330Kbp downstream from *ELOVL6*. This tentative trait association would be consistent with previous findings that demonstrate the primary role of *ELOVL6* gene product in elongating MUFAs ^26^. This variant rs114816312 is also found within phospholipase A2 group XIIA (*PLA2G12A*); the primary function of PLA2 enzymes is to remove arachidonic acid (AA) from phospholipids for the production of eicosanoids.

ACSL6 is part of the Acyl-CoA synthetase (ACS) family of enzymes which catalyze the formation of acyl-CoAs from free fatty acids ^27^. We report two novel and externally replicated associations (Supplementary Table 15): n-3 with rs273913 (MAF = 0.39; locus start = chr5:131,407,493; Supplementary Figure 5), and DHA with rs166635 (MAF = 0.31; locus start = chr5:131,590,114), that are ∼60Kbp and ∼242Kbp upstream from the *ACSL6* gene (chr5:131,142,683-131,347,936; reverse strand). It should be noted with regard to novelty, this locus was previously reported to be associated with AA at rs274559 ^6^, but their *P*-value at 3.81e-06 did not reach genome-wide significance. *ACSL6* expression has been previously linked to DHA enrichment in the brain. Our finding of significant associations with only n-3 and DHA, not n-6, LA, or MUFAs, is consistent with previous experimental reports ^27,28^.

### Trait correlations and heritability

We evaluated the shared genetic basis across PUFA and MUFA traits using genetic correlations (*r*_*g*_). For the fourteen traits in the discovery analysis, the levels of phenotypic (*r*_*p*_) and genetic correlations were broadly consistent across all 91 trait-pairs, with slightly stronger genetic correlations (Figures 4A, 4B; Supplementary Table 7). Among the 78 trait-pairs that had both nominally significant phenotypic and genetic correlations (adj*P* < 0.05), 57 had stronger correlations at the genetic level (binomial test *p* = 2.79e-5). Interestingly, the genetic correlations between the absolute concentrations and their relative percentages of total fatty acids were not always high, ranging from 0.89 for MUFAs, 0.84 for omega-3, 0.73 for DHA, -0.56 for omega-6, -0.32 for PUFAs, to 0.017 for LA. These medium to low genetic correlations suggest the involvement of different biological mechanisms and emphasize the need to perform separate GWAS for absolute concentrations and relative percentages. Moreover, the correlation between n-3 and n-6 was moderate with the absolute concentrations (*r*_*g*_ = 0.67; *r*_*p*_ = 0.45), and low using their respective percentages of total fatty acids (*r*_*g*_ = -0.11; *r*_*p*_ = -0.12), indicating that there is substantial unique genetic basis for either trait.

**Figure 4.**
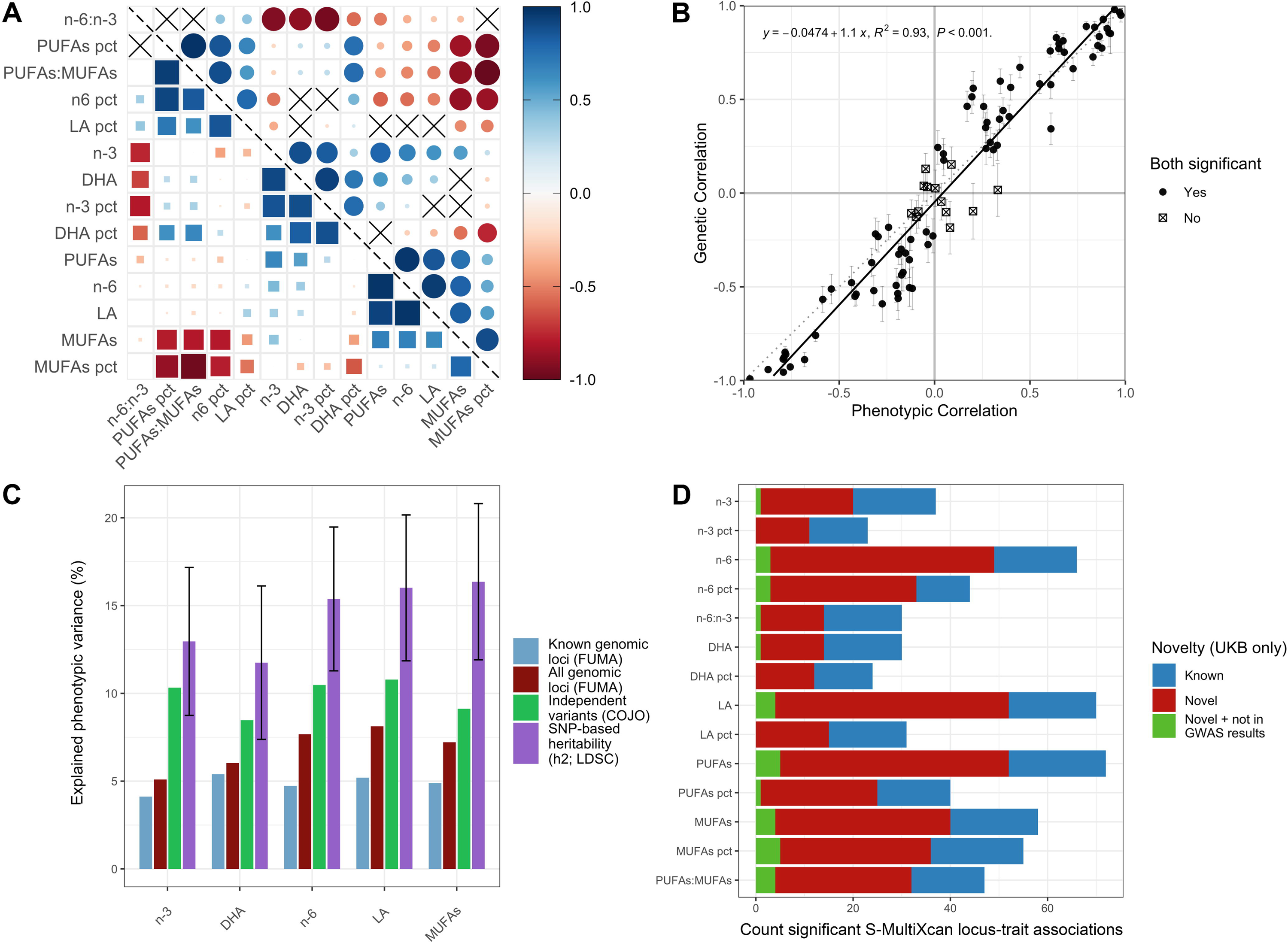
Results from genetic and phenotypic correlations, heritability, and S-MultiXcan. **(A)** Genetic and phenotypic correlations (*r*_*g*_ and *r*_*p*_) across all UK Biobank discovery cohort (UKB-EUR) polyunsaturated fatty acid (PUFA) and monounsaturated fatty acid (MUFA) traits. Above diagonal (circles) are genetic correlations; below diagonal (squares) are phenotypic correlations. Color and shape size both correspond to direction and strength of correlation. An “X” signifies a non-significant adjusted *P*-value for correlation coefficient. **(B)** Plot of genetic correlation vs. phenotypic correlation coefficients for 91 trait pairs. Error bars designate s.e.m. of correlation coefficient. Points with “X” did not reach significance for phenotypic correlation, genetic correlation, or both. **(C)** Explained phenotypic variance by different variant-grouping methods. SNP-based heritability shown with 95% confidence intervals. **(D)** Counts of significant locus-trait associations identified by S-MultiXcan for each trait in UKB-EUR, colored by novelty status.

In the fourteen traits tested in the discovery cohort, SNP-based heritability (h^2^) calculated in LDSC ranged from 0.12 (SE = 0.018) for LA percentage to 0.20 (0.032) for MUFA percentage (Supplementary Table 4). Using individual-level genotype data in the discovery cohort, BOLT-REML found the h^2^ of the six traits measured in absolute concentration units (n-3, n-6, DHA, LA, PUFAs, MUFAs) to range from 0.16 (0.0065) for LA to 0.22 (0.0066) for MUFAs (Supplementary Table 7). The lower range of LDSC when compared to BOLT-REML is consistent with our expectation that LDSC reports the lower bound of h^2^ estimates. SNP-based h^2^ was similar in meta-analysis, ranging from 0.12 (0.022) for DHA to 0.16 (0.023) for MUFAs (Figure 4C; Supplementary Table 13). Meta-analysis FUMA-defined significant loci explained between 5.09 to 8.12% of variance in the five traits examined; the novel loci we identified contributed between 0.63-2.95% of that variance. COJO independent variants explained between 8.47-10.78% of trait variance; the amount of SNP heritability explained ranged from 55.77% for MUFAs to 79.65% for n-3 (Fig 4C; Supplementary Table 13). Our independent association signals capture majority of the common variants underlying these five PUFA and MUFA traits.

### Transcriptome-wide association analysis

To identify genes with expression associated with PUFA and MUFA traits, S-PrediXcan was used to integrate GTEx (v8) eQTL (expression quantitative trait loci) data from 49 tissues and UKB-EUR cohort GWAS summary statistics. Across fourteen traits in the discovery stage, 24,666 Bonferroni-corrected significant gene-trait associations comprised of 527 unique genes were identified (Supplementary Table 8). We then used S-MultiXcan to find joint effects of gene expression associations across tissues. We found 2,818 associations (601 unique genes), of which 392 unique genes have not been found in previous GWAS for PUFA and MUFA traits (Supplementary Figure 6; Supplementary Tables 9, 10).

Since there was a high degree of overlap between TWAS and GWAS results, we searched for novel gene-trait associations in S-MultiXcan that had not been found in our discovery or meta-analysis GWAS analyses. We found 55 genes, spanning 12 loci, that were identified exclusively in TWAS and are novel though unreplicated (Figure 4D). Many of these genes (44) are found in a cluster at 6p21. These 44 genes are significantly enriched for “immune system process” (GO: GO:0002376; 17 genes; FDR = 7.41E-04) and “regulation of immune system process” (GO:0002682; 17 genes; FDR = 2.09E-06). Three other novel genes, *F2* (MUFAs), *WDR81* (LA), and *PTK2* (PUFAs), are involved in “regulation of lipid kinase activity” (GO:0043550).

### Gene set enrichment analysis

MAGMA tissue expression analysis for sets of positionally mapped genes from each of the five meta-analyzed PUFA and MUFA traits revealed that liver was exclusively the significantly enriched tissue type (Supplementary Figure 7). Because of this, we sent genes mapped from GTEx liver eQTLs and HiC liver chromatin data, in addition to positionally mapped genes, to GENE2FUNC for gene set enrichment (Supplementary Table 16).

Across the five traits, the most significant gene sets in the categories of Curated gene set, Positional gene set, Gene Ontology (GO): Biological process, GO: Cellular component, GO: Molecular function, Cancer modules, Canonical pathways, Computational gene sets, KEGG pathways, and TF targets, were all driven by genes in the major histocompatibility complex (MHC). However, the MHC region is the most polymorphic in the human genome, associated with the most disease traits, and determining causal variants in this region is highly prone to confounding ^29^, so we have excluded these from subsequent enrichment analyses. The next most significant positional gene set in all five PUFA and MUFA traits is chr6p22, corresponding to the *GCKR* locus; this locus has been identified in several previous GWAS (Supplementary Table 1). We found that significantly enriched gene sets in Wikipathways included statin pathway, histone modifications, pathways of LDL, HDL, and triglycerides, and metabolism of several nutrients, including zinc, copper, Vitamin B12, folate, and Vitamin A (Fig 5A). In the gene sets defined by GWAS catalog, the most significant enrichments are in genes that have previously been associated with blood lipids, including total cholesterol, LDL cholesterol, and triglycerides, as well as several traits related to mental characteristics such as “autism spectrum disorder or schizophrenia” (n-6, LA, and MUFAs only), “Bipolar disorder (I and II)” and “General factor of neuroticism” (Fig 5B).

**Figure 5.**
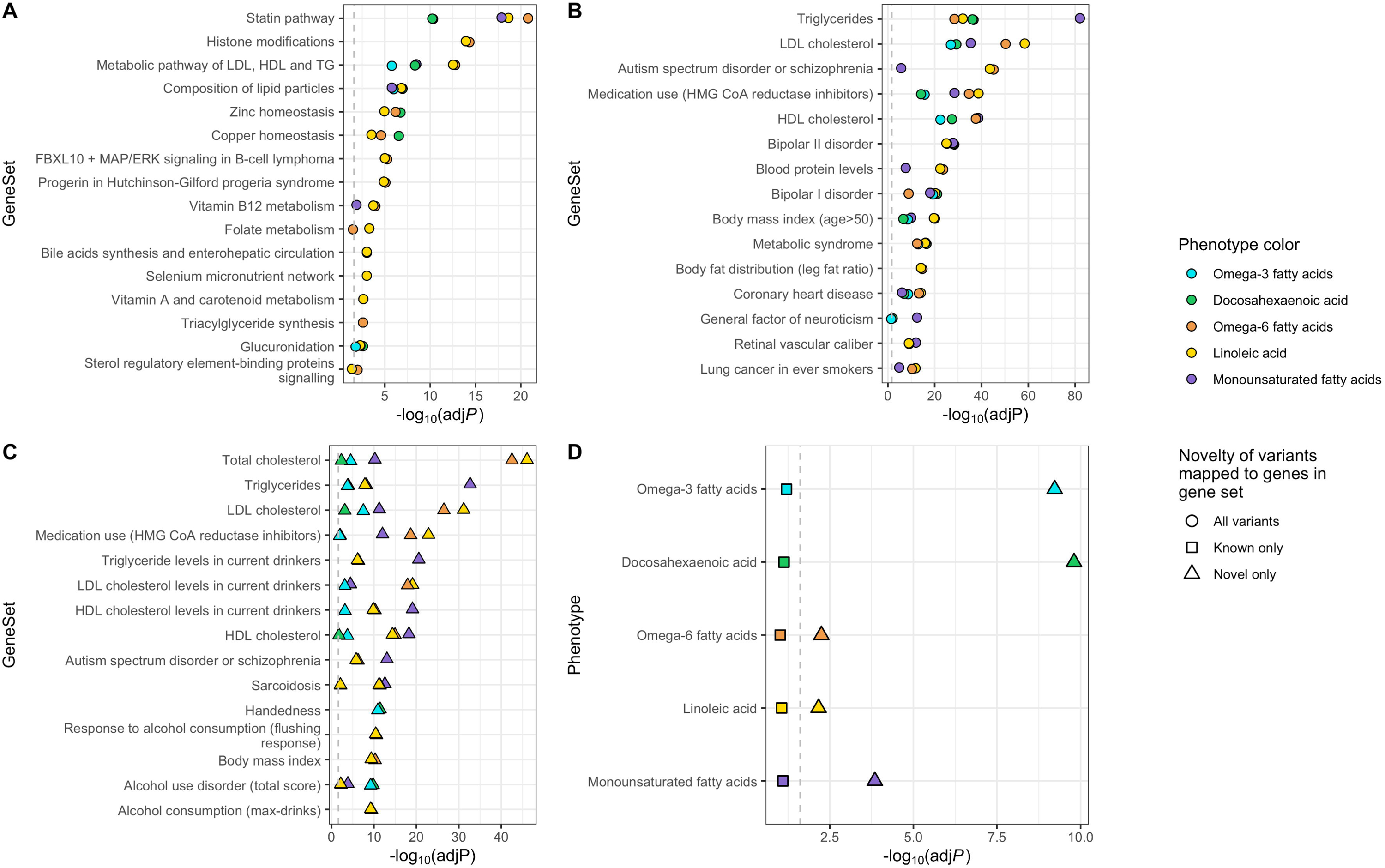
Gene sets mapped to significant meta-analysis loci. Enrichment -log_10_(adj*P*) for gene sets mapped to significant variants for five meta-analyzed polyunsaturated fatty acid (PUFA) and monounsaturated fatty acid (MUFA) traits. FDR adjusted *P*-values shown by trait for: **(A)** Wikipathways **(B)** GWAS Catalog **(C)** GWAS Catalog (novel variants only) **(D)** Variants mapped to the GWAS catalog trait “alcohol use disorder (total score),” stratified by novelty.

To identify new relationships between novel genes associated with our meta-analyzed traits and previously reported traits in GWAS Catalog, we stratified GENE2FUNC analysis based on novelty (Figure 5C; Supplementary Table 17). The second most significant gene set enrichment of non-lipid GWAS catalog traits for n-3 and DHA (after “Handedness”) was “Alcohol use disorder (total score)” (n-3: adj*P*_novel_ = 5.95E-10, adj*P*_known_ = 0.0052, adj*P*_all_ = 3.38E-09; DHA: adj*P*_novel_ = 1.57E-10, adj*P*_known_=0.075, adj*P*_all_ = 1.73E-10; Figure 5D). Additionally, many alcoholism-adjacent traits were found to be significantly enriched for n-6, LA, and MUFAs, such as “Triglyceride/LDL/HDL levels in current drinkers”, “Response to alcohol consumption (flushing response”, and “Alcohol consumption (max-drinks).” There was a total of 72 genes mapped to novel PUFA and MUFA variants that were significantly enriched for fifteen GWAS Catalog alcohol-related traits. These were located at 46 loci across 19 chromosomes, indicating the enrichment signal was not driven by a few gene clusters. Alcohol-related traits have been experimentally linked to PUFAs in multiple studies ^30-34^ (more in discussion). Therefore, our gene set enrichment analysis highlights a possible role of PUFAs and MUFAs in mental health, especially related to alcohol usage.

## Discussion

Here we report the largest GWAS to-date of PUFA and MUFA phenotypes, performed in European ancestry cohorts (EUR; N_EUR_ = 124,024), consisting of fourteen traits in the discovery stage, five of which were replicated and meta-analyzed in external EUR cohorts. The discovery cohort, those designated EUR in UK Biobank (UKB-EUR), is the largest publicly available human dataset with measures of these traits in genotyped participants (N_UKB-EUR_=101,729 after QC). We have identified 51 novel and externally replicated loci, as well as 36 loci that were not replicated, but have not been reported in previous GWAS (Supplementary Tables 1, 15). Considering that only 37 genomic risk loci were previously reported in relation to these traits, this study greatly increases our scope of understanding the genetic architecture of PUFAs and MUFAs. Of the 37 previously reported loci, we have replicated 23 loci in our discovery analysis (UKB-EUR) and 22 loci in our EUR meta-analysis (Supplementary Table 1). We have included plausible biological mechanistic explanations for many of our novel loci, and we have also added context to previously identified GWAS loci, to provide the most comprehensive functional analysis of variants associated with PUFA and MUFA traits to-date.

In our follow-up analysis of genes mapped to loci from our meta-analysis, we found notable differences between novel and known gene enrichment *P*-values for the GWAS catalog trait “Alcohol use disorder (total score)” (AUD) (Supplementary Table 15, Figure 5D). Across the phenotypes n-3, n-6, DHA, and LA, there are two novel clusters of PUFA-gene associations that have previously been associated with AUD. These genes are *PLEKHM1, CRHR1, SPPL2C, MAPT, STH*, and *KANSL1, NSF*, and *WNT3* at chr17q21.31, mapped to n-3 and DHA; and *FUT2, MAMSTR, RASIP1*, and *IZUMO1* at chr19q13.33, mapped to n-6 and LA. The inversion at chr17q21.31 has recently been associated with alcohol intake in a GWAS of ∼127,000 European participants from the Million Veterans Program cohort ^30^. The association of the gene cluster at chr19q13.33 with AUD was reported as a novel association in an analysis of ∼435,000 European participants of UKB ^31^.

In addition to the shared genetic variants between PUFAs and AUD, such as variants in *SNX17* and *GCKR*, variability in PUFA levels has been associated directly with AUD. The direction of causality between these traits has not been clearly disentangled. DHA has a neuroprotective effect against binge alcohol drinking, and is depleted with alcohol exposure ^35^. In the opposite causal direction, high alcohol consumption was associated with lower fatty acid intake measured by 24 hour recall in the 2001-2002 National Health and Nutrition Examination Survey in 4,168 adults ^32^. Deficiencies in n-3s are associated with bipolar disorder ^36^, which can lead to higher cravings for alcohol. Additionally, alcohol abuse has been characterized by an increase in oleic acid / LA ratio; Teubert et al. demonstrated a shift back to higher LA during alcohol detoxification in a small study of 45 alcoholic patients ^34^. Overall, the data on this topic are sparse, and more research should be done to elucidate this relationship.

Our study has several limitations. First, the PUFA and MUFA traits that we were able to investigate are limited to those reported by the UKB NMR metabolomics panel. We cannot resolve, for instance, differences in specific PUFAs that are often reported with higher resolution metabolite analyses, such as the difference in effects associated with DHA and other n-3s, notably eicosapentaenoic acid (EPA). Second, UKB is known to have volunteer bias, which can skew results, as has previously been shown ^37^.

Next, our analysis is mostly limited to determining the genetic associations of PUFA and MUFA traits in EUR populations. We recognize that an overwhelming number of genomic analyses to date have been conducted on EUR populations ^38^, to the detriment of understanding other ancestry groups. Further, our replication analysis shows that of 115 discovery loci in UKB-EUR, only 47 were replicated at *P* < 0.05 in one or more of the AFR, CSA, or EAS multi-ancestry groups. While power calculation shows that the difference was mainly driven by small sample sizes of non-EUR samples, it may also be that there is a distinct set of variants associated with PUFA and MUFA traits in non-EUR groups. We hope that the results of this analysis can be meta-analyzed with ancestrally diverse participant groups in future studies.

Another limitation is possibly introduced by the quantification of PUFA and MUFA traits using nuclear magnetic resonance spectroscopy (NMR). NMR has advantages and disadvantages as compared to the gold standard methods for quantitative fatty acid analysis, specifically high-resolution capillary gas chromatography coupled to flame ionization detection (FID) or mass spectrometry (GC-MS), or alternatively, liquid chromatography mass spectrometry (LC-MS) ^39^. First, the speed and cost advantages of NMR over GC or MS are advantageous in biobank-scale sample quantification ^40^. NMR is also a non-destructive technique, meaning samples can be stored and re-measured in the future. However, NMR is of reduced sensitivity and selectivity compared to GC-based techniques. GC resolves all fatty acids at picogram levels, compared to NMR which operates at minimum on milligram scale ^41^. GC-MS is also able to perform fatty acid analysis of high selectivity and completely resolve analytes ^42^; NMR resolution is limited, and great care must be taken to ensure confounding overlapping signals are avoided, particularly in complex mixtures ^41^. Nevertheless, as discussed above, our most significant results are congruent with biochemical expectations and with previous GWAS studies, including those studies which used MS-based quantification (Supplementary Table 1). This adds confidence to our usage of NMR-measured phenotypes and strengthens our novel findings.

Finally, as with any GWAS, significant associations are no more than candidates for mechanistic processes that, when altered, will have a reproducible influence on traits and ultimately human health. Replication of the associations and detailed investigation in experimental models and in randomized control trials are required to lead to clinical and precision nutrition applications. This study adds to a growing body of genomics literature that may help realize these applications in relation to PUFA and MUFA traits ^43^.

## Methods

### Ethics

Participant data use was approved by UK Biobank (UKB; Project ID 48818). UKB participants have consented to the use of their medical and genetic data in research studies. This research was performed on a University of Georgia (UGA) computing cluster with strict data protection protocols and two-factor authentication. The UGA Institutional Review Board (IRB) approved the use of human subject data in this study. Additional datasets use publicly available summary statistics from previous GWAS, and approval was not required.

### Participants

The full UKB consists of > 500,000 volunteer participants between ages 40 and 70 that were recruited between 2006 and 2010 in England, Scotland, and Wales. Approximately 120,000 participants had metabolic traits measured from plasma samples taken at recruitment using NMR between June 2019 and April 2020. Participants used in this study were removed on the following criteria: withdrawn consent, mismatches between self-reported and genetic sex, poor quality genotyping as flagged by UKB, sex chromosome aneuploidy, or poor-quality NMR measurement flagged by UKB. After quality control (QC) and stratification by ancestry using Pan UKBB designations ^16^, counts of UKB participants included in our analyses were: 101,729 European (EUR); 1,564 African (AFR); 2,203 Central South Asian (CSA); and 633 East Asian (EAS). UKB participant characteristics can be found in Supplementary Table 2.

For replication and meta-analysis, we included two external EUR studies. First, the Finnish Metabolic Sequencing (FinMetSeq) study ^11^, which consists of a combination of FINRISK and METSIM cohorts. METSIM participants were 10,197 men from Kuopio, Eastern Finland, aged 45 to 73 years during initial examinations from 2005 to 2010. FINRISK participants were recruited every five years from 1972 to 2012, and consisted of random population samples of men and women aged 30-59 years. FinMetSeq used 10,192 participants from 1992-2007 FINRISK surveys who had a residence in northeastern Finland. Pregnant women, type 1 and 2 diabetics, and those fasting less than eight hours were excluded from this cohort. Of the approximately 19,000 participants in FinMetSeq, 8,751 had NMR metabolomics data available for the fatty acid phenotypes of interest and were used in this study.

We also utilized a meta-analysis consisting of fourteen genotyped datasets derived from ten EUR studies performed by Kettunnen et al. in 2016 ^14^ in our replication and meta-analysis. The number of participants contributed by the Kettunnen et al. summary statistics in this analysis ranges from 3,644-13,544 participants (from six to ten studies), depending on the variant; this participant range is consistent across the five traits meta-analyzed. There is an overlap of 225 participants between the FinMetSeq and Kettunnen et al. cohorts, which we determined would not affect type I error in a meaningful way ^44^.

### Fatty acid phenotypes

UKB EDTA (ethylenediaminetetraacetic acid) plasma samples were taken at the baseline recruitment timepoint and measured between June 2019 and April 2020 by the metabolic biomarker profiling platform of Nightingale Health Ltd., as described previously ^40^. We analyzed fourteen quantitative polyunsaturated fatty acid (PUFA) and monounsaturated fatty acid (MUFA) phenotypes in UKB cohorts, specifically, n-3 LCPUFAs, n-6 LCPUFAs, docosahexaenoic acid (DHA), linoleic acid (LA), total PUFAs, and total monounsaturated fatty acids (MUFAs), all reported in mmol/L, the percentages of n-3 LCPUFAs, n-6 LCPUFAs, DHA, LA, MUFAs, and PUFAs out of the total amount of fatty acids (designated “pct”), as well as n-6/n-3 ratio, and PUFA/MUFA ratio. We regressed all PUFA and MUFA traits on selected covariates in each model (described below) and applied rank-based inverse normal transformation (indirect INT) to the residuals for use in all analyses ^45^; this was consistent with transformations performed in our external GWAS replication studies.

### Genotype data

Genotype data was initially QCed and imputed with Haplotype Reference Consortium (HRC) and 1000 Genomes variants by UKB (v3) as previously described ^46^. For discovery analyses, we excluded variants with imputation quality (INFO) score < 0.3, minor allele frequency (MAF) < 0.1%, missing genotype per individual > 5%, missing genotype per variant > 5%, or Hardy-Weinberg equilibrium (HWE) *P* < 1×10^-8^. After quality control, a total of 15,587,898 variants among 101,729 participants were included in the UKB-EUR discovery cohort. QC and genotype file format conversions were performed using PLINK2 alpha-v2.3 ^47,48^. All genomic positions in this study refer to autosomal chromosomes in the Genome Reference Consortium Human Build 37 (GRCh37), also known as hg19.

### Generating a pruned variant set

A pruned set of variants were inputted as PLINK-format genotypes to BOLT-LMM for model-fitting in the UKB-EUR discovery GWAS ^49^. After filtering for only participants included in this analysis, the exclusion criteria for variants in the pruned set were INFO score < 0.8, MAF < 1%, missing genotype per variant > 1%, or HWE *P* < 1×10^-8^. A hard-call threshold of 0.1 was applied to the filtered variants. The lactase locus on chromosome 2, the major histocompatibility complex (MHC) on chromosome 6, and inversions on chromosomes 8 and 17 were excluded. Linkage disequilibrium (LD) pruning was performed at r^2^ = 0.2, (plink2 --indep-pairwise 50 5 0.2). After pruning, 821,405 variants remained.

### Discovery model selection

Two sensitivity models were used in our initial discovery stage analysis. Model 1 (M1) included the covariates sex, age, age^2^, genotyping array, and assessment center. Model 2 (M2) included the M1 covariates, plus body mass index (BMI), lipid medication usage, and socioeconomic status as measured by Townsend deprivation index (Supplementary Table 3). The first twenty principal components for study participants as calculated by PLINK2 (randomized algorithm) ^47,48^ were also included as covariates in both models. We compared our summary statistics from M1 and M2 using Spearman’s rank correlation (two-sided test).

### Identification of significant GWAS signals

BOLT-LMM v2.3.6 ^49^ was used to perform linear mixed-effects model association analyses on fourteen PUFA and MUFA traits in the UKB-EUR discovery stage analysis. The provided 1000G European LD scores ^50^ were used to calibrate the BOLT-LMM statistic. Covariates and pruned variant sets were included in models as described above. Non-infinitesimal BOLT-LMM *P*-values (“P_BOLT_LMM”) were used in all reporting and downstream analyses. Because the fourteen PUFA and MUFA traits were highly related, we calculated the effective number of traits to use for Bonferroni multiple testing correction. Eigenvalues (*λ*) for the fourteen traits were used to calculate the number of effective traits as: 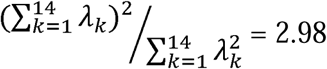^51^. The threshold of *P* < (5 × 10^-8^ / 2.98) = 1.678 × 10^-8^ was used to designate significant variant-trait associations in the discovery stage.

### Transcriptome-wide association analysis

Transcriptome-wide association analysis (TWAS) was performed on UKB-EUR discovery stage summary statistics using S-PrediXcan ^52^. This method used Genotype-Tissue Expression (GTEx) v8 ^53^ expression quantitative trait loci (eQTL) data across 49 available tissues. Summary statistics were harmonized and imputed to GTEx models. A total of 601,176 gene-tissue pairs were analyzed across fourteen PUFA and MUFA traits, using a Bonferroni corrected significance threshold of *P*□< (0.05 / (601,176 × 2.98)) = 2.791 × 10^-8^. S-MultiXcan was used to integrate tissue-level associations and increase association detection. The cutoff condition number of eigenvalues was set to 30 for truncating singular-value decomposition components. S-MultiXcan was run across 21,846 genes, using a Bonferroni corrected significance threshold of *P* < (0.05 / (21,846 × 2.98)) = 7.68 × 10^-7^.

### Replication and meta-analysis

Variant-trait associations from the discovery stage for five traits were sent to replication and meta-analysis steps: omega-3 fatty acids (n-3 LCPUFAs), omega-6 fatty acids (n-6 LCPUFAs), docosahexaenoic acid (DHA), linoleic acid (LA), and total monounsaturated fatty acids (MUFAs). Summary statistics were obtained from the publicly available Finnish Metabolic Sequencing (FinMetSeq) study ^11^ and the meta-analysis of 10 studies by Kettunen et al. ^14^. FinMetSeq summary statistics as provided had been adjusted by age, age^2^, sex, cohort year, BMI, sex hormones, and lipid medications. Kettunnen et al. had adjusted for sex, age and ten genetic principal components in their analyses. Discovery stage variants were considered replicated in the external cohorts on a per-variant basis at *P* < 0.05.

Meta-analyses were performed using the METAL ^54^ software using the STDERR scheme, which weights effect size estimates using the inverse of the corresponding standard errors. The meta-analysis of this study consists of the three EUR participant studies: UKB-EUR+ FinMetSeq+ Kettunen et al. datasets (N=114,124 to 124,024). MungeSumstats ^55^ was used in pre-processing to harmonize effect alleles from separate cohorts to the reference genome. UKB-EUR was used to estimate the number of effective traits for the three EUR meta-analysis cohorts. Number of effective traits was calculated from phenotype eigenvalues as 2.05 using the formula shown above, and the multiple-testing corrected threshold of *P* < (5 × 10^-8^ / 2.05) = 2.439 × 10^-8^ was used to designate significant meta-analysis variant-trait associations.

### Heritability and LD score regression

Restricted maximum likelihood (REML) estimates for genetic correlation and multi-trait heritability (h^2^) were calculated using BOLT-REML ^49^. The six phenotypes measured in absolute concentration units (mmol/L) from the discovery UKB-EUR cohort (n-3, n-6, DHA, LA, PUFAs, MUFAs) were inputted with the covariates age, age^2^, sex, assessment center, genotype batch, and the first twenty PCs. The provided 1000G European LD scores ^50^ were used to calibrate the BOLT statistic. The refinement step was skipped (--remlNoRefine) to increase computational efficiency.

LD score regression (LDSC) ^56^ was used to calculate LD Score regression intercept, genomic control (λ), and non-partitioned SNP-based h^2^. The 1000 Genomes European set was used as the LD reference panel ^50^. MungeSumstats ^55^ was used to harmonize alleles and convert summary statistics to LDSC format for this and subsequent steps. Variance explained was calculated by the formula (2 × MAF × (1 - MAF) × β^2^). We calculated variance explained for variants identified as independent significant associations, as well as for lead variants of genomic risk loci. Pairwise genetic correlations (*r*_*g*_) were computed from munged summary statistics using LDSC ^57^. Pairwise phenotypic correlations (*r*_*p*_) were calculated as Pearson’s correlation coefficient (two-sided). *P*-values for *r*_*g*_ and *r*_*p*_ were adjusted for false discovery rate (FDR).

### Identifying genetic loci

Lead variants for each independent genomic risk loci were defined in both the discovery (UKB-EUR) and meta-analysis cohorts (UKB-EUR + Kettunen et al. + FinMetSeq) by inputting summary statistics to the Functional Mapping and Annotation of Genome-Wide Association Studies (FUMA) web server ^58^. The UKB release2b 10k European set was used as the LD reference panel. The maximum *P*-value cutoff was set to 0.05, and a first LD threshold of *r*^2^ ≥ 0.6 and second threshold of *r*^2^ ≥ 0.1 were used to define loci and lead SNPs. SNPs not available in the GWAS input but contained in the reference panel were included in output. The maximum distance between LD blocks to merge into a locus was 250 Kb. Summary statistics *P*-values were set with a lower cap of *P* = 1e-300 to resolve FUMA processing errors that we identified related to minimum Python float size limit. Variants from meta-analysis were annotated to genes with SNP2GENE using positional mapping (maximum distance 10 Kb), eQTL mapping from GTEx v8 liver tissue, and chromatin interaction mapping using built-in data from Hi-C (GSE87112) liver tissue. All other FUMA mapping settings were kept as default.

### Identifying novel loci

A table of previously reported (“known”) PUFA- and MUFA-associated lead variants was prepared from full summary statistics (where available) or from significance tables found within previous GWAS publications (Supplementary Table 1). All reported genomic coordinates were set to hg19 using liftOver ^59^. Genomic risk loci coordinates were identified in each study by *P*-values of reported variants using FUMA (setting LD reference by ancestry of study, otherwise default settings). These loci were grouped together within a ± 500 Kb window prior to checking for novelty of our results, regardless of the ancestry of the study cohorts. Trait names were harmonized across studies. LDtrait ^60^ was used to cross-check our novelty table; no additional loci were found using this method.

### Multi-ancestry replication in non-EUR UKB Cohorts

Genome-wide Complex Trait Analysis mixed linear model-based association analysis (GCTA-MLMA) ^61^ was used to perform mixed-model GWAS analyses in the UKB African (AFR), Central/South Asian (CSA), and East Asian (EAS) cohorts (Supplementary Tables 2, 12). A genetic relatedness matrix (GRM) was generated for each population using GCTA-GRM ^62^. Covariates used in these models were age, age^2^, sex, and the first ten principal components. Genotype quality filtering parameters were INFO < 0.3, MAF < 1 %, missing genotype per individual > 5 %, missing genotype per variant > 5%, or HWE P < 1×10^-8^.

### Gene sets and pathway analysis

FUMA GENE2FUNC ^58^ was performed on genes mapped from SNP2GENE using parameters described above including all background gene-sets in hypergeometric tests, and using expression data from all GTEx v8 datasets. Benjamini-Hochberg (FDR) was used as the gene set enrichment multiple test correction method. Gene Ontology (GO) was used to categorize sets of genes in downstream analyses ^63^.

### Conditional and joint association analysis

Genome-wide Complex Trait Analysis Conditional and Joint Association Analysis (GCTA-COJO) with stepwise model selection to identify conditionally independent variants was performed using discovery and meta-analysis summary statistics (--cojo-slct) ^64^. A random set of 20,000 unrelated UKB-EUR participants were used as the LD reference (--bfile input). Variants with MAF < 1% were removed. COJO was run per chromosome with significance thresholds based on effective trait Bonferroni corrections (described above), using default settings for collinearity and window size. Summary statistics standard error (SE) values were re-calculated with higher precision based on effect size and *P*-values prior to input to COJO, to ensure GCTA-COJO output columns matched input.

### Visualizing results

CMplot ^65^ was used to generate the circular Manhattan plot in Figure 2. The regional Manhattan plots in Figure 3 were generated using karyoploteR ^66^. The correlation plot in Figure 4A was generated using ggcorrplot2 ^67^. The qqman R package ^68^ was used to generate Manhattan and QQ plots in Supplementary Figures 3 and 4. S-MultiXcan gene-based Manhattan plots in Supplementary Figure 6 were generated using the Manhattan R package ^69^. Bar plots and scatterplots were generated using ggplot2 ^70^ in R v4.1.0. Color palettes in all figures were optimized for accessibility with three major types of color blindness (deuteranopia, protanopia, and tritanopia) using https://color.adobe.com/create/color-accessibility.

## Data Availability

Summary statistics are available on GWAS Catalog. All relevant data are included within the manuscript. Additional intermediate data or scripts are available upon request to the authors.

https://github.com/michaelofrancis/PUFA_GWAS

## Acknowledgements

Research reported in this publication was supported by the National Institute of General Medical Sciences of the National Institute of Health under award numbers T32GM007103 (MF) and R35GM143060 (KY). The content is solely the responsibility of the authors and does not necessarily represent the official views of the National Institutes of Health.

Special thanks to the Georgia Advanced Computing Resource Center (GACRC) at the University of Georgia for supporting our data analyses.

## Author contributions

KY conceived and supervised the study. KY and MF designed the analyses. MF led the data analyses, with assistance from YS and HX. MF, KY, and JTB interpreted the results. MF and KY wrote the manuscript. MF created data visualizations. All authors reviewed, revised, and approved the final paper.

## Data availability

Full summary statistics can be found on GWAS Catalog, using the accession codes provided in Supplementary Table 18.

## Code availability

Script repository for this analysis can be found at https://github.com/michaelofrancis/PUFA_GWAS.

## Supplementary Figures

**S1. Participant characteristics for PUFA and MUFA traits of four ancestries in UK Biobank**. (**A**) Mean and s.d. of polyunsaturated fatty acid (PUFA) and monounsaturated fatty acid (MUFA) traits that were measured in absolute concentration units (mmol/L). (**B**) Left: mean percentage and s.d. of each trait measured in percentage of total amount of fatty acids. Right: Mean of ratio-based traits and s.d. of these values.

**S2. Correlation plot comparing *P*-values of our Models 2 versus Model 1**. GWAS -log_10_(*P*) between two sensitivity models were compared. Each plot is one of fourteen traits analyzed in the UK Biobank European discovery stage. Each point represents one variant. Spearman’s Rho (R) and correlation *P*-value shown.

**S3. Manhattan and QQ plots of UK Biobank discovery (EUR) dataset**. Left: Manhattan plots showing the -log_10_(*P*) of associations in each of fourteen polyunsaturated fatty acid (PUFA) and monounsaturated fatty acid (MUFA) traits in the UK Biobank European discovery cohort. Alternating point color shades indicate associations across 22 chromosomes. Red line at *P*=1.678×10^-8^ indicates genome-wide significance threshold corrected for number of effective traits. Right: Quantile-quantile (QQ) plots showing observed versus expected distributions of association *P*-values for each trait. Genomic control (λ) and Linkage Disequilibrium Score Regression intercept are shown. N_UKB-EUR_ = 101,729.

**S4. Manhattan and QQ plots of three UK Biobank multi-ancestry cohorts**. Manhattan plots showing the -log_10_(*P*) across 22 chromosomes for associations in each of fourteen polyunsaturated fatty acid (PUFA) and monounsaturated fatty acid (MUFA) traits in three UK Biobank multi-ancestry cohorts: African (AFR); Central and South Asian (CSA); and East Asian (EAS). Red line at *P* < 1.678×10^-8^ shows genome-wide significance threshold corrected for number of effective traits. Right: Quantile-quantile (QQ) plots showing observed versus expected distributions of association *P*-values for each trait. N_UKB-AFR_ = 1,564; N_UKB-CSA_ = 2,203; NUKB-EAS = 633.

**S5. ACLS6 novel association with omega-3 fatty acids**. Regional Manhattan plot showing local associations between genes in close proximity to *ACLS6* and omega-3 fatty acids in the meta-analysis GWAS of three European cohorts.

**S6. S-MultiXcan gene-based Manhattan plots**. Manhattan plots showing -log_10_(*P*) of significant associations between gene expression levels and summary statistics from fourteen polyunsaturated fatty acid (PUFA) and monounsaturated fatty acid (MUFA) traits in the UK Biobank discovery cohort. The red line indicates the corrected significance threshold at *P* < 7.68 ×10^-7^. The most significant genes for each 5Mb window of significant associations are labeled. Alternating color shades designate breaks between chromosomes.

**S7. Tissue expression analysis for meta-analyzed traits**. Significant tissue-expression specificity by tissue type for five meta-analyzed polyunsaturated fatty acid (PUFA) and monounsaturated fatty acid (MUFA) traits. Plots were created by MAGMA. Liver is the only significant tissue type identified in these traits.

## Supplementary Tables

**S1. Known PUFA loci**. Previously reported lead variants from significant genome-wide association studies of polyunsaturated fatty acids (PUFA) and monounsaturated fatty acids (MUFA). Each row represents a genomic risk locus identified by inputting summary statistics from previous publications into FUMA SNP2GENE. Phenotype abbreviations: AA: arachidonic acid; AdrA: adrenic acid; ALA: alpha-linolenic acid; DGLA: dihomo-gamma-linolenic acid; DHA: docosahexaenoic acid; DPA: *cis*-7,10,13,16,19-docosapentaenoic acid; DPAn6: *cis*-4,7,10,13,16-docosapentaenoic acid; EDA: eicosadienoic acid; EPA: eicosapentaenoic acid; FAw3: omega-3 fatty acids; FAw6: omega-6 fatty acids; FAw67: omega-6 and -7 fatty acids; GLA: gamma-linolenic acid; LA: linoleic acid; MUFA: monounsaturated fatty acids; OA: oleic acid; otPUFA: polyunsaturated fatty acids (other than 18:2); POA: palmitoleic acid; PUFA: polyunsaturated fatty acids.

**S2. Participant characteristics table for UK Biobank cohorts**. Phenotype and covariate data for UK Biobank cohorts. Continuous variables are represented as: mean (standard deviation). BMI: body mass index. EUR: European; AFR: African; CSA: Central and South Asian; EAS: East Asian.

**S3. Comparison of discovery models one and two**. Counts of genome-wide significant variants and compare Spearman correlation coefficient between *P*-values for variants in UKB-EUR discovery analysis sensitivity Models 1 and 2. See Supplementary Figure 2 for plots. Number of variants in analysis, genomic control (λ) and Linkage Disequilibrium Score Regression (LDSC) intercepts are shown for each trait of each model.

**S4. Discovery stage GWAS summary**. Number of significant variants, independent significant variants (from COJO), and significant loci for each of fourteen polyunsaturated fatty acid (PUFA) and monounsaturated fatty acid (MUFA) traits tested in the UK Biobank European discovery cohort. Novel loci for each trait and unique novel loci across are shown. SNP-based heritability (h^2^) and standard error (SE) are reported.

**S5. Discovery GCTA-COJO results**. Output from Genome-wide Complex Trait Analysis Conditional and Joint Analysis (GCTA-COJO) using summary statistics from fourteen polyunsaturated fatty acid (PUFA) and monounsaturated fatty acid (MUFA) traits in the the UK Biobank European discovery cohort. RefA: effect allele; freq: frequency of the effect allele in the original data; b: effect size; se: standard error; p: *p*-value from original GWAS; n: estimated effective sample size; freq_geno: frequency of the effect allele in the reference sample; bJ: effect size from joint analysis of selected SNPs; bJ_se: standard error from joint analysis of selected SNPs; pJ: *p*-value from joint analysis of selected SNPs; LD_r: LD correlation between the SNP i and SNP i + 1 for the SNPs on the list.

**S6. Discovery cohort significant loci**. Functional Mapping and Annotation of Genome-Wide Association Studies (FUMA) genomic risk loci from the UK Biobank (UKB) European discovery cohort summary statistics. Corresponding summary statistics from UKB multi-ancestry African (AFR), Central and South Asian (CSA), and East Asian (EAS) cohorts are also provided.

**S7. Genetic and phenotypic correlation matrices**. Top: Coefficients, standard error, and FDR adjusted *P*-values for genotypic and phenotypic correlations of fourteen polyunsaturated fatty acid (PUFA) and monounsaturated fatty acid (MUFA) traits in the UK Biobank European discovery cohort (UKB-EUR). Above diagonal are genetic correlations calculated using LDSC using UKB GWAS summary statistics. Below diagonal are phenotypic correlations by Pearson correlation coefficient. Bottom: Output table from BOLT-REML multi-trait heritability correlations for the six traits in the discovery UKB-EUR cohort measured in absolute concentration units (mmol/L). The diagonal represents heritability explained by genotyped SNPs, other values are genetic correlations between traits.

**S8. S-PrediXcan results**. Gene-trait associations by tissue type from S-PrediXcan, which reached the Bonferroni corrected significance threshold *P*□<□ 2.791×10^-8^ (0.05/(601,176×2.98). Gene: gene ID; gene_name: HUGO gene name; Zscore: S-PrediXcan association result for the gene; Pvalue: P-value of Zscore; var_g: variance of the gene expression, calculated as W’ × G × W (where W is the vector of SNP weights in a gene’s model, W’ is its transpose, and G is the covariance matrix); n_snps_used: number of SNPs in the covariance matrix; n_snps_in_model: number of SNPs in the model.

**S9. S-MultiXcan results**. Significant genes across tissue types from S-MultiXcan, which reached the Bonferroni corrected significance threshold of *P* < 7.68 ×10^-7^ (0.05/(21,846×2.98). Gene: gene ID; gene_name: HUGO gene name; pvalue: significance p-value of S-MultiXcan association; n: number of “tissues” available for this gene; n_indep: number of independent components of variation kept among the tissues’ predictions. (Synthetic independent tissues); p_i_best: best p-value of single-tissue S-PrediXcan association; t_i_best: name of best single-tissue S-PrediXcan association; p_i_worst: worst p-value of single-tissue S-PrediXcan association; t_i_worst: name of worst single-tissue S-PrediXcan association.

**S10. Summarize S-MultiXcan results**. Number of significant associations from S-PrediXcan and S-MultiXcan results for fourteen polyunsaturated fatty acid (PUFA) and monounsaturated fatty acid (MUFA) traits in the UK Biobank European discovery cohort. Novelty of results also shown.

**S11. UKB-EUR external replication**. Number of variants in common between UK Biobank European discovery cohort (UKB-EUR) and the external European cohorts FinMetSeq and Kettunnen et al., after munging. Counts of variants from UKB-EUR that were replicated at *P* < 0.05.

**S12. UKB multi-ancestry replication**. Number of variants in common between the UK Biobank European discovery cohort (UKB-EUR) and UKB multi-ancestry African (AFR), Central and South Asian (CSA), and East Asian (EAS) cohorts after quality control protocol. Counts of variants from UKB-EUR that were replicated at *P* < 0.05.

**S13. Meta-analysis stage GWAS summary**. Number of significant variants, independent significant variants, and significant loci for each of five meta-analyzed polyunsaturated fatty acid (PUFA) and monounsaturated fatty acid (MUFA) traits. Novel loci for each trait and unique novel loci are counted here. Linkage disequilibrium score regression (LDSC) intercept, genomic control, variance explained (%) and heritability (h^2^) are shown.

**S14. Meta-analysis GCTA-COJO results**. Genome-wide Complex Trait Analysis Conditional and Joint Analysis (GCTA-COJO) output using summary statistics from five meta-analyzed polyunsaturated fatty acid (PUFA) and monounsaturated fatty acid (MUFA) traits. RefA: effect allele; freq: frequency of the effect allele in the original data; b: effect size; se: standard error; p: *p*-value from original GWAS; n: estimated effective sample size; freq_geno: frequency of the effect allele in the reference sample; bJ: effect size from joint analysis of selected SNPs; bJ_se: standard error from joint analysis of selected SNPs; pJ: *p*-value from joint analysis of selected SNPs; LD_r: LD correlation between the SNP i and SNP i + 1 for the SNPs on the list.

**S15. Meta-analysis significant loci**. Functional Mapping and Annotation of Genome-Wide Association Studies (FUMA) genomic risk loci from meta-analysis of UK Biobank European discovery cohort, FinMetSeq, and Kettunen et al. studies. Corresponding summary statistics from each study are shown.

**S16. Gene set enrichment**. GWAS Catalog gene set enrichment of significant variant associations for five meta-analyzed polyunsaturated fatty acid (PUFA) and monounsaturated fatty acid (MUFA) traits. Genes in query gene sets mapped from significant meta-analysis associations by position, GTEx liver eQTLs, and HiC liver chromatin data using GENE2FUNC implemented by Functional Mapping and Annotation of Genome-Wide Association Studies (FUMA).

**S17. Gene set enrichment by novelty**. GWAS Catalog gene set enrichment, stratified by novelty of significant variant associations for five meta-analyzed polyunsaturated fatty acid (PUFA) and monounsaturated fatty acid (MUFA) traits. Genes in query gene sets mapped from significant meta-analysis associations by position, GTEx liver eQTLs, and HiC liver chromatin data using GENE2FUNC implemented by Functional Mapping and Annotation of Genome-Wide Association Studies (FUMA).

**S18. GWAS Catalog Accessions**. GWAS Catalog accession codes for all summary statistics generated in this study.

