## Supplementary Figures S1-S7 for "Fifty-one novel and replicated GWAS loci for polyunsaturated and monounsaturated fatty acids in 124,024 Europeans"

### Supplementary figures for the manuscript:
### Fifty-one novel, replicated loci identified in genome-wide association study of polyunsaturated and monounsaturated fatty acids in 124,024 European individuals
#### Francis et al. (2022)

| | PAGE |
|---|---|
| **S1. Participant characteristics for PUFA and MUFA traits of four ancestries in UK Biobank.** (**A**) Mean and standard deviation (SD) of polyunsaturated fatty acid (PUFA) and monounsaturated fatty acid (MUFA) traits that were measured in absolute concentration units (mmol/L). (**B**) Left: mean percentage and SD of each trait measured in percentage of total amount of fatty acids. Right: Mean of ratio-based traits and SD of these values. | 2 |
| **S2. Correlation plot comparing *P*-values of our Models 2 versus Model 1.** GWAS $-\log_{10}(P)$ between two sensitivity models were compared. Each plot is one of fourteen traits analyzed in the UK Biobank European discovery stage. Each point represents one variant. Spearman's Rho (R) and correlation *P*-value shown. | 3 |
| **S3. Manhattan and QQ plots of UK Biobank discovery (EUR) dataset.** Left: Manhattan plots showing the $-\log_{10}(P)$ of associations in each of fourteen polyunsaturated fatty acid (PUFA) and monounsaturated fatty acid (MUFA) traits in the UK Biobank European discovery cohort. Alternating point color shades indicate associations across 22 chromosomes. Red line at $P$=1.678*$10^{-8}$ indicates genome-wide significance threshold corrected for number of effective traits. Right: Quantile-quantile (QQ) plots showing observed versus expected distributions of association *P*-values for each trait. Genomic control (lambda) and linkage disequilibrium score regression intercept are shown. $N_{UKB-EUR}$ = 101,729. | 5 |
| **S4. Manhattan and QQ plots of three UK Biobank multi-ancestry cohorts.** Manhattan plots showing the $-\log_{10}(P)$ across 22 chromosomes for associations in each of fourteen polyunsaturated fatty acid (PUFA) and monounsaturated fatty acid (MUFA) traits in three UK Biobank multi-ancestry cohorts: African (AFR); Central and South Asian (CSA); and East Asian (EAS). Red line at $P$ < 1.678*$10^{-8}$ shows genome-wide significance threshold corrected for number of effective traits. Right: Quantile-quantile (QQ) plots showing observed versus expected distributions of association *P*-values for each trait. $N_{UKB-AFR}$ = 1,564; $N_{UKB-CSA}$ = 2,203; $N_{UKB-EAS}$ = 633. | 9 |
| **S5. ACLS6 novel association with omega-3 fatty acids.** Regional Manhattan plot showing local associations between genes in close proximity to *ACLS6* and omega-3 fatty acids in the meta-analysis GWAS of three European cohorts. | 20 |
| **S6. S-MultiXcan gene-based Manhattan plots.** Manhattan plots showing $-\log_{10}(P)$ of significant associations between gene expression levels and summary statistics from fourteen polyunsaturated fatty acid (PUFA) and monounsaturated fatty acid (MUFA) traits in the UK Biobank discovery cohort. The red line indicates the corrected significance threshold at $P$ < 7.68 *$10^{-7}$. The most significant genes for each 5Mb window of significant associations are labeled. Alternating color shades designate breaks between chromosomes. | 21 |
| **S7. Tissue expression analysis for meta-analyzed traits.** Significant tissue-expression specificity by tissue type for five meta-analyzed polyunsaturated fatty acid (PUFA) and monounsaturated fatty acid (MUFA) traits. Plots were created by MAGMA. Liver is the only significant tissue type identified in these traits. | 28 |

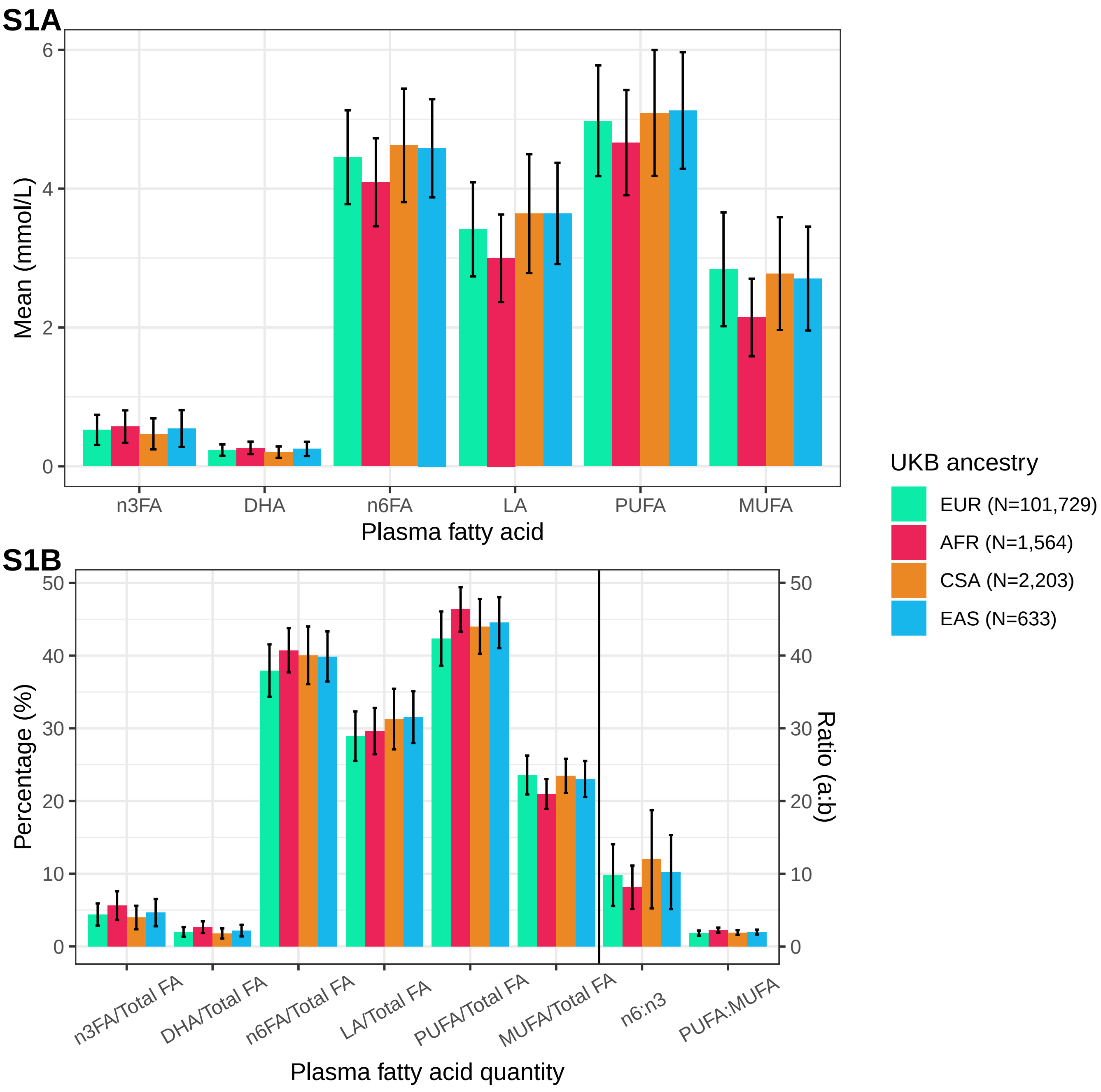
S1A
Mean (mmol/L)
0
2
4
6
n3FA
DHA
n6FA
LA
PUFA
MUFA
Plasma fatty acid
S1B
Percentage (%)
Ratio (a:b)
0
10
20
30
40
50
n3FA/Total FA
DHA/Total FA
n6FA/Total FA
LA/Total FA
PUFA/Total FA
MUFA/Total FA
n6:n3
PUFA:MUFA
Plasma fatty acid quantity
UKB ancestry
EUR (N=101,729)
AFR (N=1,564)
CSA (N=2,203)
EAS (N=633)

**S2**

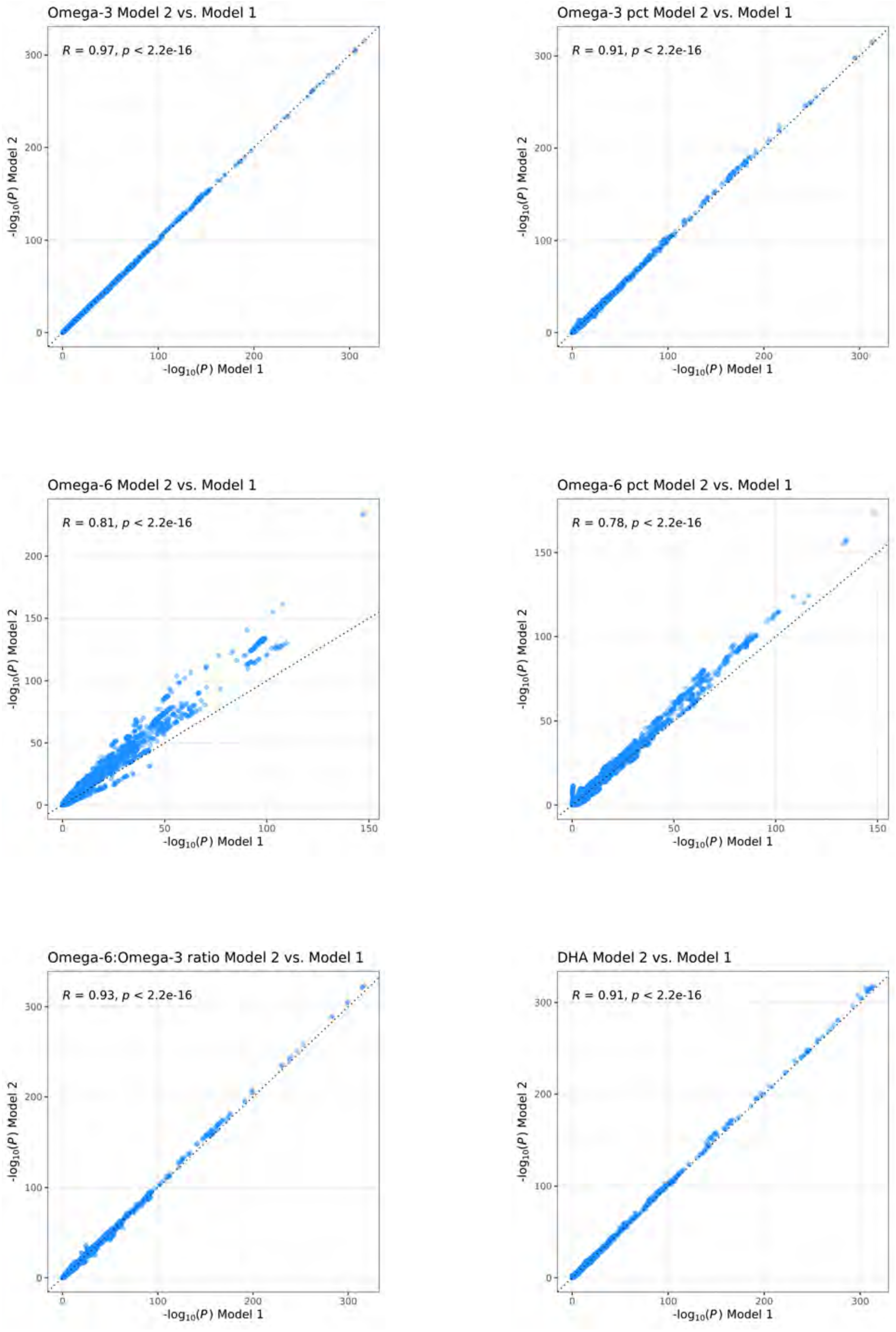
Omega-3 Model 2 vs. Model 1
R = 0.97, p < 2.2e-16
Omega-3 pct Model 2 vs. Model 1
R = 0.91, p < 2.2e-16
Omega-6 Model 2 vs. Model 1
R = 0.81, p < 2.2e-16
Omega-6 pct Model 2 vs. Model 1
R = 0.78, p < 2.2e-16
Omega-6:Omega-3 ratio Model 2 vs. Model 1
R = 0.93, p < 2.2e-16
DHA Model 2 vs. Model 1
R = 0.91, p < 2.2e-16
-log10(P) Model 1
-log10(P) Model 2

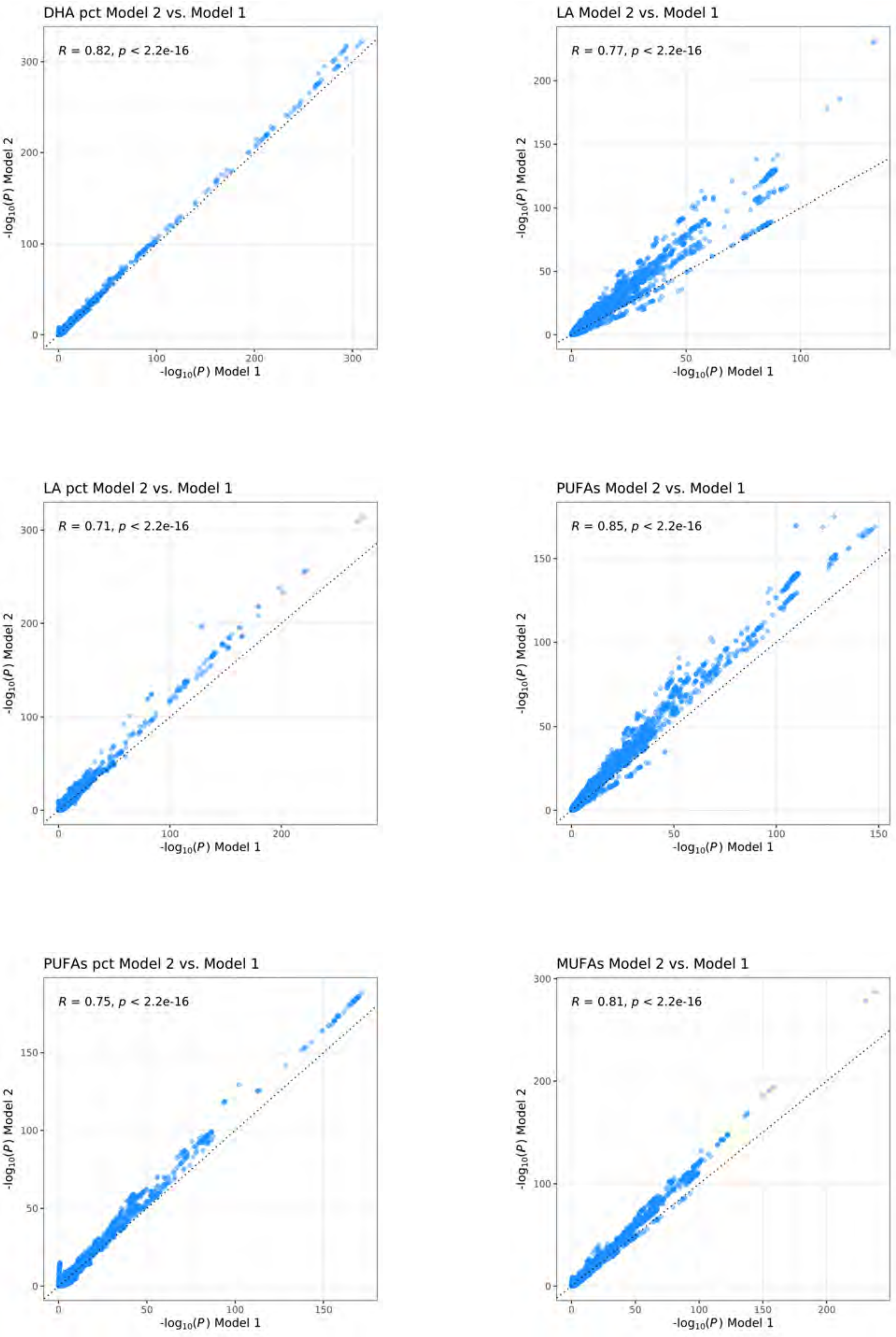
DHA pct Model 2 vs. Model 1
R = 0.82, p < 2.2e-16
-log10(P) Model 2
-log10(P) Model 1
LA Model 2 vs. Model 1
R = 0.77, p < 2.2e-16
-log10(P) Model 2
-log10(P) Model 1
LA pct Model 2 vs. Model 1
R = 0.71, p < 2.2e-16
-log10(P) Model 2
-log10(P) Model 1
PUFAs Model 2 vs. Model 1
R = 0.85, p < 2.2e-16
-log10(P) Model 2
-log10(P) Model 1
PUFAs pct Model 2 vs. Model 1
R = 0.75, p < 2.2e-16
-log10(P) Model 2
-log10(P) Model 1
MUFAs Model 2 vs. Model 1
R = 0.81, p < 2.2e-16
-log10(P) Model 2
-log10(P) Model 1

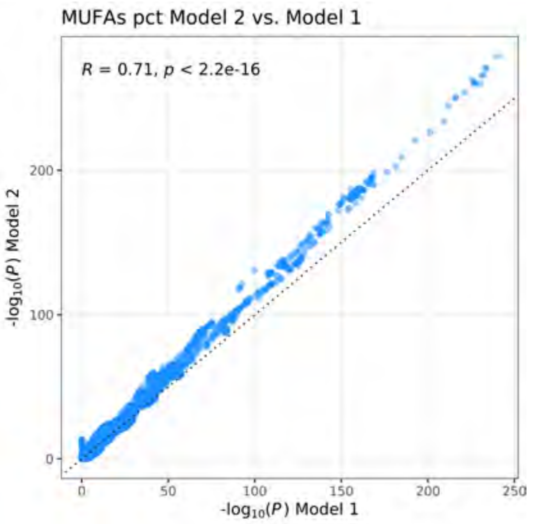
MUFAs pct Model 2 vs. Model 1
R = 0.71, p < 2.2e-16
-log10(P) Model 2
-log10(P) Model 1
0
100
200
0
50
100
150
200
250

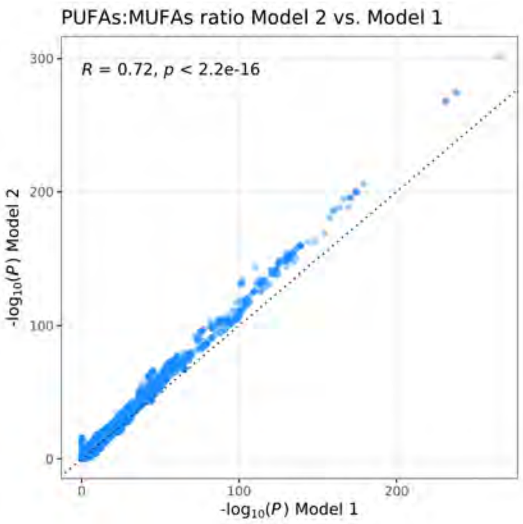
PUFAs:MUFAs ratio Model 2 vs. Model 1
R = 0.72, p < 2.2e-16
-log10(P) Model 2
-log10(P) Model 1
0
100
200
300
0
100
200

**S3**

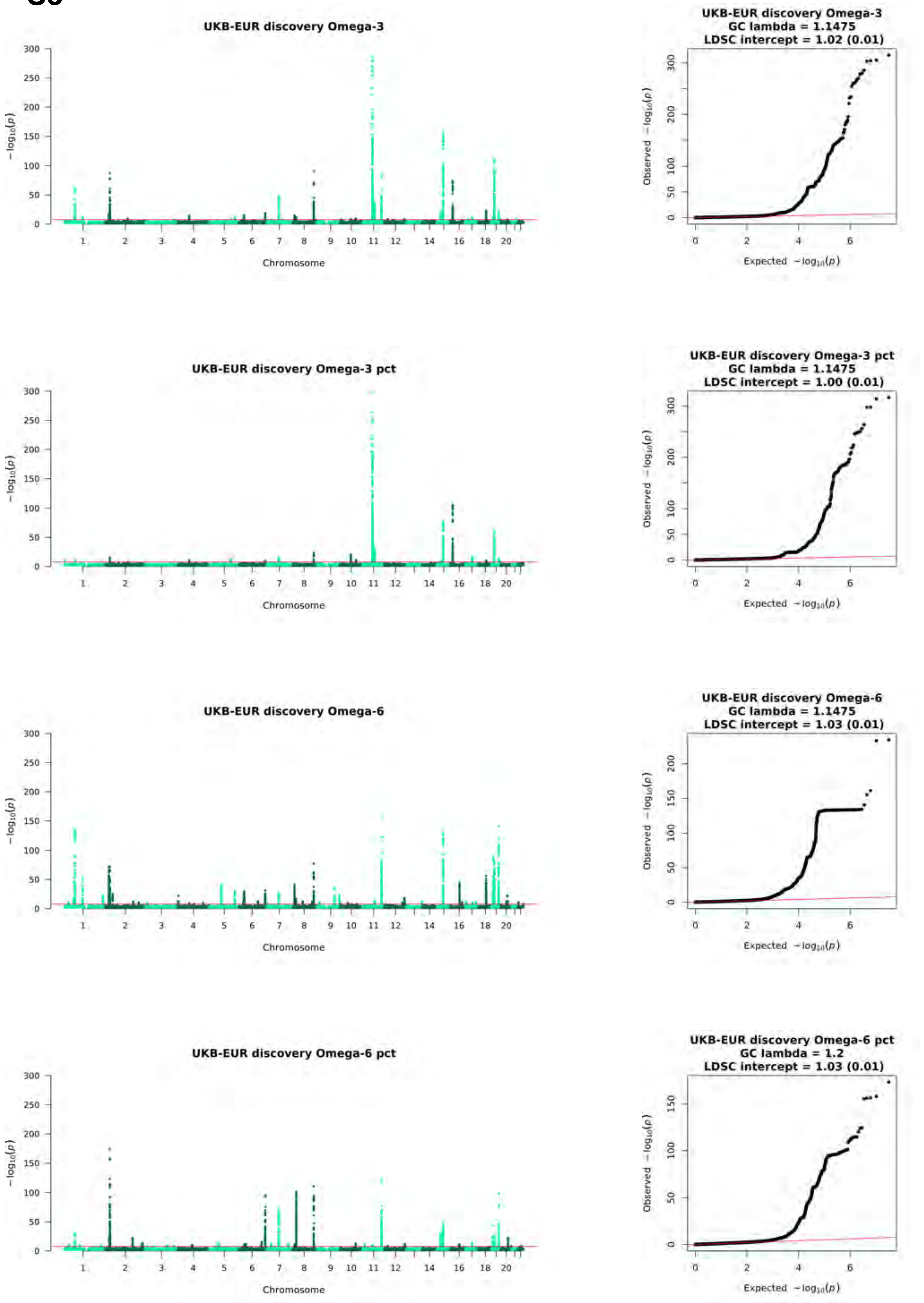
UKB-EUR discovery Omega-3
Chromosome
UKB-EUR discovery Omega-3
GC lambda = 1.1475
LDSC intercept = 1.02 (0.01)
Expected −log10(p)
Observed −log10(p)
UKB-EUR discovery Omega-3 pct
Chromosome
UKB-EUR discovery Omega-3 pct
GC lambda = 1.1475
LDSC intercept = 1.00 (0.01)
UKB-EUR discovery Omega-6
Chromosome
UKB-EUR discovery Omega-6
GC lambda = 1.1475
LDSC intercept = 1.03 (0.01)
UKB-EUR discovery Omega-6 pct
Chromosome
UKB-EUR discovery Omega-6 pct
GC lambda = 1.2
LDSC intercept = 1.03 (0.01)

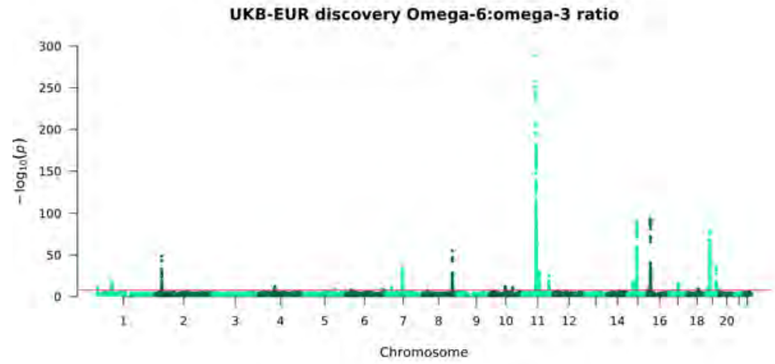
UKB-EUR discovery Omega-6:omega-3 ratio
$-\log_{10}(p)$
Chromosome

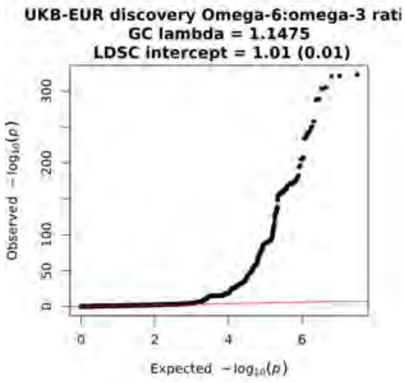
UKB-EUR discovery Omega-6:omega-3 rati
GC lambda = 1.1475
LDSC intercept = 1.01 (0.01)
Observed $-\log_{10}(p)$
Expected $-\log_{10}(p)$

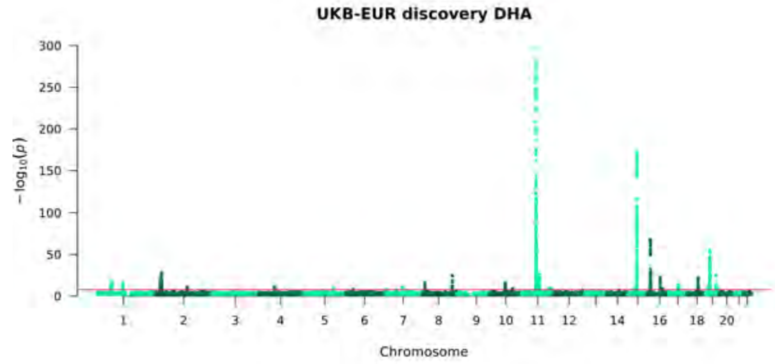
UKB-EUR discovery DHA
$-\log_{10}(p)$
Chromosome

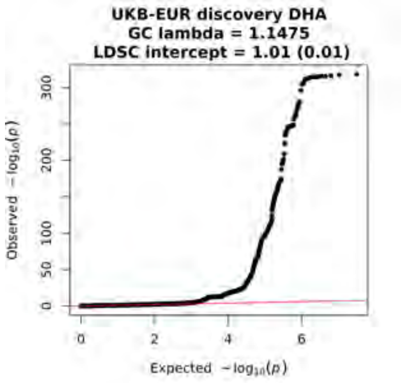
UKB-EUR discovery DHA
GC lambda = 1.1475
LDSC intercept = 1.01 (0.01)
Observed $-\log_{10}(p)$
Expected $-\log_{10}(p)$

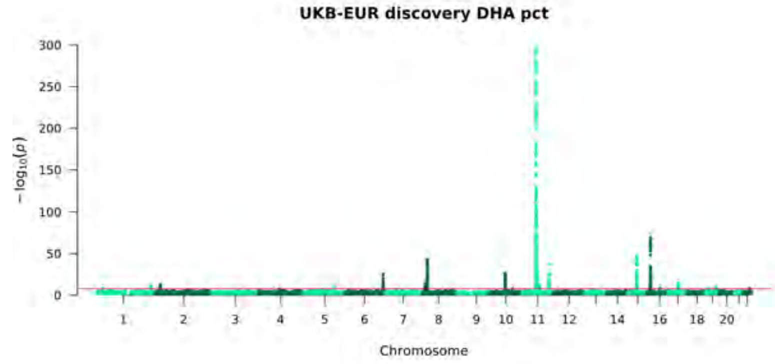
UKB-EUR discovery DHA pct
$-\log_{10}(p)$
Chromosome

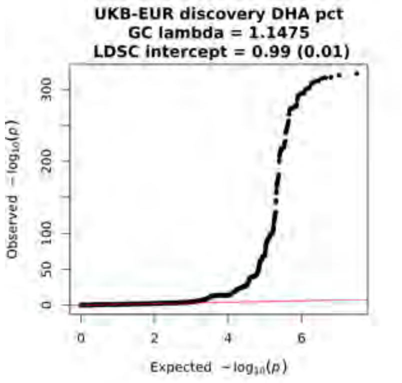
UKB-EUR discovery DHA pct
GC lambda = 1.1475
LDSC intercept = 0.99 (0.01)
Observed $-\log_{10}(p)$
Expected $-\log_{10}(p)$

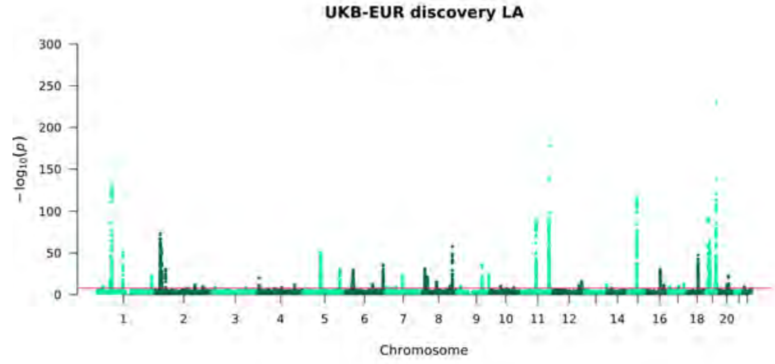
UKB-EUR discovery LA
$-\log_{10}(p)$
Chromosome

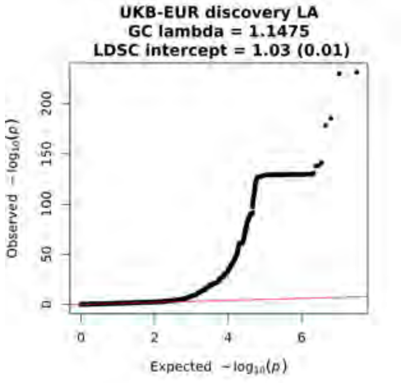
UKB-EUR discovery LA
GC lambda = 1.1475
LDSC intercept = 1.03 (0.01)
Observed $-\log_{10}(p)$
Expected $-\log_{10}(p)$

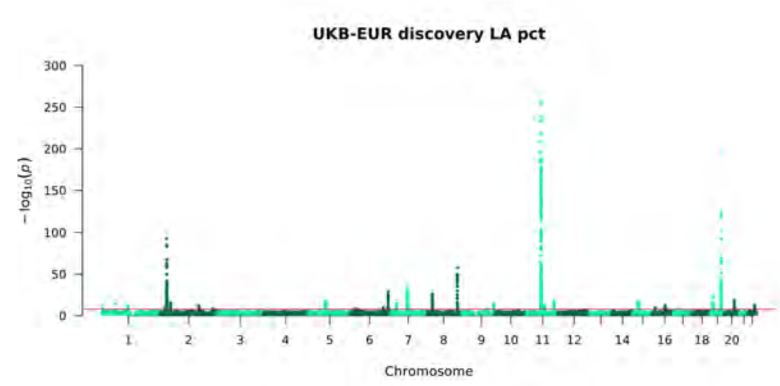
UKB-EUR discovery LA pct
$-\log_{10}(p)$
Chromosome

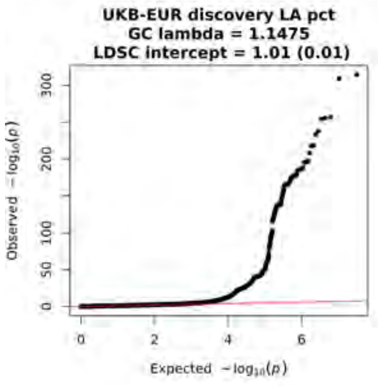
UKB-EUR discovery LA pct
GC lambda = 1.1475
LDSC intercept = 1.01 (0.01)
Observed $-\log_{10}(p)$
Expected $-\log_{10}(p)$

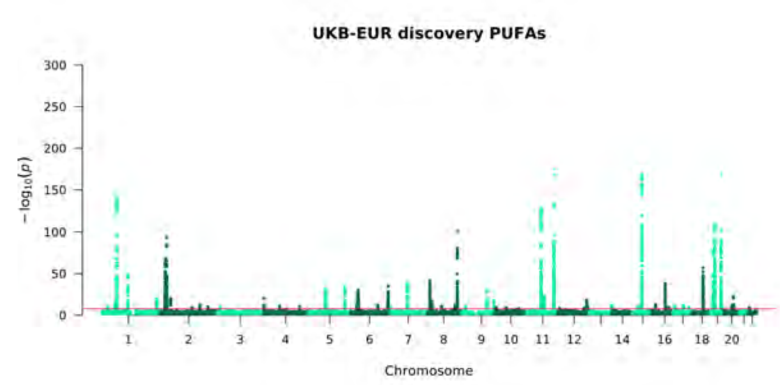
UKB-EUR discovery PUFAs
$-\log_{10}(p)$
Chromosome

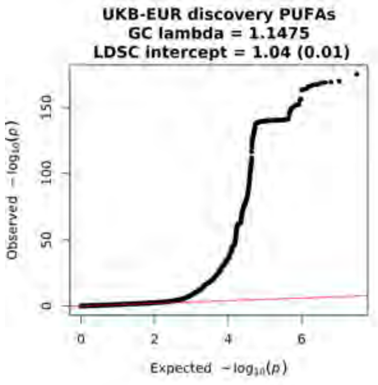
UKB-EUR discovery PUFAs
GC lambda = 1.1475
LDSC intercept = 1.04 (0.01)
Observed $-\log_{10}(p)$
Expected $-\log_{10}(p)$

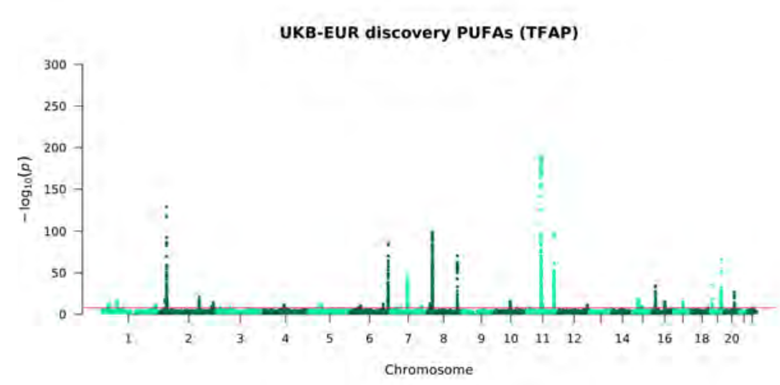
UKB-EUR discovery PUFAs (TFAP)
$-\log_{10}(p)$
Chromosome

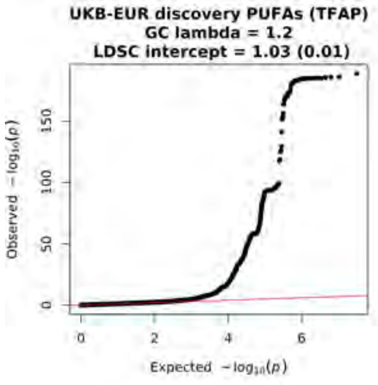
UKB-EUR discovery PUFAs (TFAP)
GC lambda = 1.2
LDSC intercept = 1.03 (0.01)
Observed $-\log_{10}(p)$
Expected $-\log_{10}(p)$

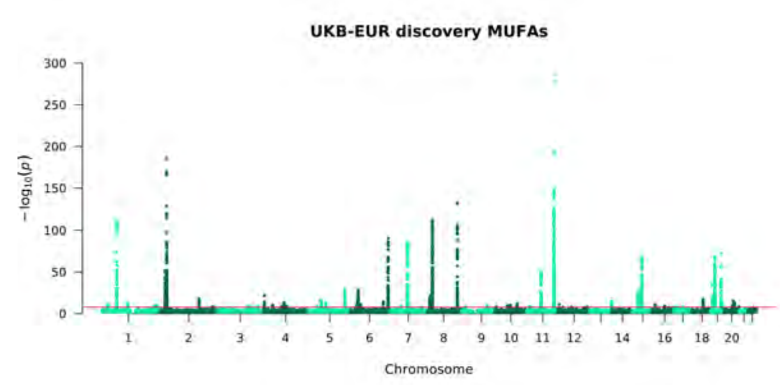
UKB-EUR discovery MUFAs
$-\log_{10}(p)$
Chromosome

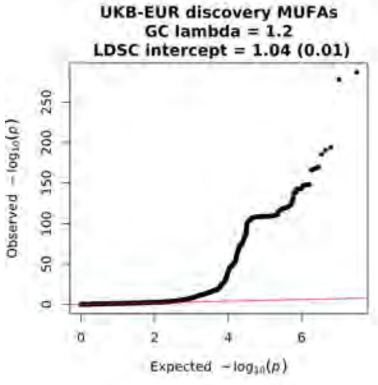
UKB-EUR discovery MUFAs
GC lambda = 1.2
LDSC intercept = 1.04 (0.01)
Observed $-\log_{10}(p)$
Expected $-\log_{10}(p)$

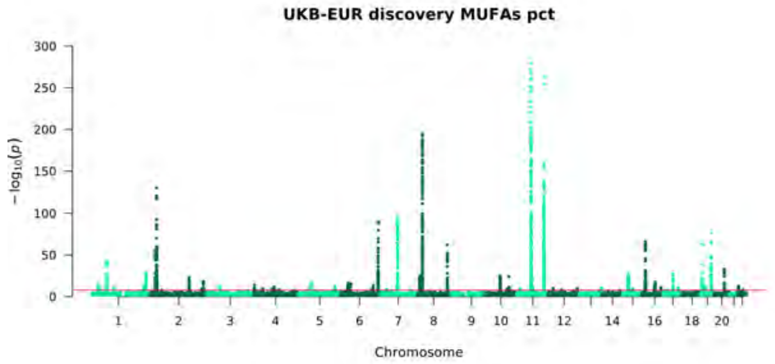
UKB-EUR discovery MUFAs pct
−log10(p)
Chromosome

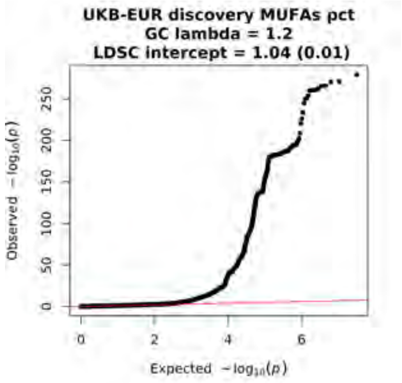
UKB-EUR discovery MUFAs pct
GC lambda = 1.2
LDSC intercept = 1.04 (0.01)
Observed −log10(p)
Expected −log10(p)

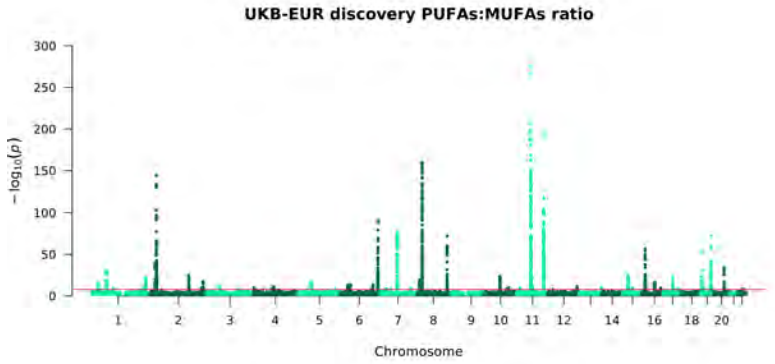
UKB-EUR discovery PUFAs:MUFAs ratio
−log10(p)
Chromosome

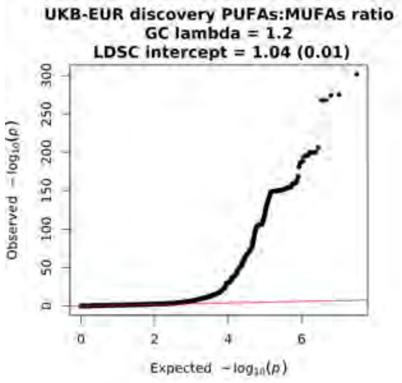
UKB-EUR discovery PUFAs:MUFAs ratio
GC lambda = 1.2
LDSC intercept = 1.04 (0.01)
Observed −log10(p)
Expected −log10(p)

**S4**

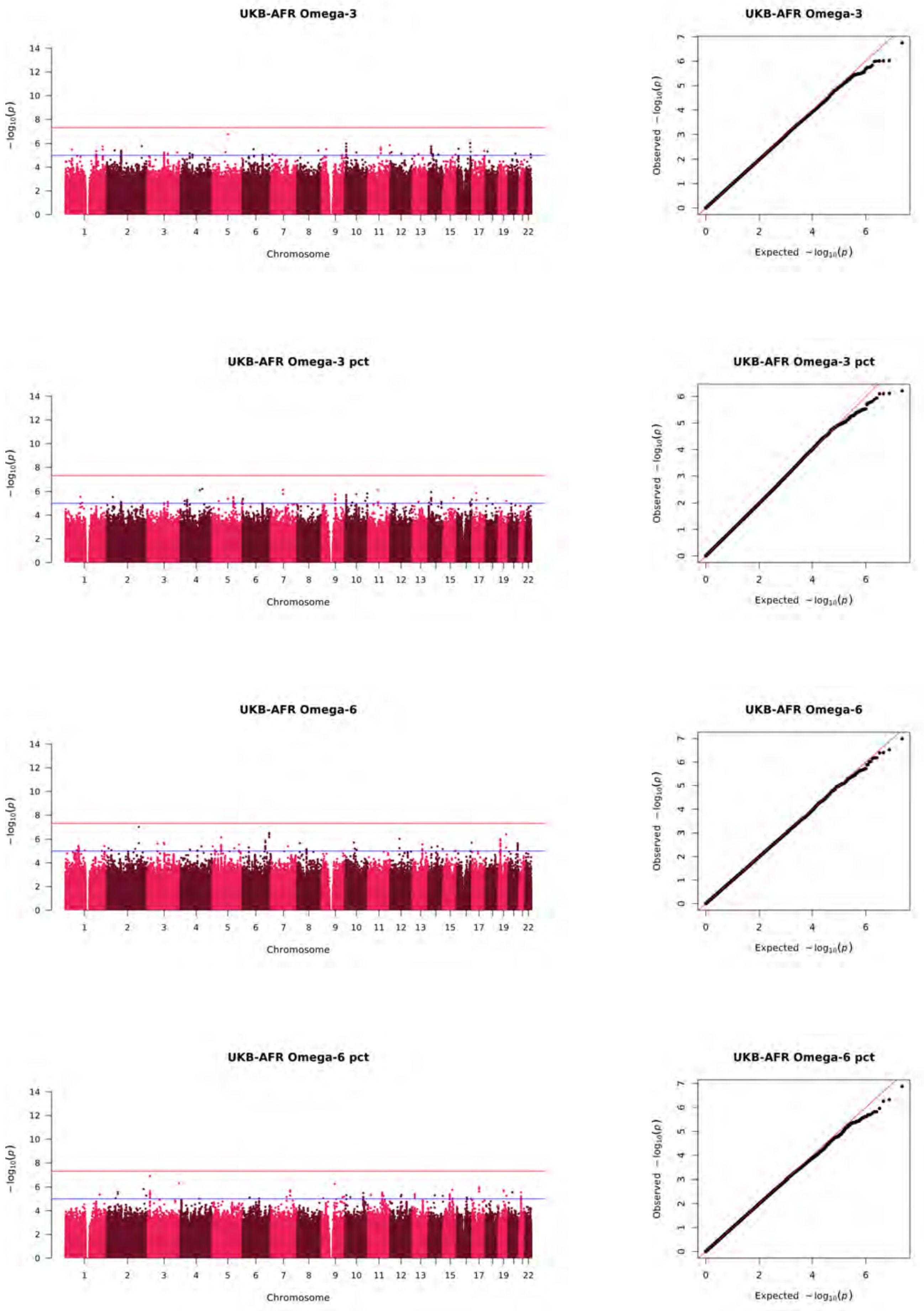
UKB-AFR Omega-3
UKB-AFR Omega-3
UKB-AFR Omega-3 pct
UKB-AFR Omega-3 pct
UKB-AFR Omega-6
UKB-AFR Omega-6
UKB-AFR Omega-6 pct
UKB-AFR Omega-6 pct
Chromosome
$-\log_{10}(p)$
Observed $-\log_{10}(p)$
Expected $-\log_{10}(p)$

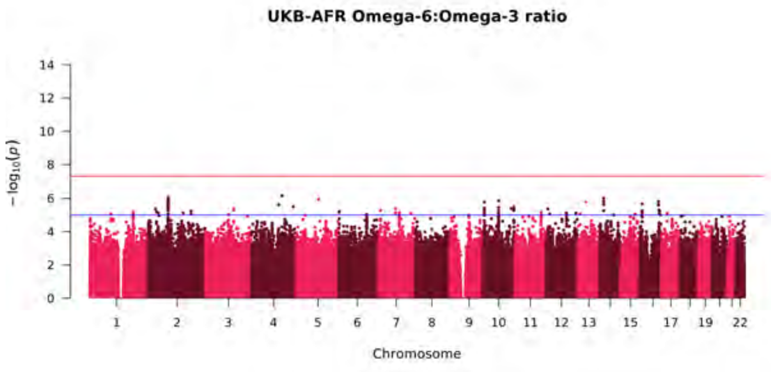
UKB-AFR Omega-6:Omega-3 ratio
$-\log_{10}(p)$
Chromosome

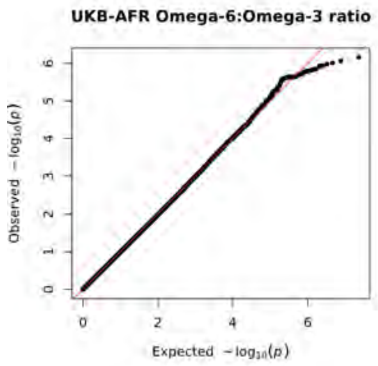
UKB-AFR Omega-6:Omega-3 ratio
Observed $-\log_{10}(p)$
Expected $-\log_{10}(p)$

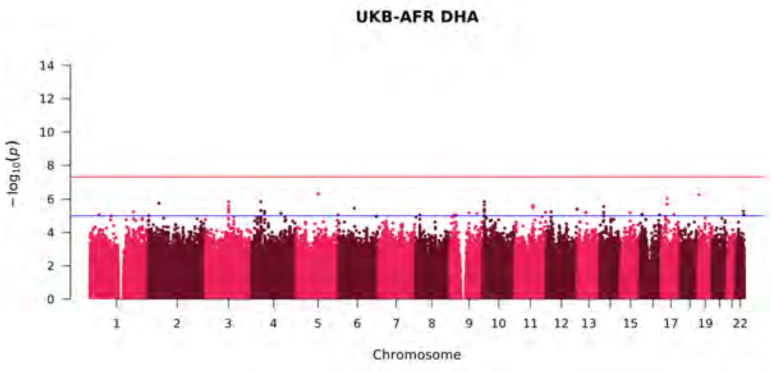
UKB-AFR DHA
$-\log_{10}(p)$
Chromosome

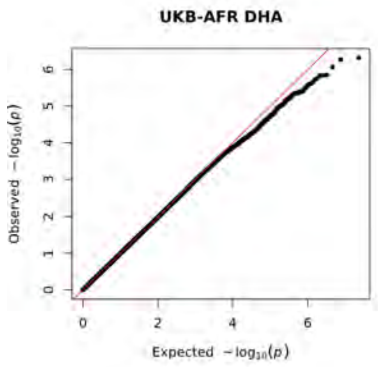
UKB-AFR DHA
Observed $-\log_{10}(p)$
Expected $-\log_{10}(p)$

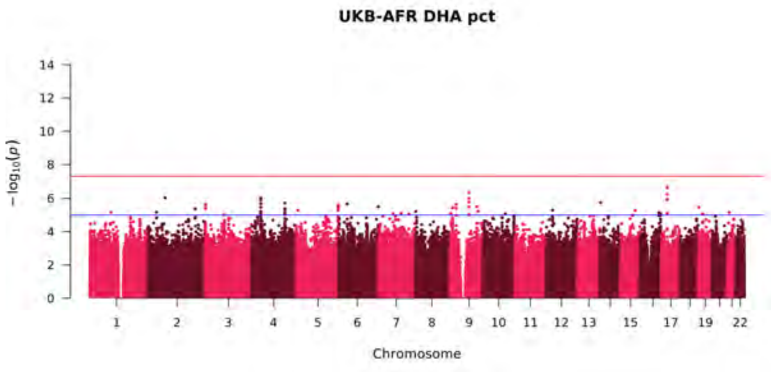
UKB-AFR DHA pct
$-\log_{10}(p)$
Chromosome

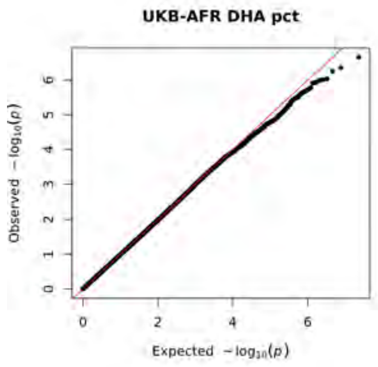
UKB-AFR DHA pct
Observed $-\log_{10}(p)$
Expected $-\log_{10}(p)$

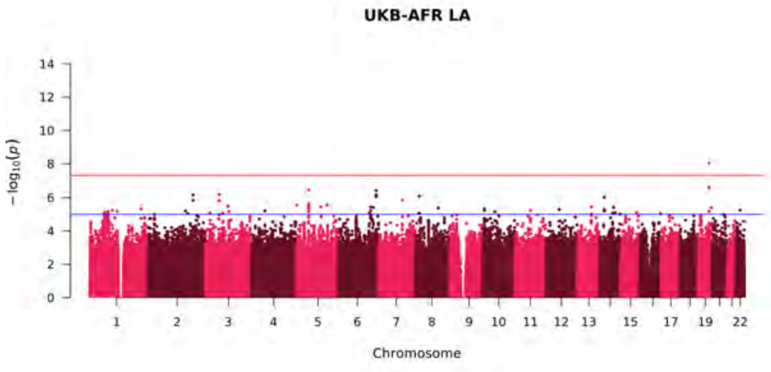
UKB-AFR LA
$-\log_{10}(p)$
Chromosome

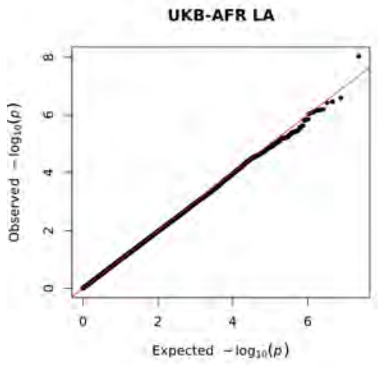
UKB-AFR LA
Observed $-\log_{10}(p)$
Expected $-\log_{10}(p)$

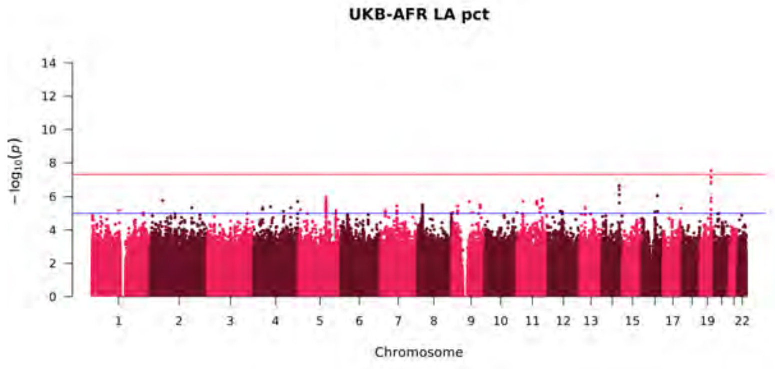
UKB-AFR LA pct
$-\log_{10}(p)$
Chromosome

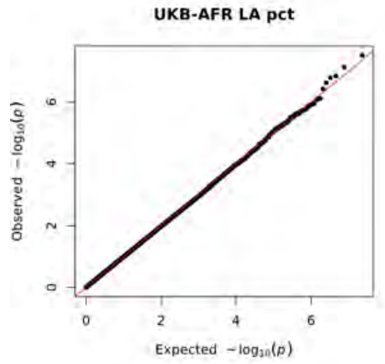
UKB-AFR LA pct
Observed $-\log_{10}(p)$
Expected $-\log_{10}(p)$

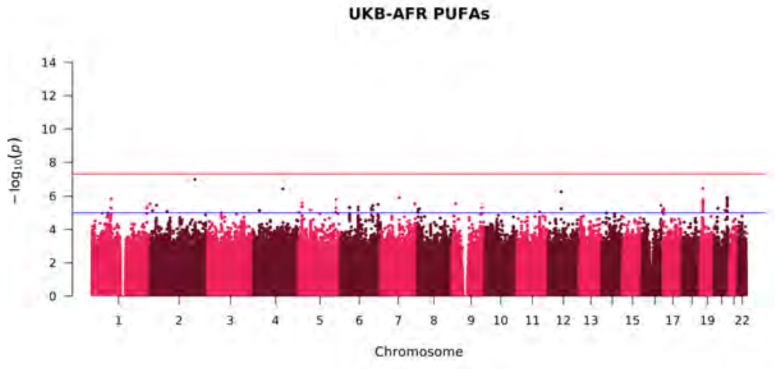
UKB-AFR PUFAs
$-\log_{10}(p)$
Chromosome

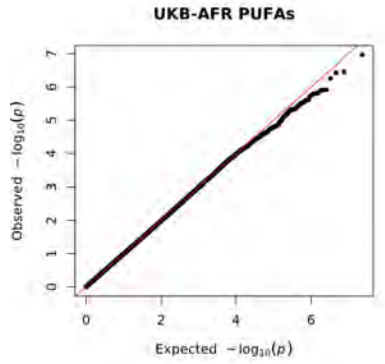
UKB-AFR PUFAs
Observed $-\log_{10}(p)$
Expected $-\log_{10}(p)$

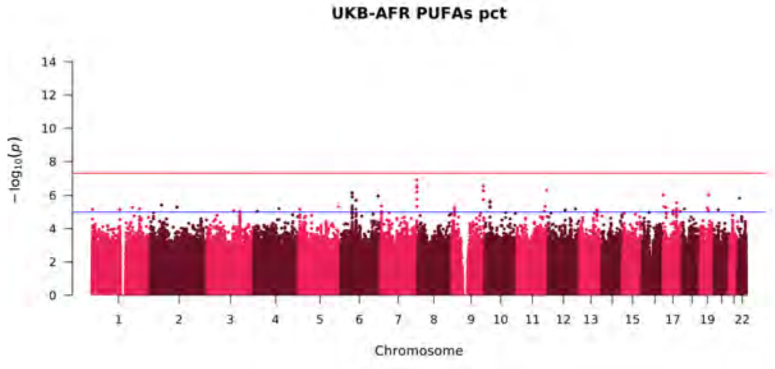
UKB-AFR PUFAs pct
$-\log_{10}(p)$
Chromosome

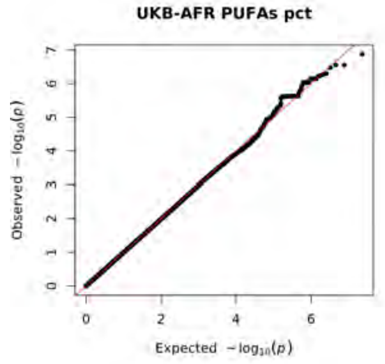
UKB-AFR PUFAs pct
Observed $-\log_{10}(p)$
Expected $-\log_{10}(p)$

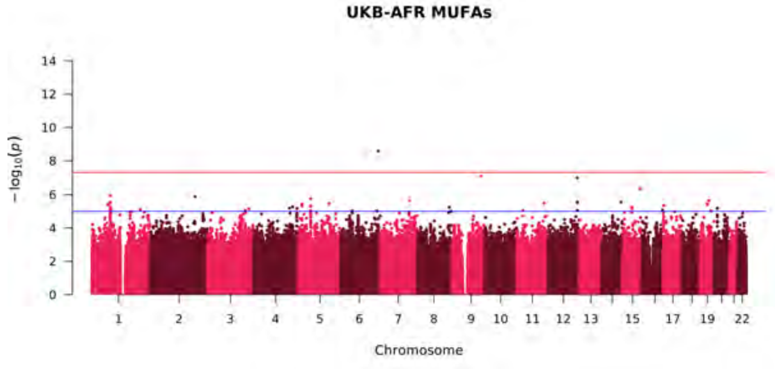
UKB-AFR MUFAs
$-\log_{10}(p)$
Chromosome

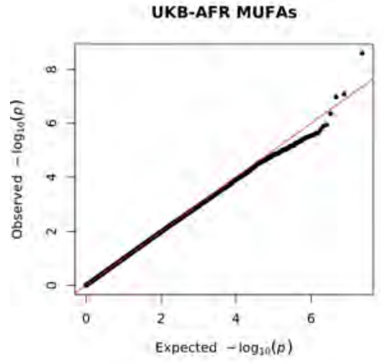
UKB-AFR MUFAs
Observed $-\log_{10}(p)$
Expected $-\log_{10}(p)$

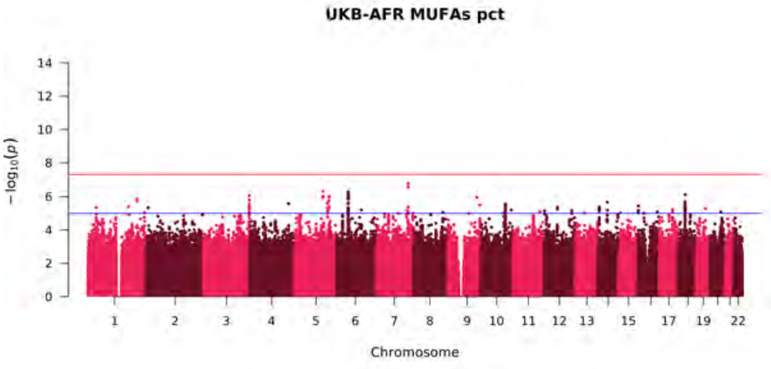
UKB-AFR MUFAs pct
$-\log_{10}(p)$
Chromosome

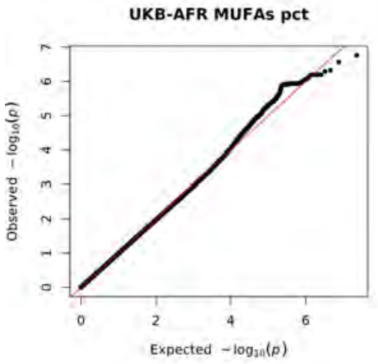
UKB-AFR MUFAs pct
Observed $-\log_{10}(p)$
Expected $-\log_{10}(p)$

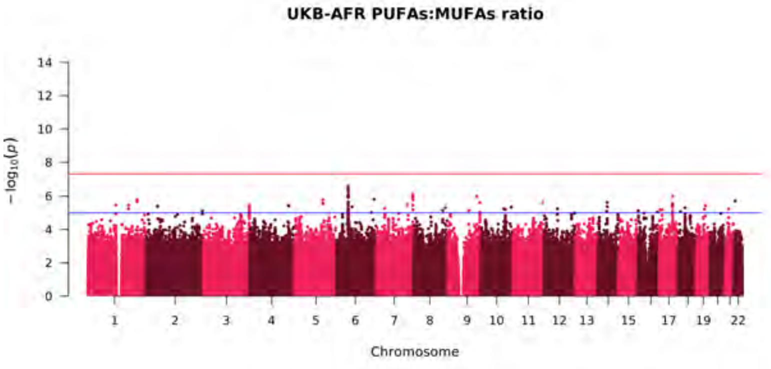
UKB-AFR PUFAs:MUFAs ratio
$-\log_{10}(p)$
Chromosome

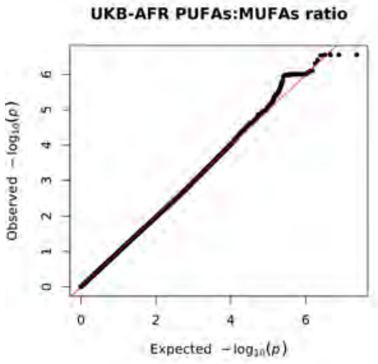
UKB-AFR PUFAs:MUFAs ratio
Observed $-\log_{10}(p)$
Expected $-\log_{10}(p)$

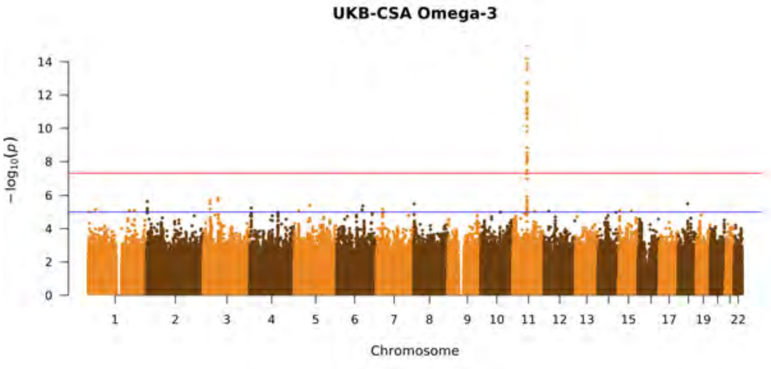
UKB-CSA Omega-3
$-\log_{10}(p)$
Chromosome

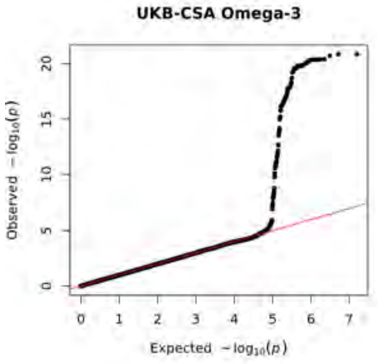
UKB-CSA Omega-3
Observed $-\log_{10}(p)$
Expected $-\log_{10}(p)$

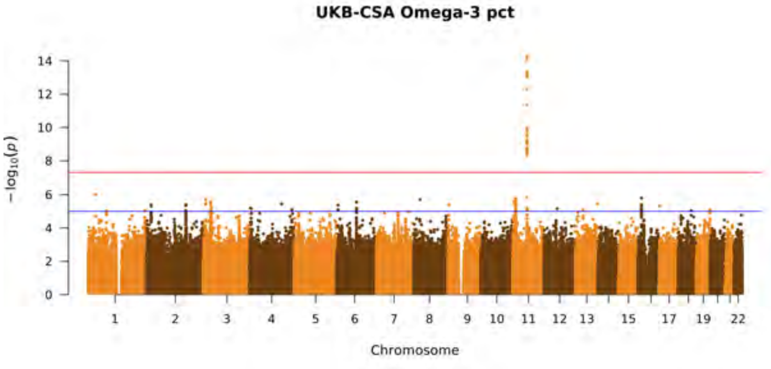
UKB-CSA Omega-3 pct
$-\log_{10}(p)$
Chromosome

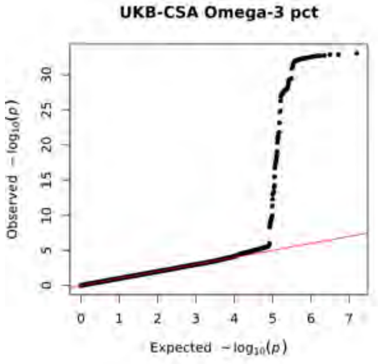
UKB-CSA Omega-3 pct
Observed $-\log_{10}(p)$
Expected $-\log_{10}(p)$

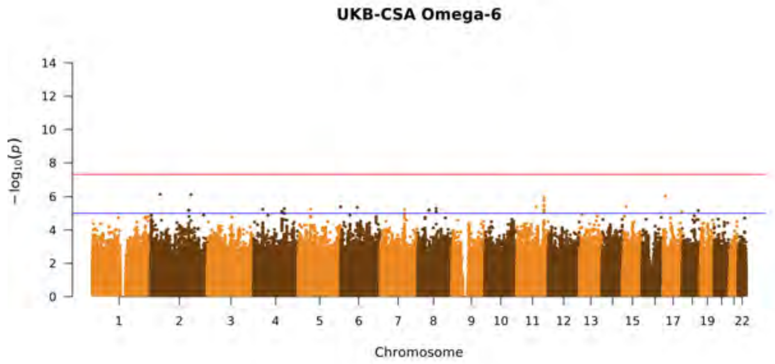
UKB-CSA Omega-6
$-\log_{10}(p)$
Chromosome

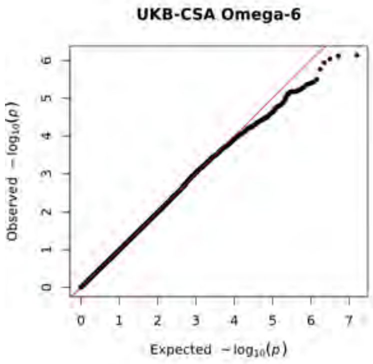
UKB-CSA Omega-6
Observed $-\log_{10}(p)$
Expected $-\log_{10}(p)$

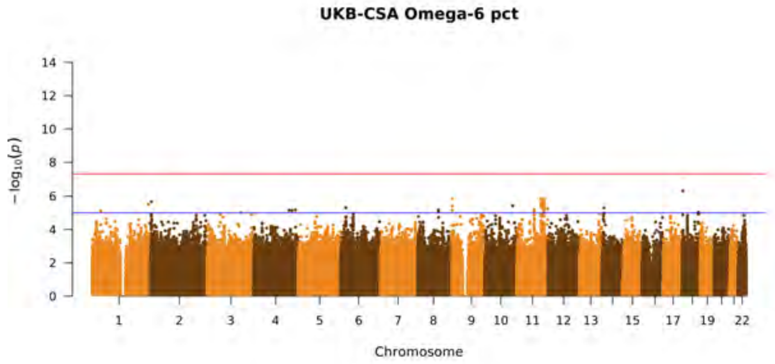
UKB-CSA Omega-6 pct
$-\log_{10}(p)$
Chromosome

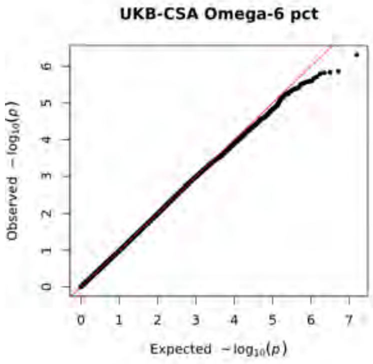
UKB-CSA Omega-6 pct
Observed $-\log_{10}(p)$
Expected $-\log_{10}(p)$

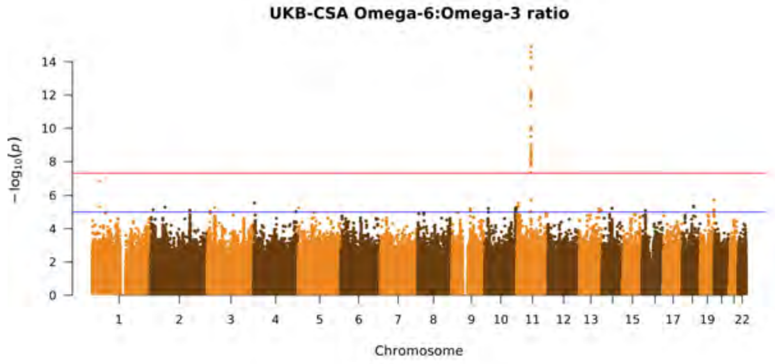
UKB-CSA Omega-6:Omega-3 ratio
$-\log_{10}(p)$
Chromosome

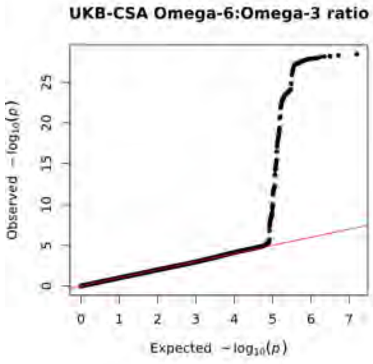
UKB-CSA Omega-6:Omega-3 ratio
Observed $-\log_{10}(p)$
Expected $-\log_{10}(p)$

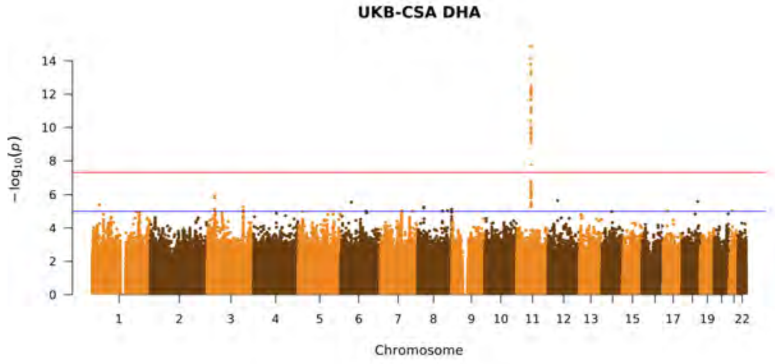
UKB-CSA DHA
$-\log_{10}(p)$
Chromosome

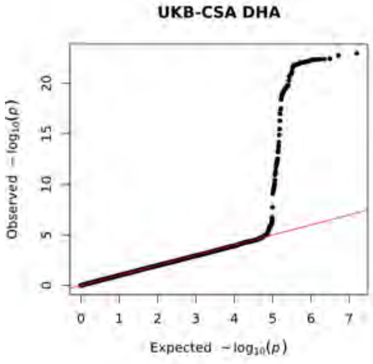
UKB-CSA DHA
Observed $-\log_{10}(p)$
Expected $-\log_{10}(p)$

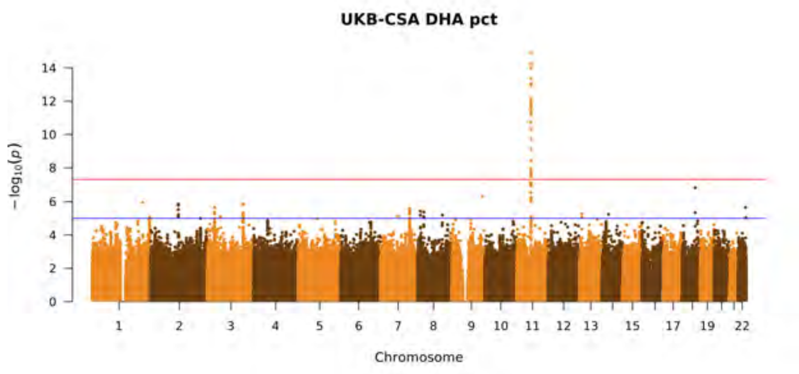
UKB-CSA DHA pct
$-\log_{10}(p)$
Chromosome

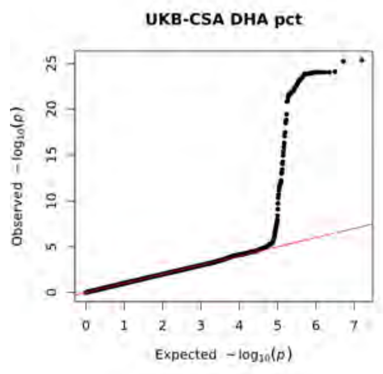
UKB-CSA DHA pct
Observed $-\log_{10}(p)$
Expected $-\log_{10}(p)$

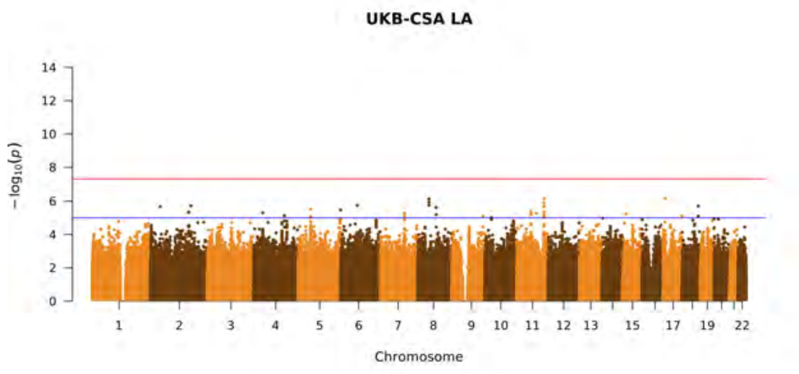
UKB-CSA LA
$-\log_{10}(p)$
Chromosome

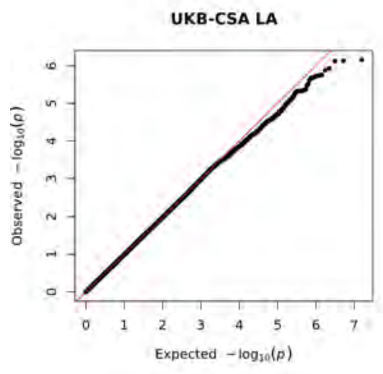
UKB-CSA LA
Observed $-\log_{10}(p)$
Expected $-\log_{10}(p)$

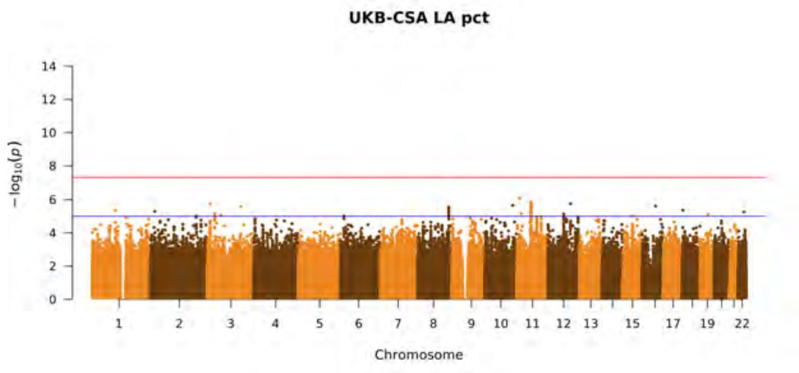
UKB-CSA LA pct
$-\log_{10}(p)$
Chromosome

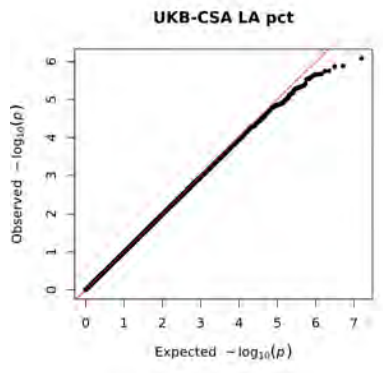
UKB-CSA LA pct
Observed $-\log_{10}(p)$
Expected $-\log_{10}(p)$

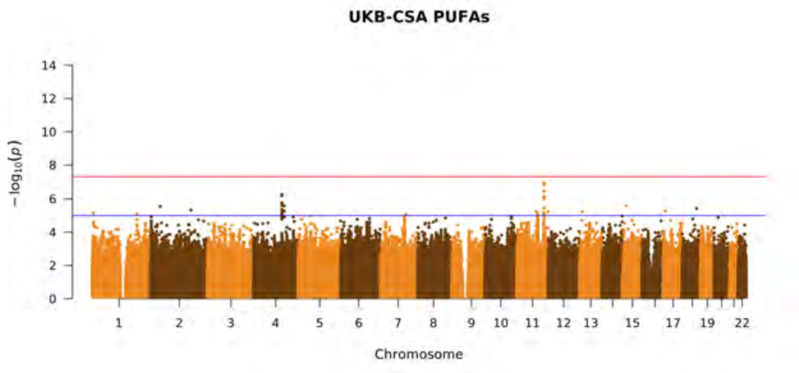
UKB-CSA PUFAs
$-\log_{10}(p)$
Chromosome

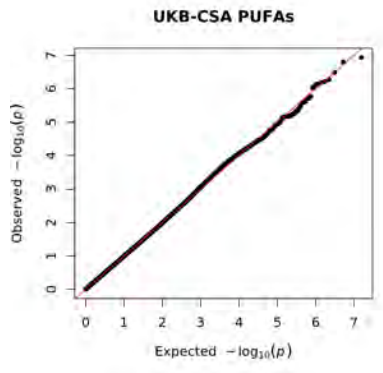
UKB-CSA PUFAs
Observed $-\log_{10}(p)$
Expected $-\log_{10}(p)$

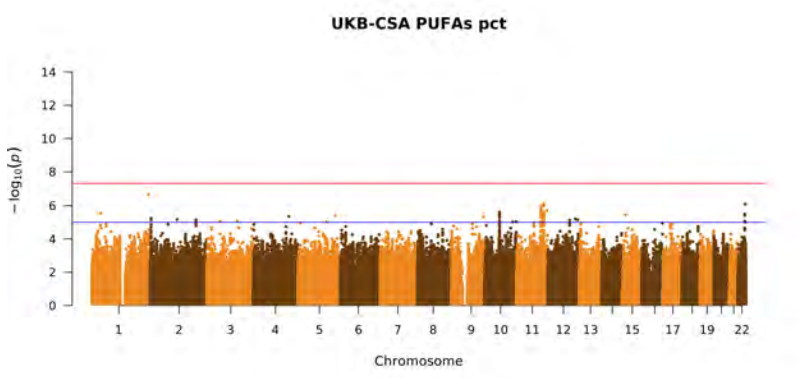
UKB-CSA PUFAs pct
−log10(p)
Chromosome

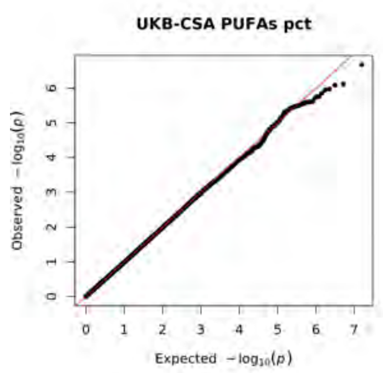
UKB-CSA PUFAs pct
Observed −log10(p)
Expected −log10(p)

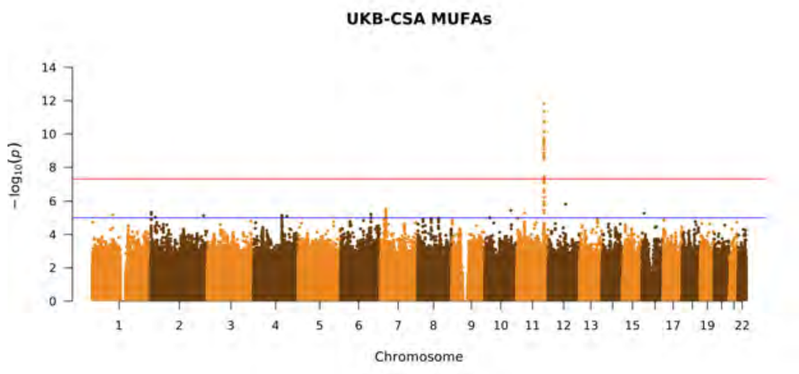
UKB-CSA MUFAs
−log10(p)
Chromosome

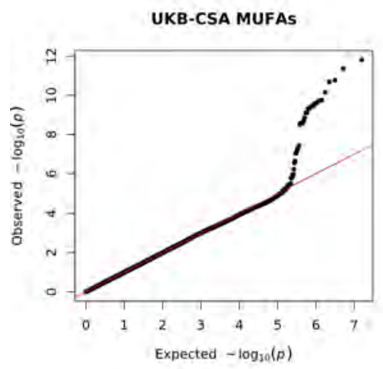
UKB-CSA MUFAs
Observed −log10(p)
Expected −log10(p)

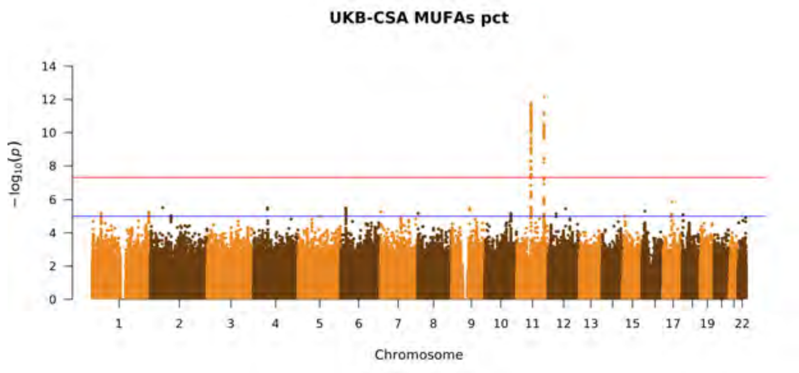
UKB-CSA MUFAs pct
−log10(p)
Chromosome

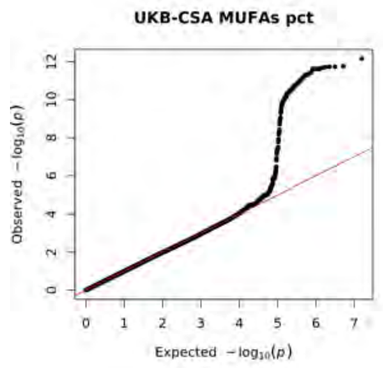
UKB-CSA MUFAs pct
Observed −log10(p)
Expected −log10(p)

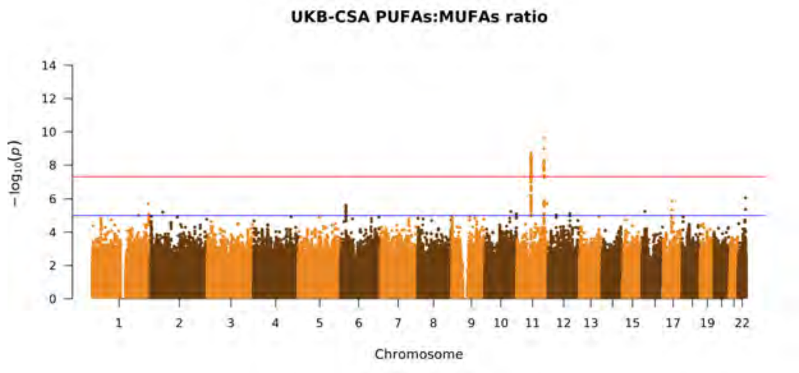
UKB-CSA PUFAs:MUFAs ratio
−log10(p)
Chromosome

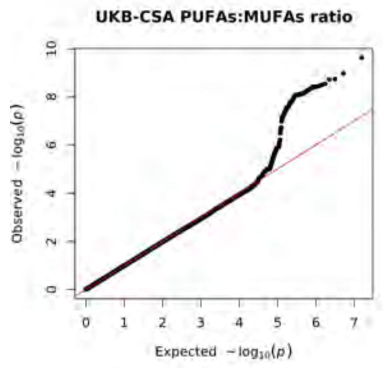
UKB-CSA PUFAs:MUFAs ratio
Observed −log10(p)
Expected −log10(p)

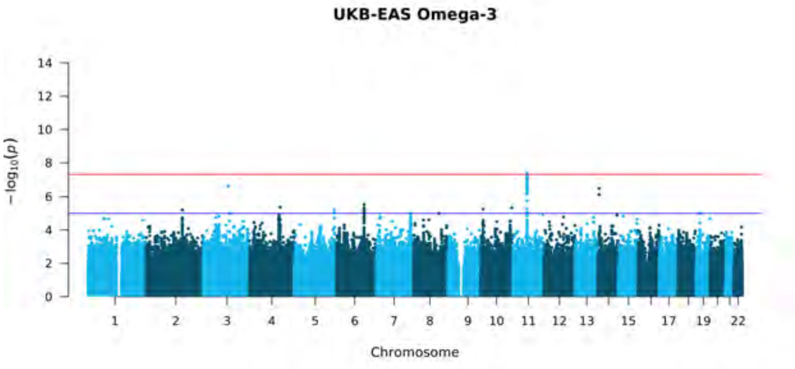
UKB-EAS Omega-3
$-\log_{10}(p)$
Chromosome

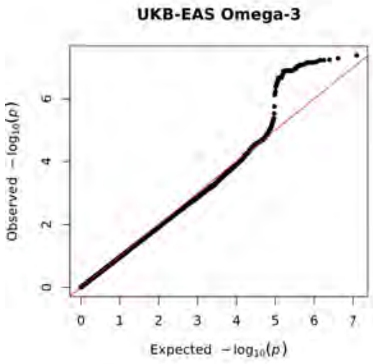
UKB-EAS Omega-3
Observed $-\log_{10}(p)$
Expected $-\log_{10}(p)$

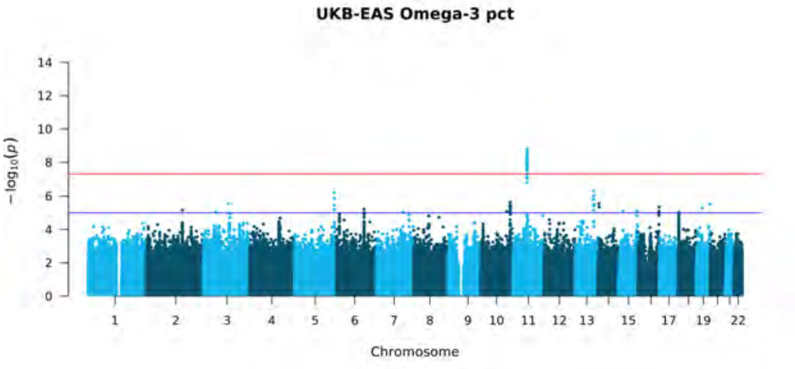
UKB-EAS Omega-3 pct
$-\log_{10}(p)$
Chromosome

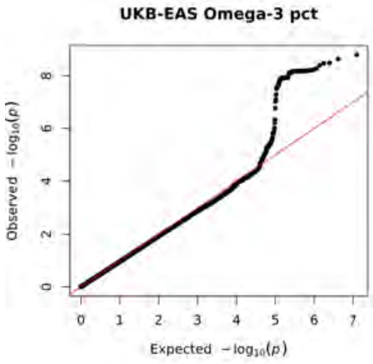
UKB-EAS Omega-3 pct
Observed $-\log_{10}(p)$
Expected $-\log_{10}(p)$

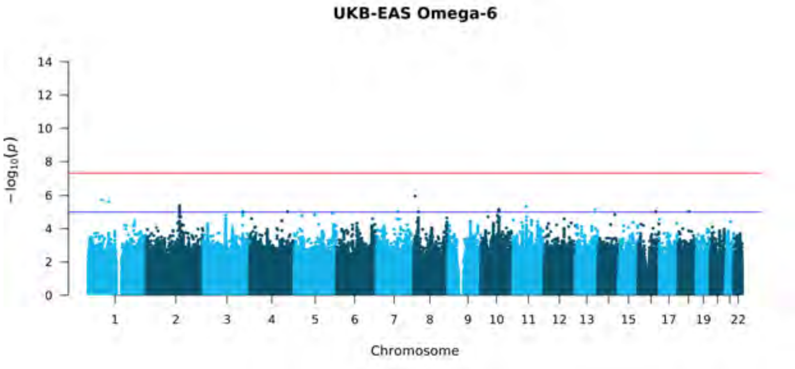
UKB-EAS Omega-6
$-\log_{10}(p)$
Chromosome

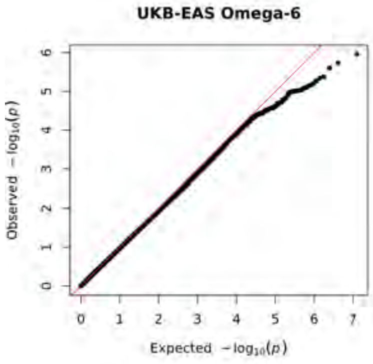
UKB-EAS Omega-6
Observed $-\log_{10}(p)$
Expected $-\log_{10}(p)$

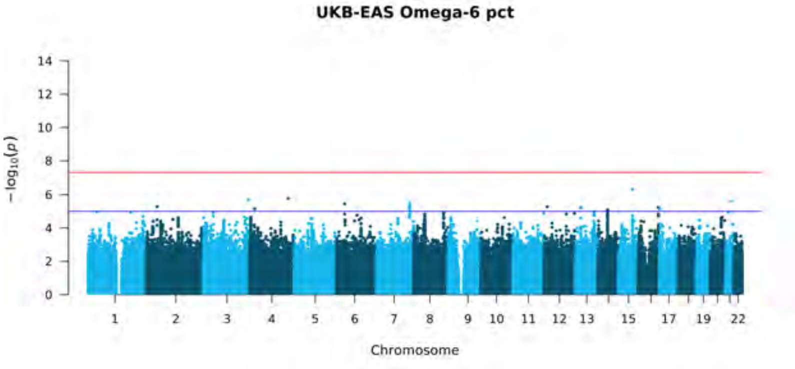
UKB-EAS Omega-6 pct
$-\log_{10}(p)$
Chromosome

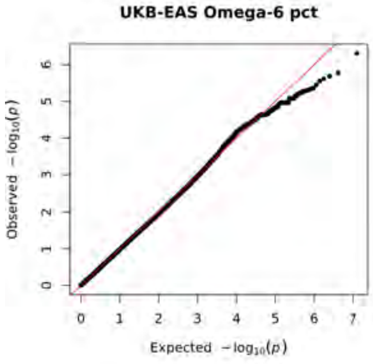
UKB-EAS Omega-6 pct
Observed $-\log_{10}(p)$
Expected $-\log_{10}(p)$

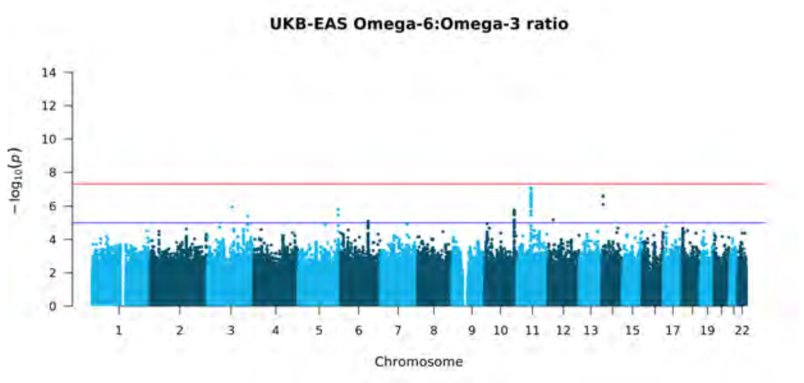
UKB-EAS Omega-6:Omega-3 ratio
$-\log_{10}(p)$
Chromosome

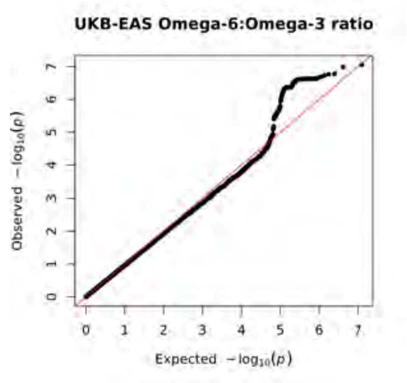
UKB-EAS Omega-6:Omega-3 ratio
Observed $-\log_{10}(p)$
Expected $-\log_{10}(p)$

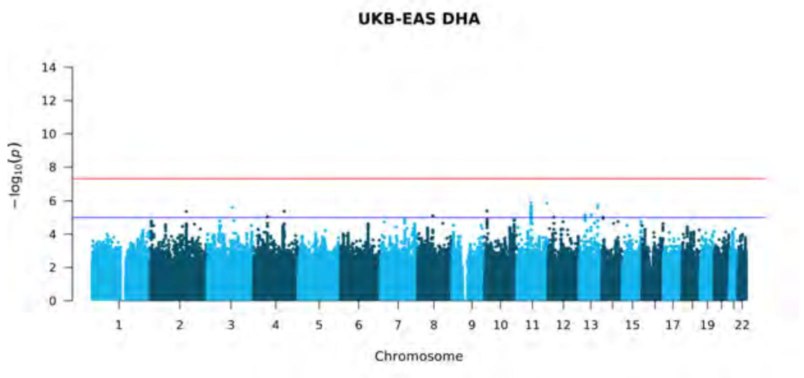
UKB-EAS DHA
$-\log_{10}(p)$
Chromosome

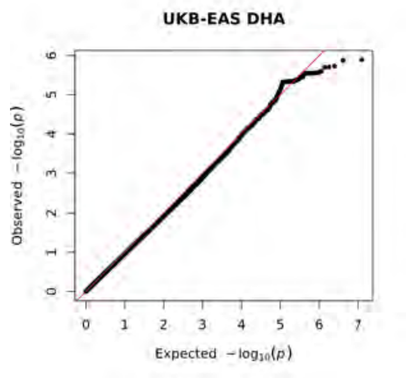
UKB-EAS DHA
Observed $-\log_{10}(p)$
Expected $-\log_{10}(p)$

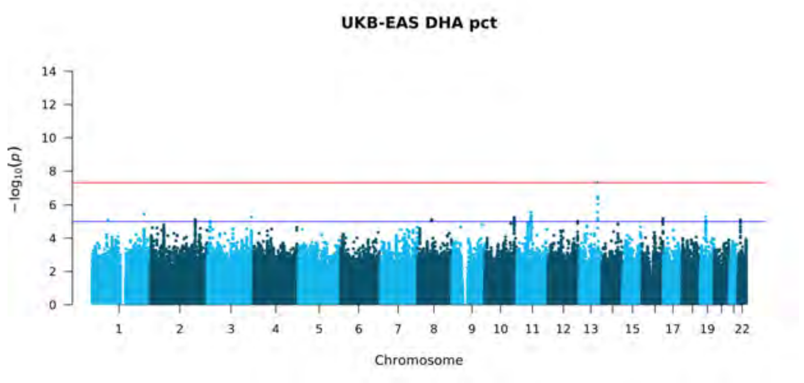
UKB-EAS DHA pct
$-\log_{10}(p)$
Chromosome

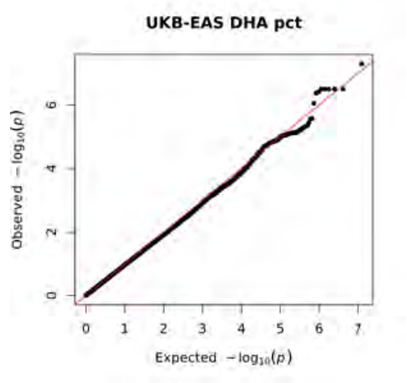
UKB-EAS DHA pct
Observed $-\log_{10}(p)$
Expected $-\log_{10}(p)$

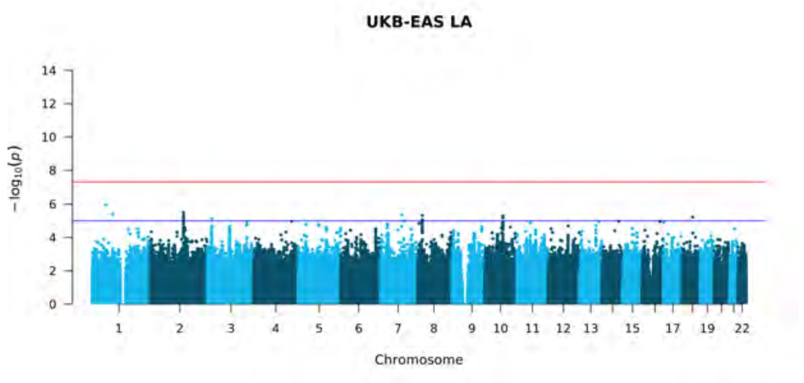
UKB-EAS LA
$-\log_{10}(p)$
Chromosome

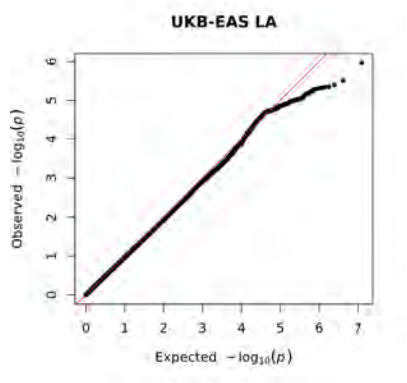
UKB-EAS LA
Observed $-\log_{10}(p)$
Expected $-\log_{10}(p)$

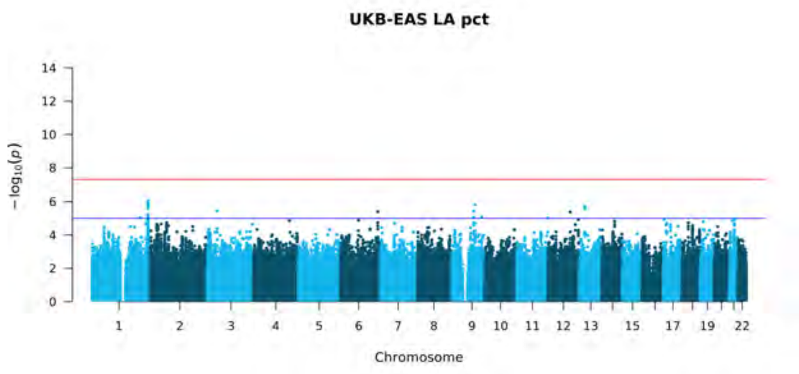
UKB-EAS LA pct
$-\log_{10}(p)$
Chromosome

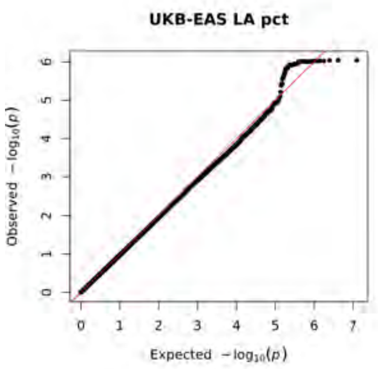
UKB-EAS LA pct
Observed $-\log_{10}(p)$
Expected $-\log_{10}(p)$

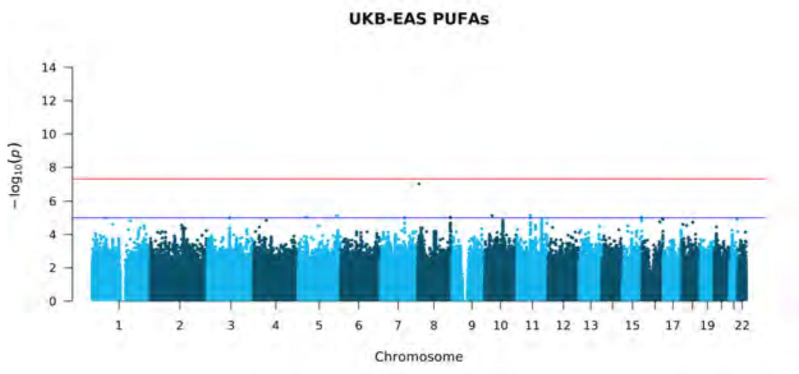
UKB-EAS PUFAs
$-\log_{10}(p)$
Chromosome

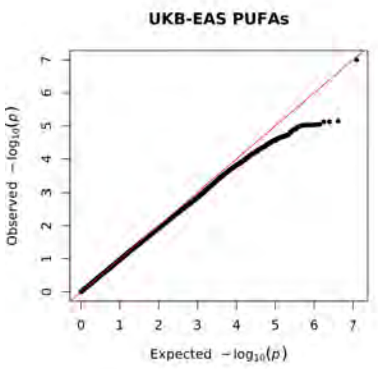
UKB-EAS PUFAs
Observed $-\log_{10}(p)$
Expected $-\log_{10}(p)$

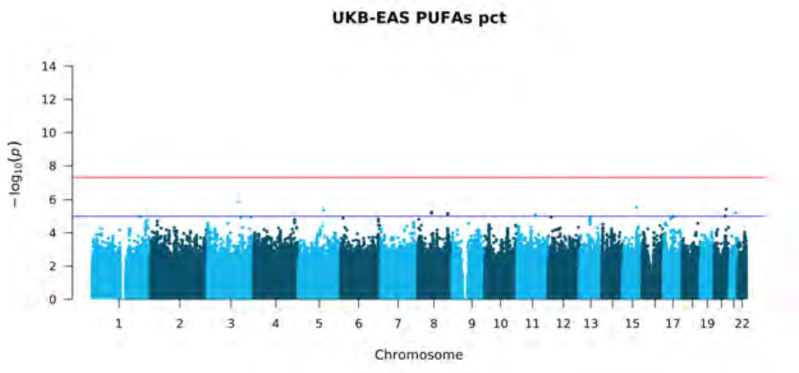
UKB-EAS PUFAs pct
$-\log_{10}(p)$
Chromosome

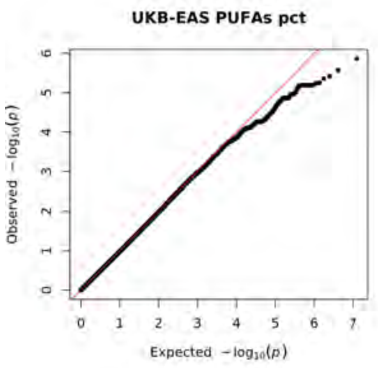
UKB-EAS PUFAs pct
Observed $-\log_{10}(p)$
Expected $-\log_{10}(p)$

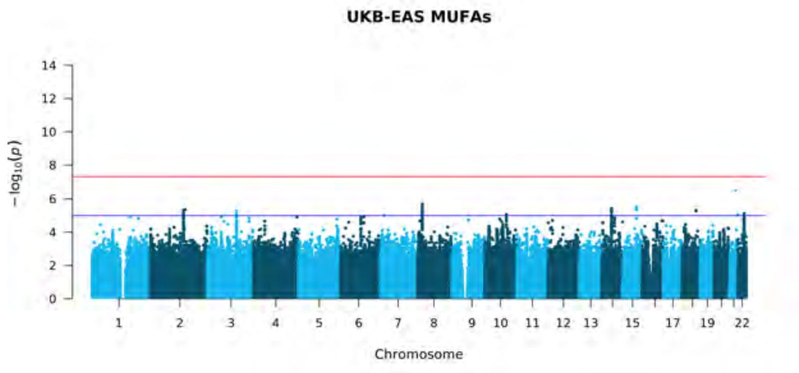
UKB-EAS MUFAs
$-\log_{10}(p)$
Chromosome

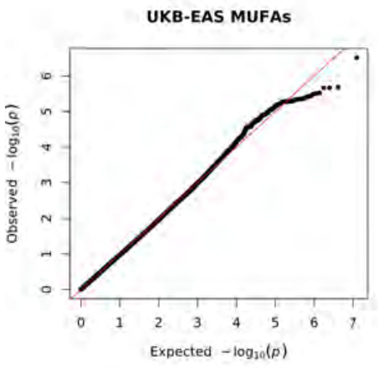
UKB-EAS MUFAs
Observed $-\log_{10}(p)$
Expected $-\log_{10}(p)$

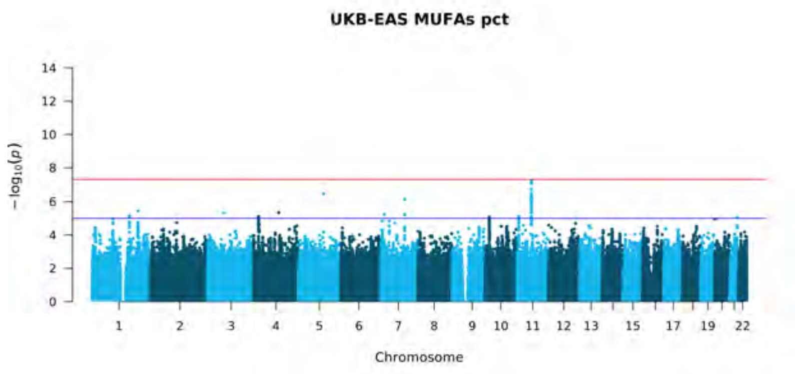
UKB-EAS MUFAs pct
$-\log_{10}(p)$
Chromosome

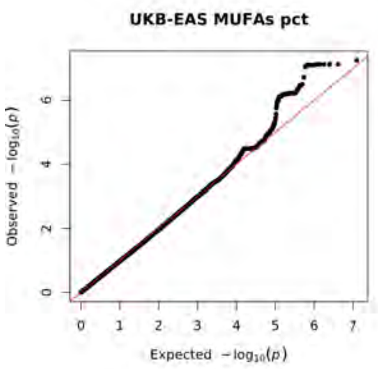
UKB-EAS MUFAs pct
Observed $-\log_{10}(p)$
Expected $-\log_{10}(p)$

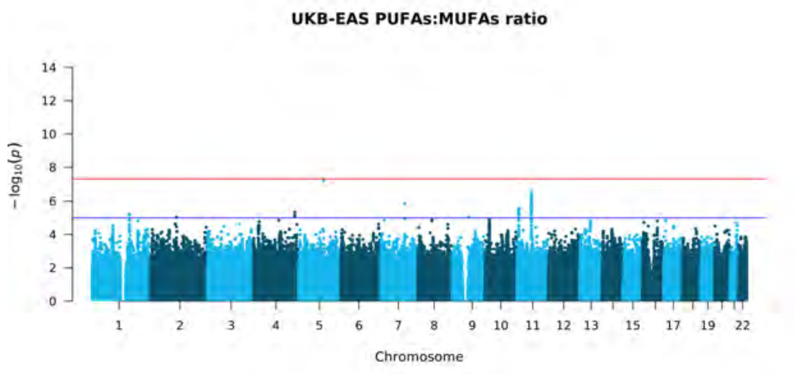
UKB-EAS PUFAs:MUFAs ratio
$-\log_{10}(p)$
Chromosome

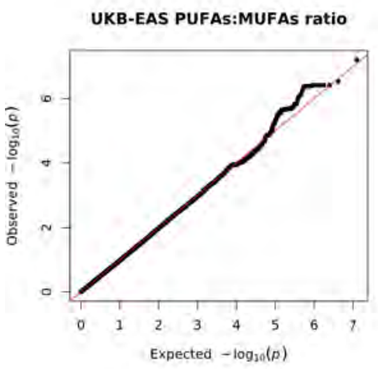
UKB-EAS PUFAs:MUFAs ratio
Observed $-\log_{10}(p)$
Expected $-\log_{10}(p)$

**S5**

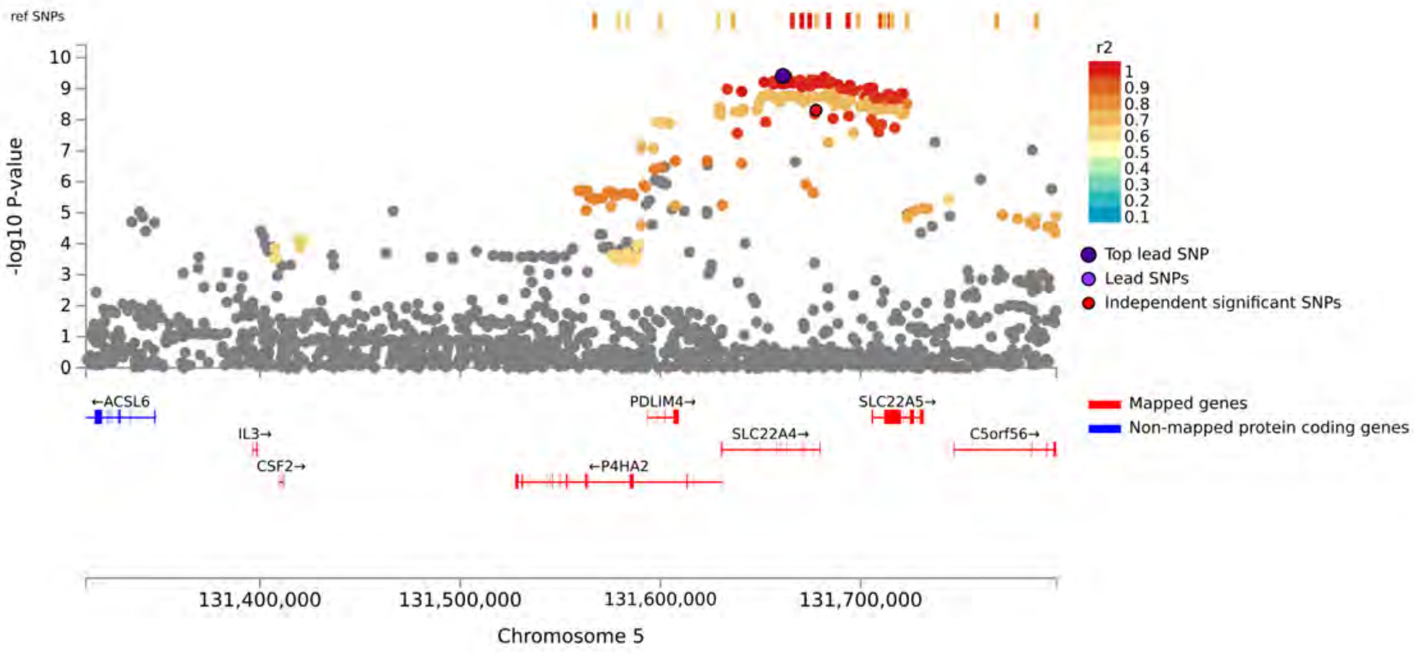
ref SNPs
-log10 P-value
10
9
8
7
6
5
4
3
2
1
0
r2
1
0.9
0.8
0.7
0.6
0.5
0.4
0.3
0.2
0.1
Top lead SNP
Lead SNPs
Independent significant SNPs
←ACSL6
IL3→
CSF2→
PDLIM4→
←P4HA2
SLC22A4→
SLC22A5→
C5orf56→
Mapped genes
Non-mapped protein coding genes
131,400,000
131,500,000
131,600,000
131,700,000
Chromosome 5

**S6**

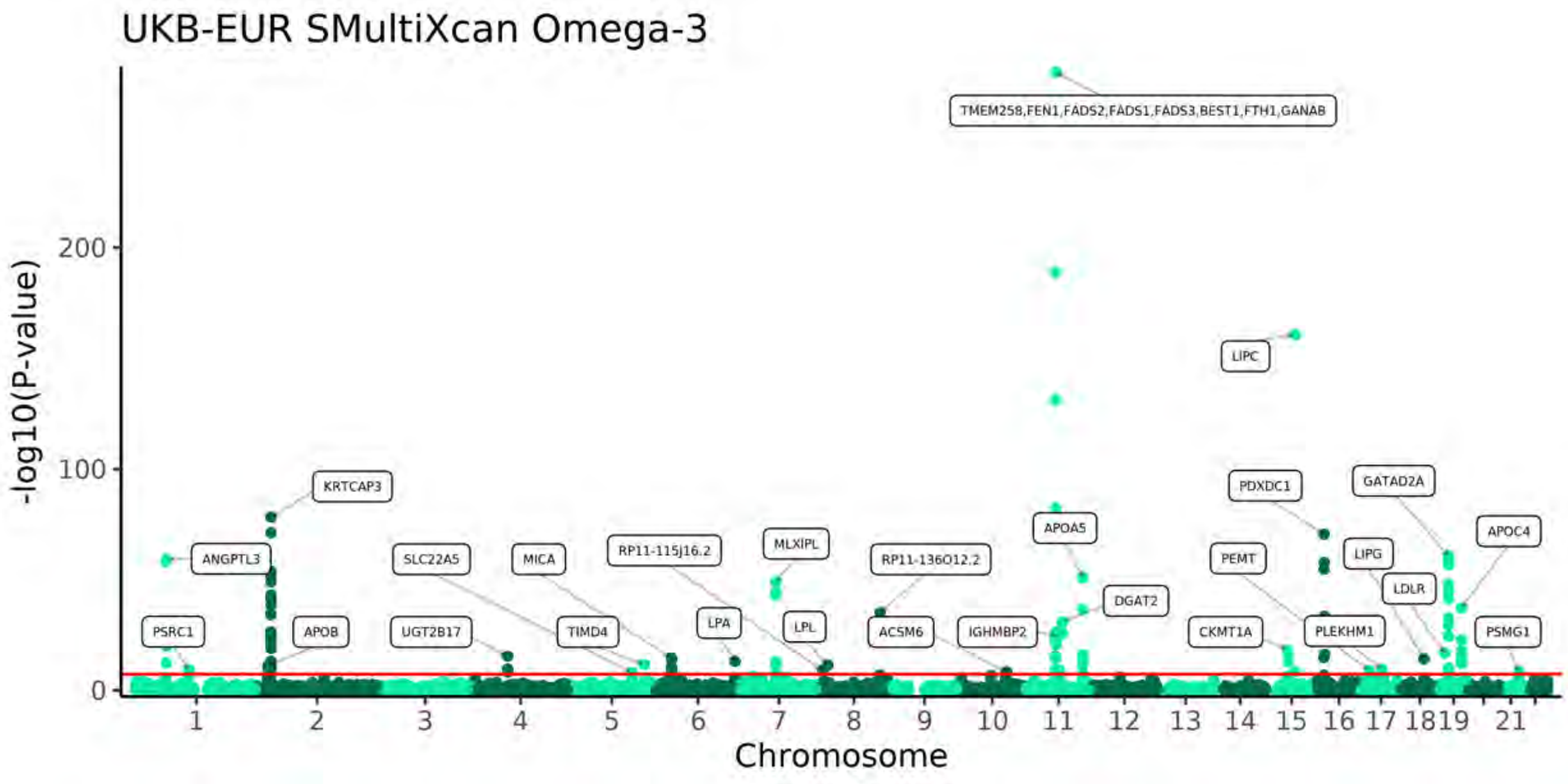
UKB-EUR SMultiXcan Omega-3
-log10(P-value)
Chromosome
TMEM258,FEN1,FADS2,FADS1,FADS3,BEST1,FTH1,GANAB
LIPC
KRTCAP3
PDXDC1
GATAD2A
APOA5
APOC4
ANGPTL3
SLC22A5
MICA
RP11-115J16.2
MLXIPL
RP11-136O12.2
PEMT
LIPG
LDLR
DGAT2
PSRC1
APOB
UGT2B17
TIMD4
LPA
LPL
ACSM6
IGHMBP2
CKMT1A
PLEKHM1
PSMG1

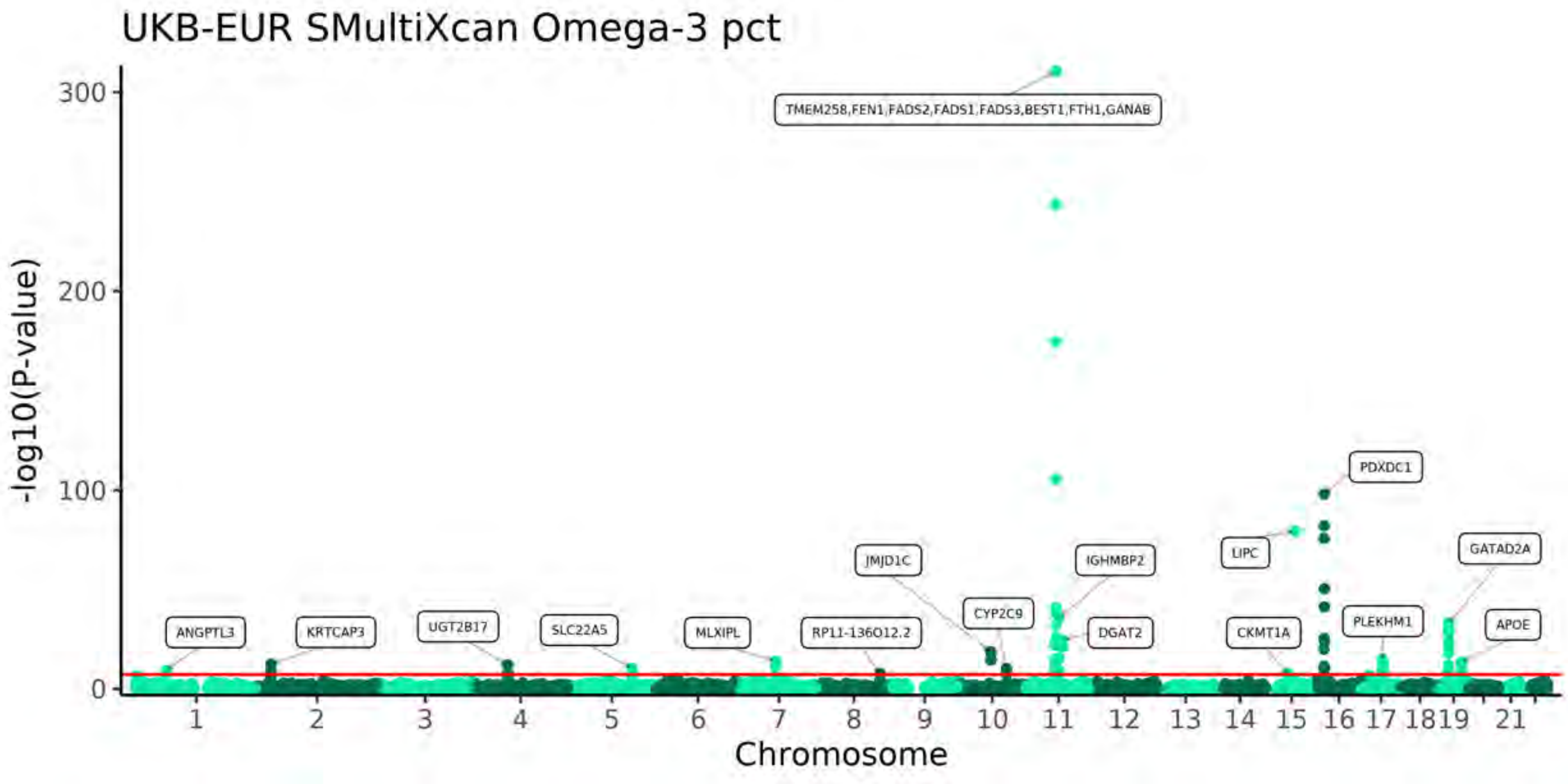
UKB-EUR SMultiXcan Omega-3 pct
-log10(P-value)
Chromosome
TMEM258,FEN1,FADS2,FADS1,FADS3,BEST1,FTH1,GANAB
PDXDC1
JMJD1C
IGHMBP2
LIPC
GATAD2A
CYP2C9
PLEKHM1
APOE
ANGPTL3
KRTCAP3
UGT2B17
SLC22A5
MLXIPL
RP11-136O12.2
DGAT2
CKMT1A

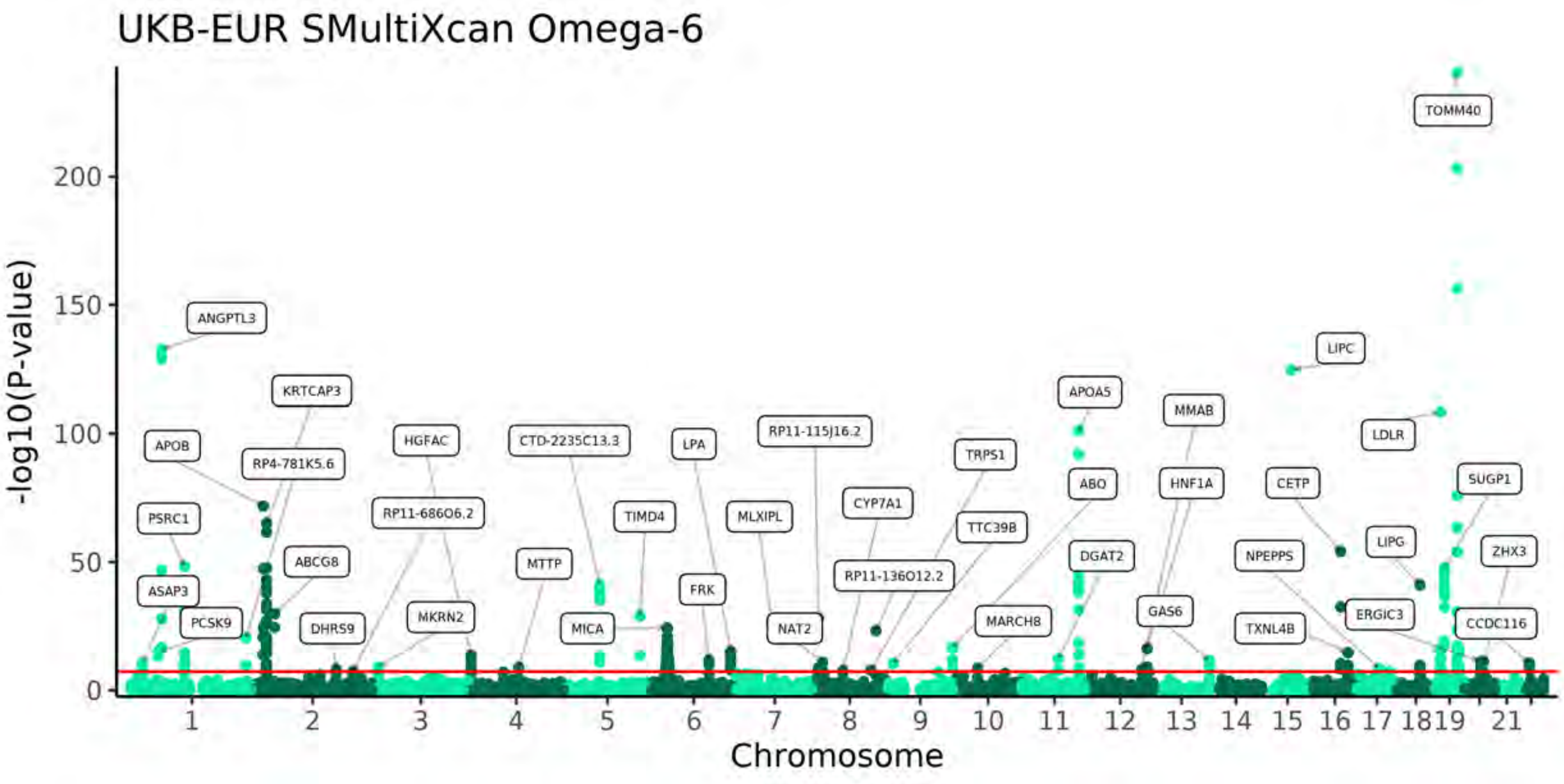
UKB-EUR SMultiXcan Omega-6
-log10(P-value)
Chromosome
0
50
100
150
200
1 2 3 4 5 6 7 8 9 10 11 12 13 14 15 16 17 18 19 21
ANGPTL3
KRTCAP3
APOB
RP4-781K5.6
PSRC1
ASAP3
PCSK9
ABCG8
DHRS9
HGFAC
RP11-686O6.2
MKRN2
CTD-2235C13.3
MTTP
MICA
TIMD4
LPA
FRK
MLXIPL
RP11-115J16.2
NAT2
CYP7A1
RP11-136O12.2
TRPS1
TTC39B
MARCH8
APOA5
ABO
DGAT2
MMAB
HNF1A
GAS6
LIPC
CETP
NPEPPS
TXNL4B
LDLR
LIPG
ERGIC3
TOMM40
SUGP1
ZHX3
CCDC116

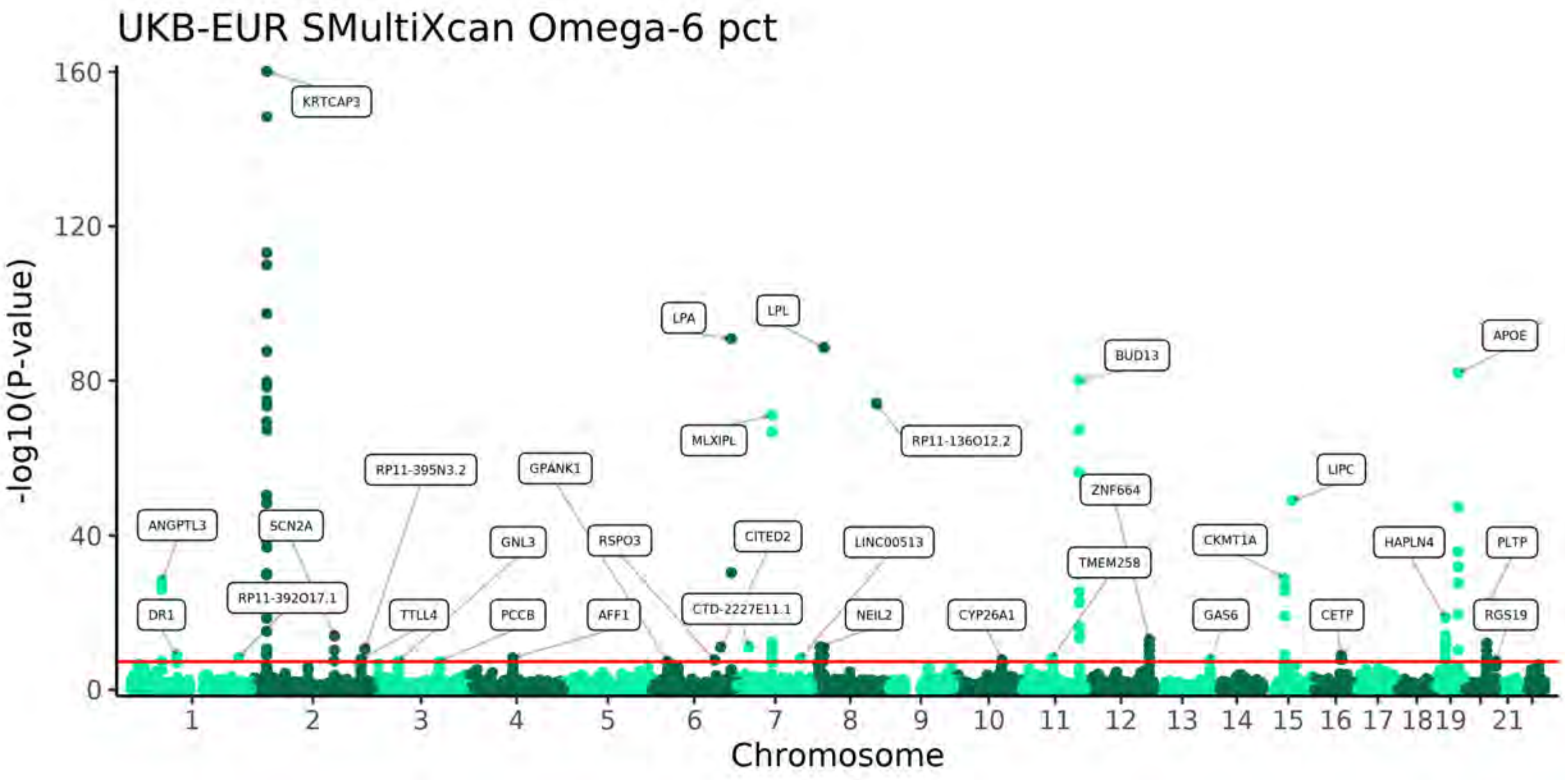
UKB-EUR SMultiXcan Omega-6 pct
-log10(P-value)
Chromosome
0
40
80
120
160
1 2 3 4 5 6 7 8 9 10 11 12 13 14 15 16 17 18 19 21
KRTCAP3
ANGPTL3
DR1
SCN2A
RP11-392O17.1
RP11-395N3.2
TTLL4
GNL3
PCCB
GPANK1
RSPO3
AFF1
LPA
MLXIPL
CITED2
CTD-2227E11.1
LPL
LINC00513
NEIL2
RP11-136O12.2
CYP26A1
BUD13
ZNF664
TMEM258
CKMT1A
GAS6
LIPC
CETP
HAPLN4
APOE
PLTP
RGS19

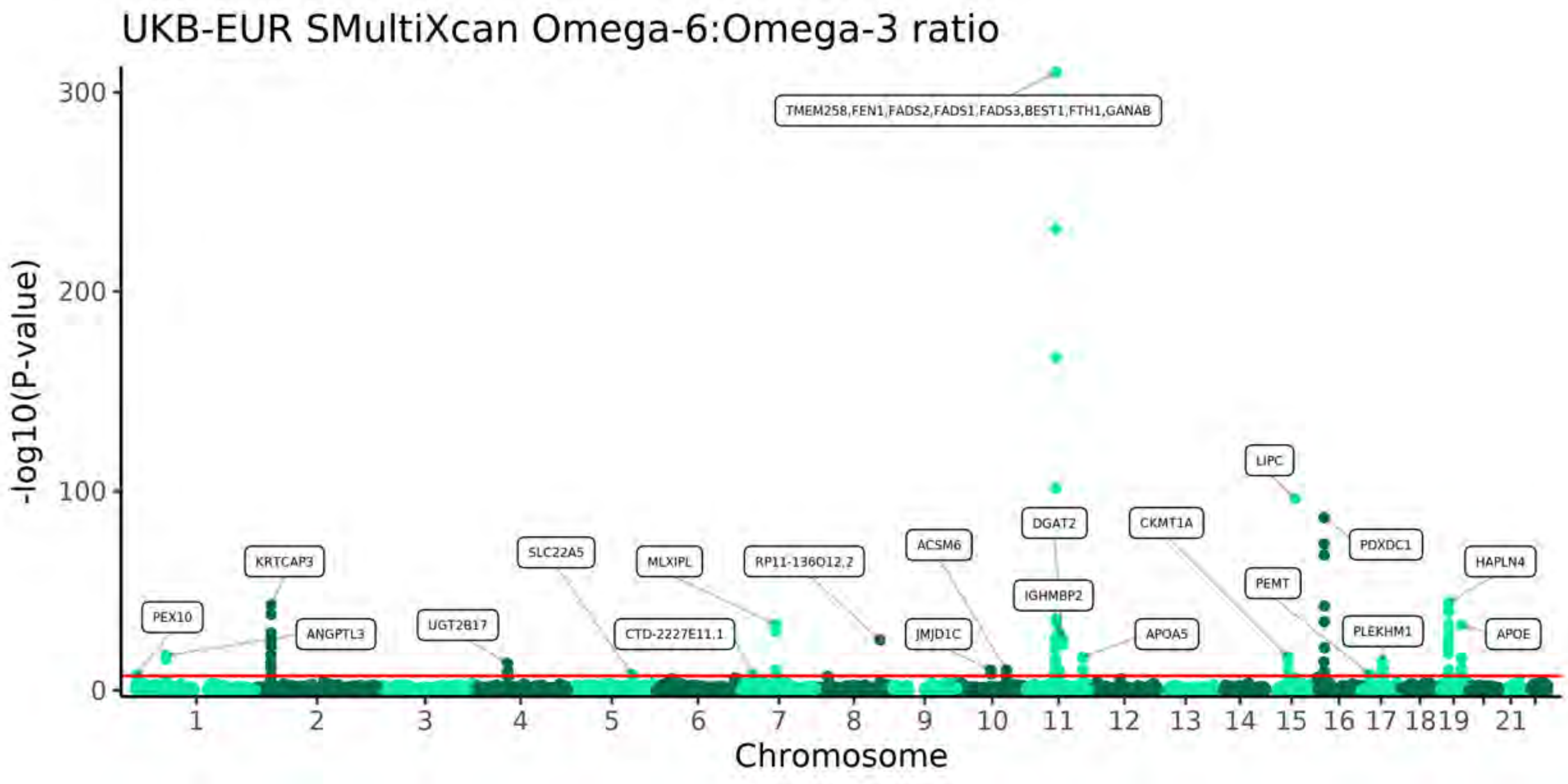
UKB-EUR SMultiXcan Omega-6:Omega-3 ratio
-log10(P-value)
Chromosome
TMEM258,FEN1,FADS2,FADS1,FADS3,BEST1,FTH1,GANAB
LIPC
DGAT2
CKMT1A
PDXDC1
HAPLN4
ACSM6
PEMT
IGHMBP2
KRTCAP3
SLC22A5
MLXIPL
RP11-136O12.2
PEX10
ANGPTL3
UGT2B17
CTD-2227E11.1
JMJD1C
APOA5
PLEKHM1
APOE

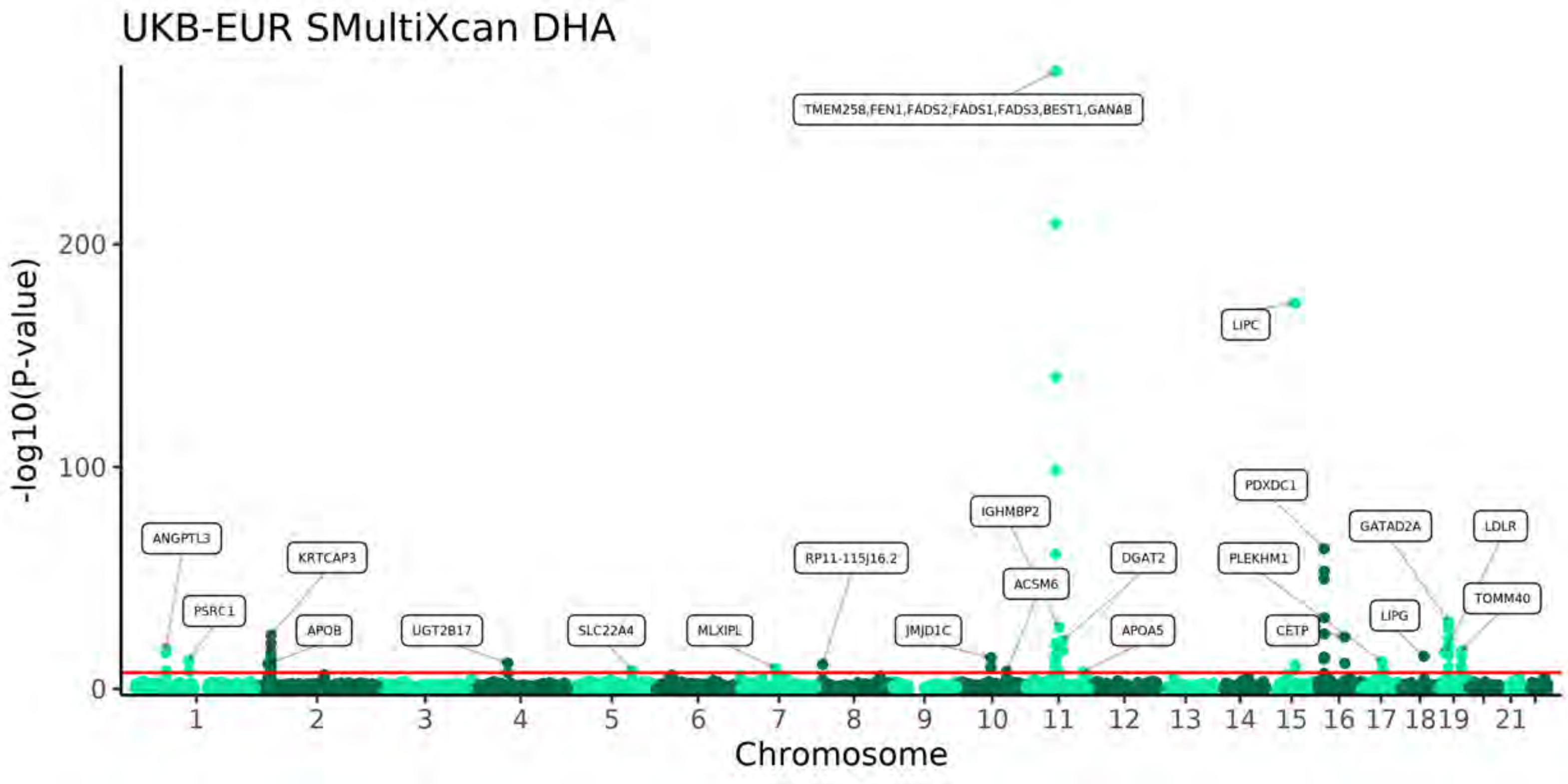
UKB-EUR SMultiXcan DHA
-log10(P-value)
Chromosome
TMEM258,FEN1,FADS2,FADS1,FADS3,BEST1,GANAB
LIPC
PDXDC1
IGHMBP2
GATAD2A
LDLR
ANGPTL3
KRTCAP3
RP11-115J16.2
DGAT2
PLEKHM1
ACSM6
TOMM40
PSRC1
LIPG
APOB
UGT2B17
SLC22A4
MLXIPL
JMJD1C
APOA5
CETP

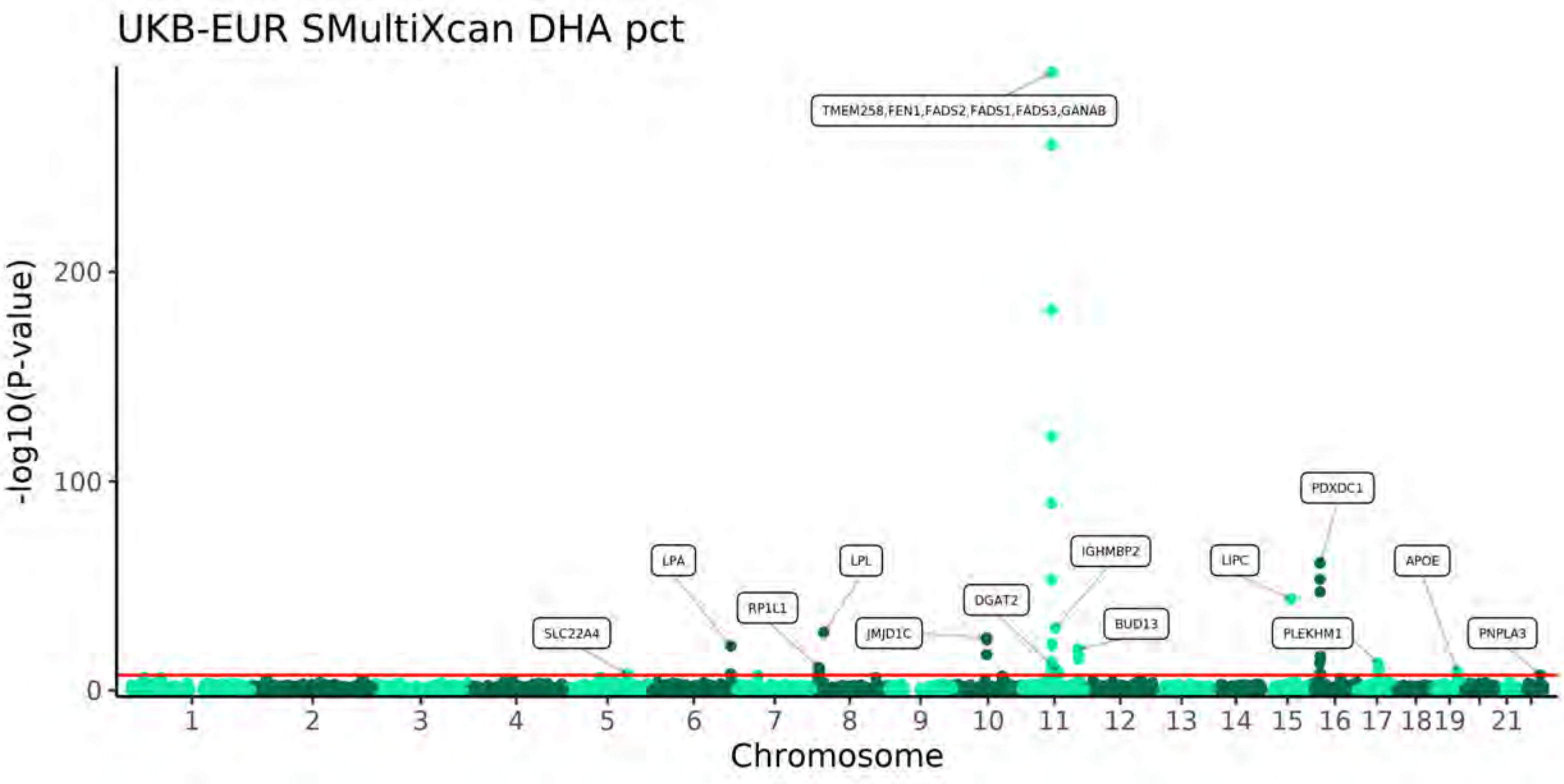
UKB-EUR SMultiXcan DHA pct
-log10(P-value)
0
100
200
Chromosome
21
TMEM258,FEN1,FADS2,FADS1,FADS3,GANAB
PDXDC1
LPA
LPL
IGHMBP2
LIPC
APOE
RP1L1
DGAT2
BUD13
SLC22A4
JMJD1C
PLEKHM1
PNPLA3

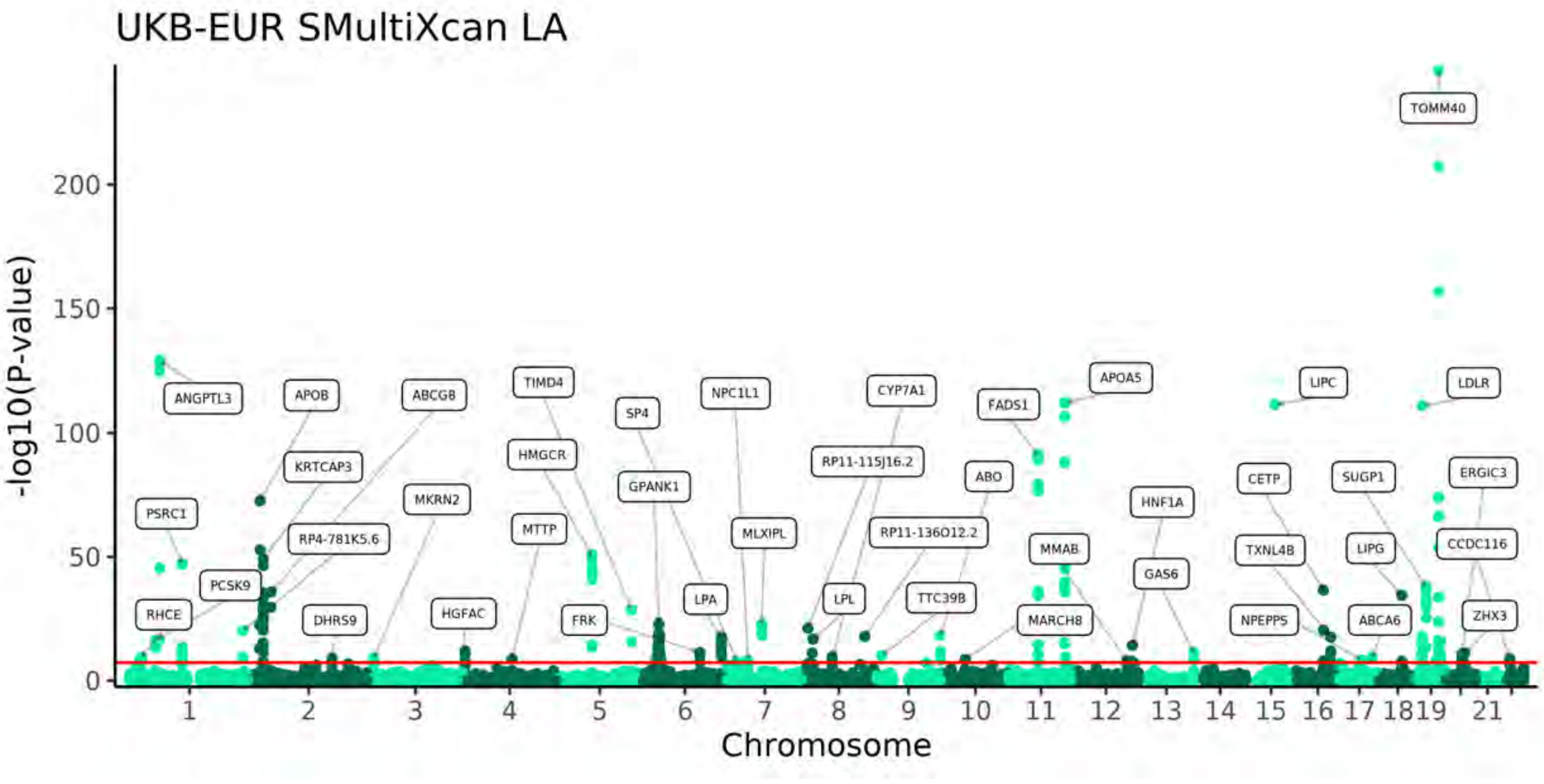
UKB-EUR SMultiXcan LA
-log10(P-value)
0
50
100
150
200
Chromosome
21
TOMM40
ANGPTL3
APOB
ABCG8
TIMD4
SP4
NPC1L1
CYP7A1
FADS1
APOA5
LIPC
LDLR
KRTCAP3
HMGCR
GPANK1
RP11-115J16.2
ABO
CETP
SUGP1
ERGIC3
PSRC1
MKRN2
RP4-781K5.6
MTTP
MLXIPL
RP11-136O12.2
HNF1A
MMAB
TXNL4B
LIPG
CCDC116
PCSK9
GAS6
RHCE
DHRS9
HGFAC
FRK
LPA
LPL
TTC39B
MARCH8
NPEPPS
ABCA6
ZHX3

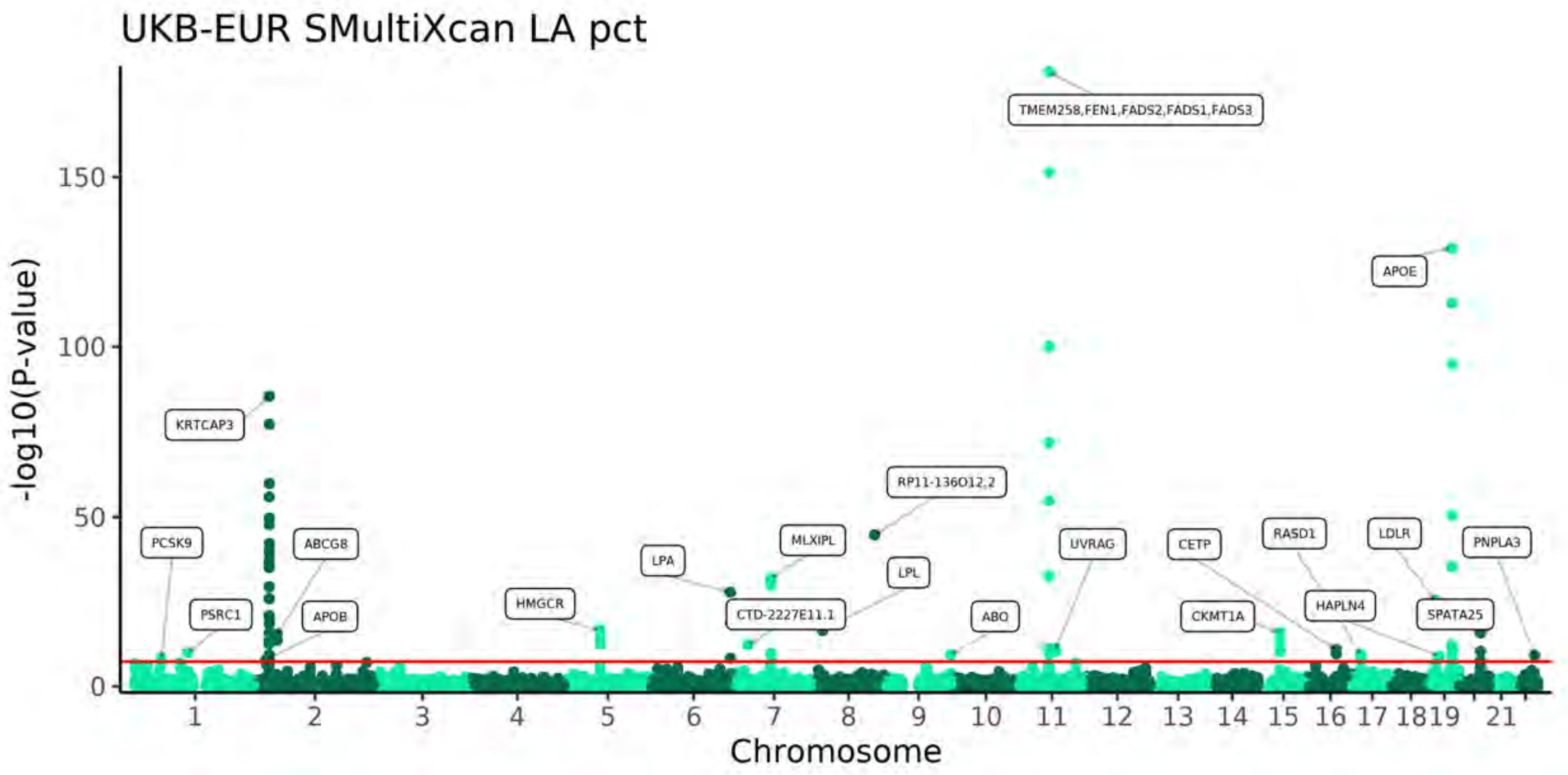
UKB-EUR SMultiXcan LA pct
-log10(P-value)
0
50
100
150
Chromosome
1 2 3 4 5 6 7 8 9 10 11 12 13 14 15 16 17 1819 21
TMEM258,FEN1,FADS2,FADS1,FADS3
APOE
KRTCAP3
RP11-136O12.2
PCSK9
ABCG8
LPA
MLXIPL
UVRAG
CETP
RASD1
LDLR
PNPLA3
LPL
PSRC1
APOB
HMGCR
CTD-2227E11.1
ABO
CKMT1A
HAPLN4
SPATA25

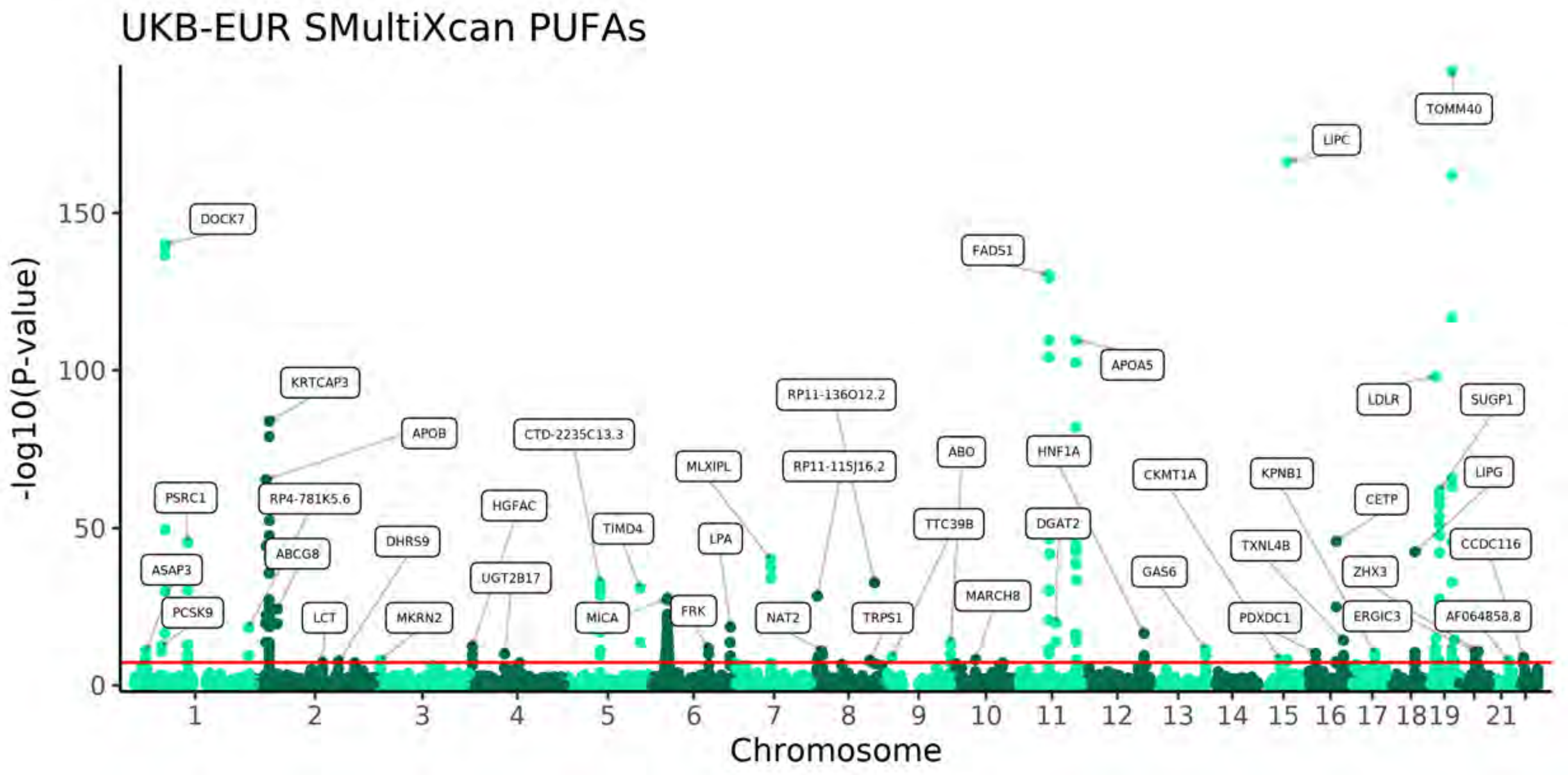
UKB-EUR SMultiXcan PUFAs
-log10(P-value)
0
50
100
150
Chromosome
1 2 3 4 5 6 7 8 9 10 11 12 13 14 15 16 17 1819 21
TOMM40
LIPC
DOCK7
FADS1
APOA5
KRTCAP3
RP11-136O12.2
LDLR
SUGP1
APOB
CTD-2235C13.3
ABO
HNF1A
MLXIPL
RP11-115J16.2
CKMT1A
KPNB1
LIPG
PSRC1
RP4-781K5.6
HGFAC
TIMD4
TTC39B
DGAT2
CETP
LPA
TXNL4B
CCDC116
ABCG8
DHRS9
ASAP3
UGT2B17
ZHX3
GAS6
MARCH8
PCSK9
LCT
MKRN2
MICA
FRK
NAT2
TRPS1
PDXDC1
ERGIC3
AF064858.8

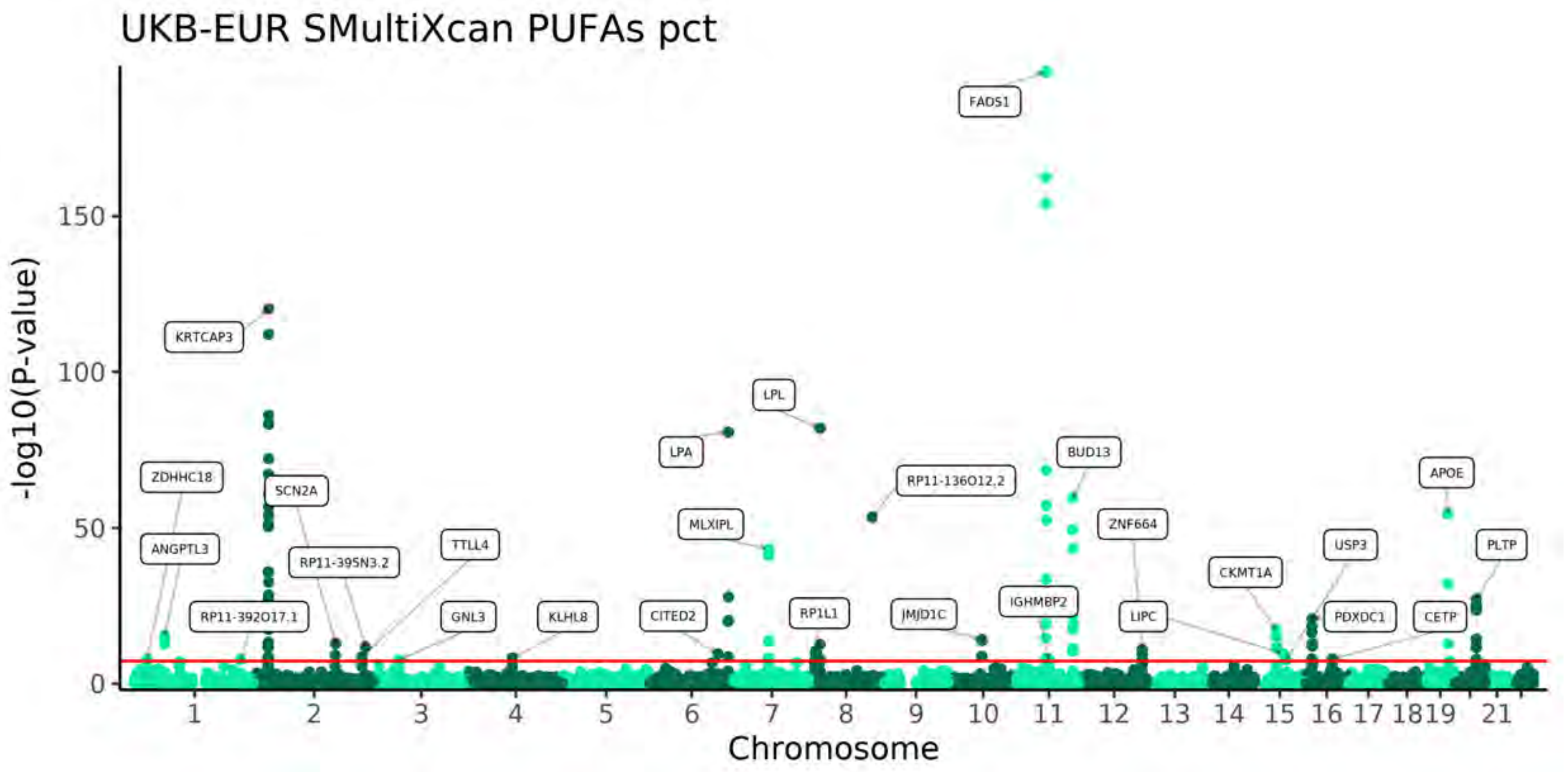
UKB-EUR SMultiXcan PUFAs pct
-log10(P-value)
0
50
100
150
Chromosome
1 2 3 4 5 6 7 8 9 10 11 12 13 14 15 16 17 1819 21
FADS1
KRTCAP3
LPL
LPA
BUD13
ZDHHC18
SCN2A
RP11-136O12.2
APOE
MLXIPL
ZNF664
ANGPTL3
TTLL4
USP3
PLTP
RP11-395N3.2
CKMT1A
IGHMBP2
RP11-392O17.1
GNL3
KLHL8
CITED2
RP1L1
JMJD1C
LIPC
PDXDC1
CETP

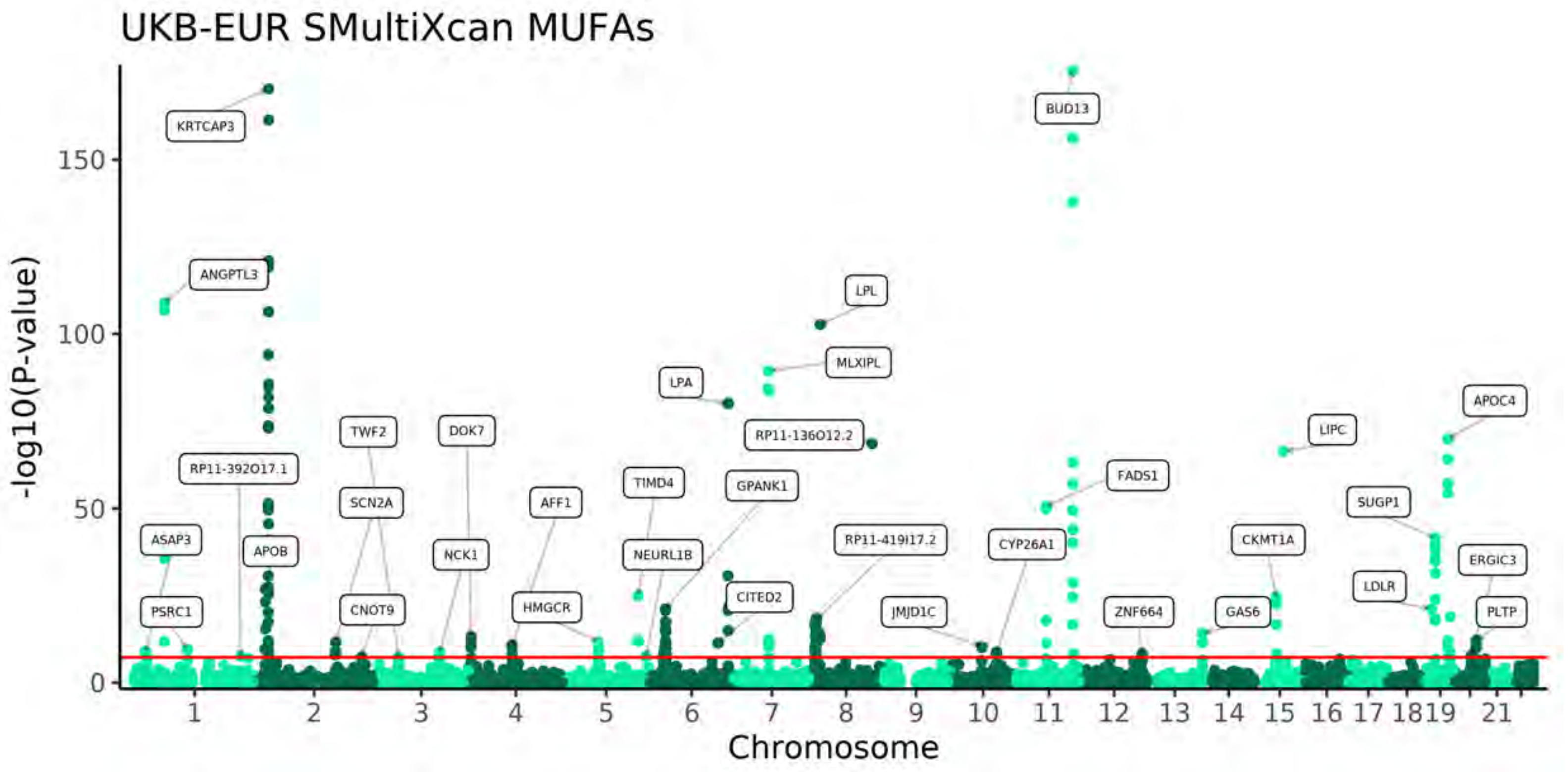
UKB-EUR SMultiXcan MUFAs
-log10(P-value)
0
50
100
150
Chromosome
1 2 3 4 5 6 7 8 9 10 11 12 13 14 15 16 17 1819 21
KRTCAP3
BUD13
ANGPTL3
LPL
MLXIPL
LPA
APOC4
TWF2
DOK7
RP11-136O12.2
LIPC
RP11-392O17.1
TIMD4
GPANK1
FADS1
SCN2A
AFF1
SUGP1
ASAP3
APOB
NCK1
NEURL1B
RP11-419I17.2
CYP26A1
CKMT1A
ERGIC3
LDLR
CITED2
PSRC1
CNOT9
HMGCR
JMJD1C
ZNF664
GAS6
PLTP

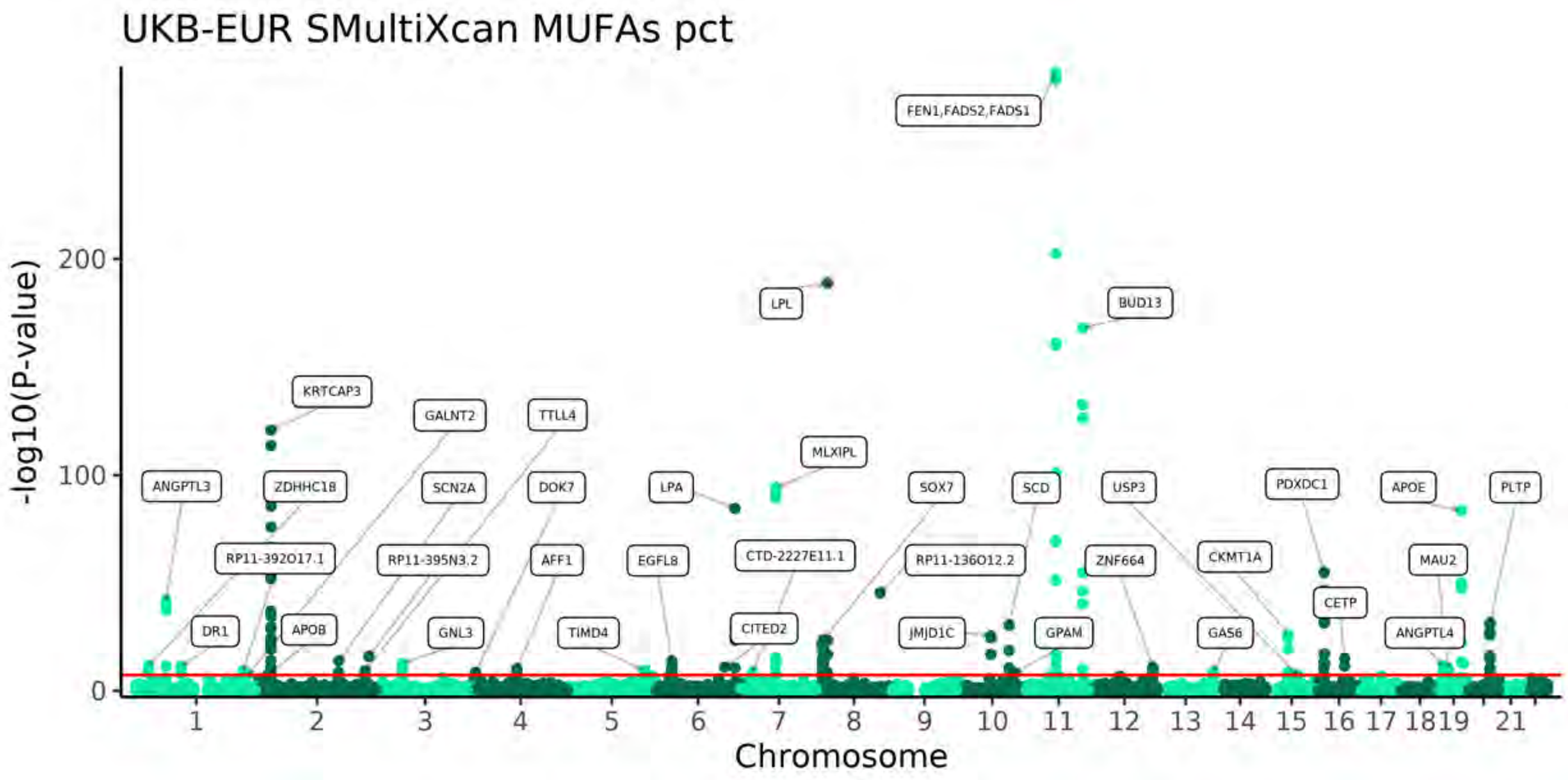
UKB-EUR SMultiXcan MUFAs pct
-log10(P-value)
Chromosome
FEN1,FADS2,FADS1
LPL
BUD13
KRTCAP3
GALNT2
TTLL4
MLXIPL
ANGPTL3
ZDHHC18
SCN2A
DOK7
LPA
SOX7
SCD
USP3
PDXDC1
APOE
PLTP
RP11-392O17.1
RP11-395N3.2
AFF1
EGFL8
CTD-2227E11.1
RP11-136O12.2
ZNF664
CKMT1A
MAU2
CETP
DR1
APOB
GNL3
TIMD4
CITED2
JMJD1C
GPAM
GAS6
ANGPTL4

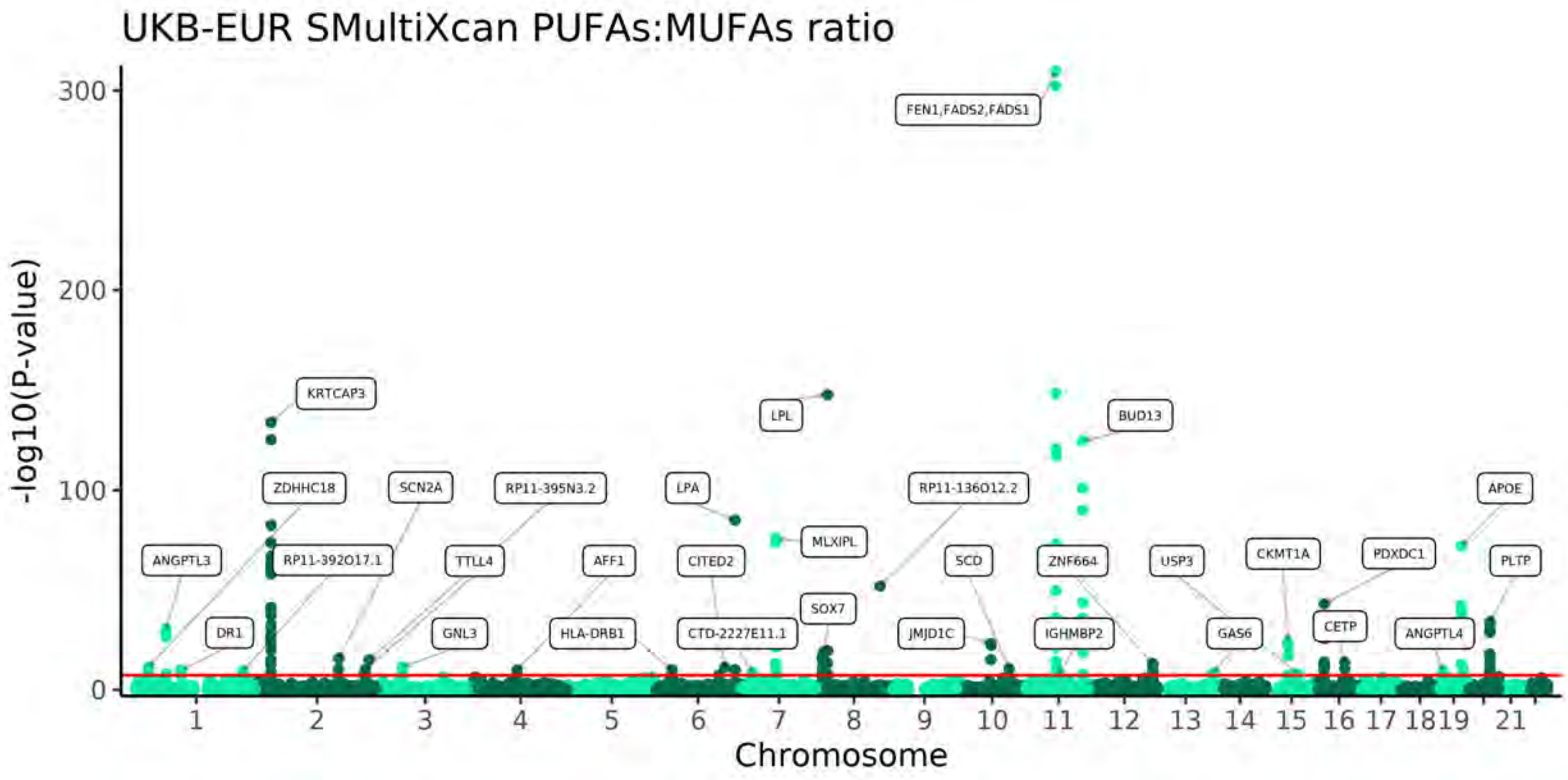
UKB-EUR SMultiXcan PUFAs:MUFAs ratio
-log10(P-value)
Chromosome
FEN1,FADS2,FADS1
KRTCAP3
LPL
BUD13
ZDHHC18
SCN2A
RP11-395N3.2
LPA
RP11-136O12.2
APOE
ANGPTL3
RP11-392O17.1
TTLL4
AFF1
CITED2
MLXIPL
SCD
ZNF664
USP3
CKMT1A
PDXDC1
PLTP
SOX7
DR1
GNL3
HLA-DRB1
CTD-2227E11.1
JMJD1C
IGHMBP2
GAS6
CETP
ANGPTL4

# S7

UKB-EUR + FinMetSeq + Ketunnen et al.
MAGMA Tissue Expression Analysis
Omega-3

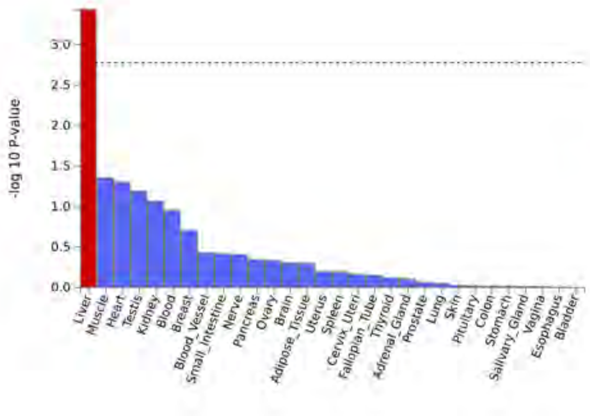

UKB-EUR + FinMetSeq + Ketunnen et al.
MAGMA Tissue Expression Analysis
DHA

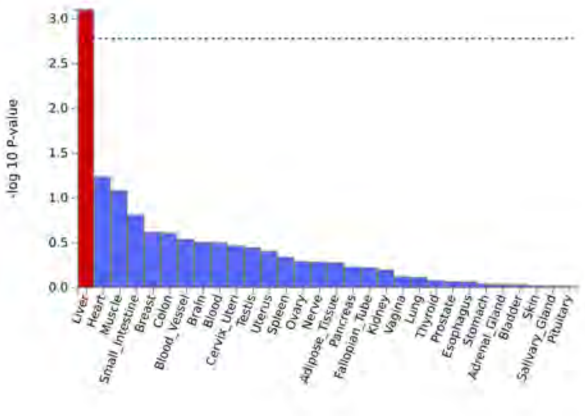

UKB-EUR + FinMetSeq + Ketunnen et al.
MAGMA Tissue Expression Analysis
Omega-6

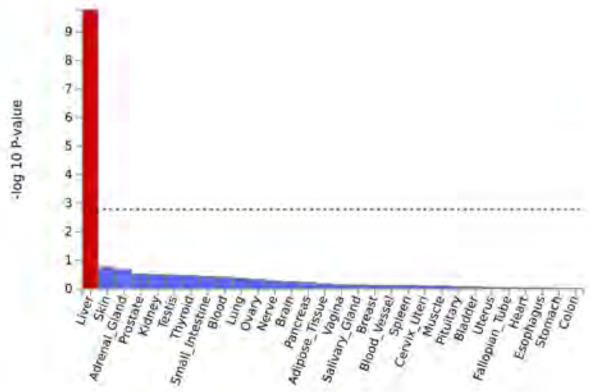

UKB-EUR + FinMetSeq + Ketunnen et al.
MAGMA Tissue Expression Analysis
LA

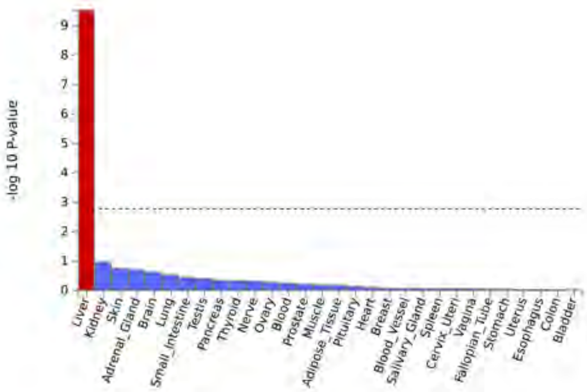

UKB-EUR + FinMetSeq + Ketunnen et al.
MAGMA Tissue Expression Analysis
MUFAs

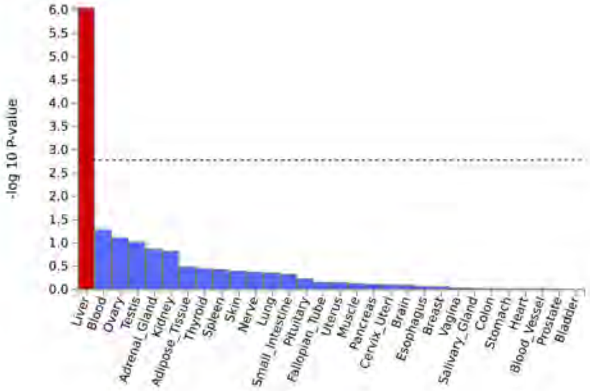
